# The Effect of Large-Scale Anti-Contagion Policies on the COVID-19 Pandemic

**DOI:** 10.1101/2020.03.22.20040642

**Authors:** Solomon Hsiang, Daniel Allen, Sébastien Annan-Phan, Kendon Bell, Ian Bolliger, Trinetta Chong, Hannah Druckenmiller, Luna Yue Huang, Andrew Hultgren, Emma Krasovich, Peiley Lau, Jaecheol Lee, Esther Rolf, Jeanette Tseng, Tiffany Wu

## Abstract

Governments around the world are responding to the novel coronavirus (COVID-19) pandemic^1^ with unprecedented policies designed to slow the growth rate of infections. Many actions, such as closing schools and restricting populations to their homes, impose large and visible costs on society, but their benefits cannot be directly observed and are currently understood only through process-based simulations.^2–4^ Here, we compile new data on 1,717 local, regional, and national non-pharmaceutical interventions deployed in the ongoing pandemic across localities in China, South Korea, Italy, Iran, France, and the United States (US). We then apply reduced-form econometric methods, commonly used to measure the effect of policies on economic growth, ^5,6^ to empirically evaluate the effect that these anti-contagion policies have had on the growth rate of infections. In the absence of policy actions, we estimate that early infections of COVID-19 exhibit exponential growth rates of roughly 38% per day. We find that anti-contagion policies have significantly and substantially slowed this growth. Some policies have different impacts on different populations, but we obtain consistent evidence that the policy packages now deployed are achieving large, beneficial, and measurable health outcomes. We estimate that across these six countries, interventions prevented or delayed on the order of 62 million confirmed cases, corresponding to averting roughly 530 million total infections. These findings may help inform whether or when these policies should be deployed, intensified, or lifted, and they can support decision-making in the other 180+ countries where COVID-19 has been reported.^7^

## Introduction

The COVID-19 pandemic is forcing societies worldwide to make consequential policy decisions with limited information. After containment of the initial outbreak failed, attention turned to implementing non-pharmaceutical interventions designed to slow contagion of the virus. In general, these policies aim to decrease virus transmission by reducing contact among individuals within or between populations, such as by closing restaurants or restricting travel, thereby slowing the spread of COVID-19 to a manageable rate. These large-scale anti-contagion policies are informed by epidemiological simulations^2, 4, 8, 9^ and a small number of natural experiments in past epidemics.^10^ However, the actual effects of these policies on infection rates in the ongoing pandemic are unknown. Because the modern world has never confronted this pathogen, nor deployed anti-contagion policies of such scale and scope, it is crucial that direct measurements of policy impacts be used alongside numerical simulations in current decision-making.

Societies around the world are weighing whether the health benefits of anti-contagion policies are worth their social and economic costs. Many of these costs are plainly seen; for example, business restrictions increase unemployment and school closures impact educational outcomes. It is therefore not surprising that some populations have hesitated before implementing such dramatic policies, especially when their costs are visible while their health benefits – infections and deaths that would have occurred but instead were avoided or delayed – are unseen. Our objective is to measure the direct health benefits of these policies; specifically, how much these policies slowed the growth rate of infections. To do this, we compare the growth rate of infections within hundreds of sub-national regions before and after each of these policies is implemented locally. Intuitively, each administrative unit observed just prior to a policy deployment serves as the “control” for the same unit in the days after it receives a policy “treatment” (see Supplementary Information for accounts of these deployments). Our hope is to learn from the recent experience of six countries where early spread of the virus triggered large-scale policy actions, in part so that societies and decision-makers in the remaining 180+ countries can access this information.

Here we directly estimate the effects of 1, 717 local, regional, and national policies on the growth rate of infections across localities within China, France, Iran, Italy, South Korea, and the US (see Figure 1 and Supplementary Table 1). We compile subnational data on daily infection rates, changes in case definitions, and the timing of policy deployments, including (1) travel restrictions, (2) social distancing through cancellations of events and suspensions of educational/commercial/religious activities, (3) quarantines and lockdowns, and (4) additional policies such as emergency declarations and expansions of paid sick leave, from the earliest available dates to April 6, 2020 (see Supplementary Notes, also Extended Data Fig. 1). During this period, populations remained almost entirely susceptible to COVID-19, causing the natural spread of infections to exhibit almost perfect exponential growth.^11, 12^ The rate of this exponential growth could change daily, determined by epidemiological factors, such as disease infectivity, as well as policies that alter behavior.^9, 11, 13^ Because policies were deployed while the epidemic unfolded, we can estimate their effects empirically. To do this, we examine how the daily growth rate of infections in each locality changes in response to the collection of ongoing policies applied to that locality on that day.

**Figure 1:**
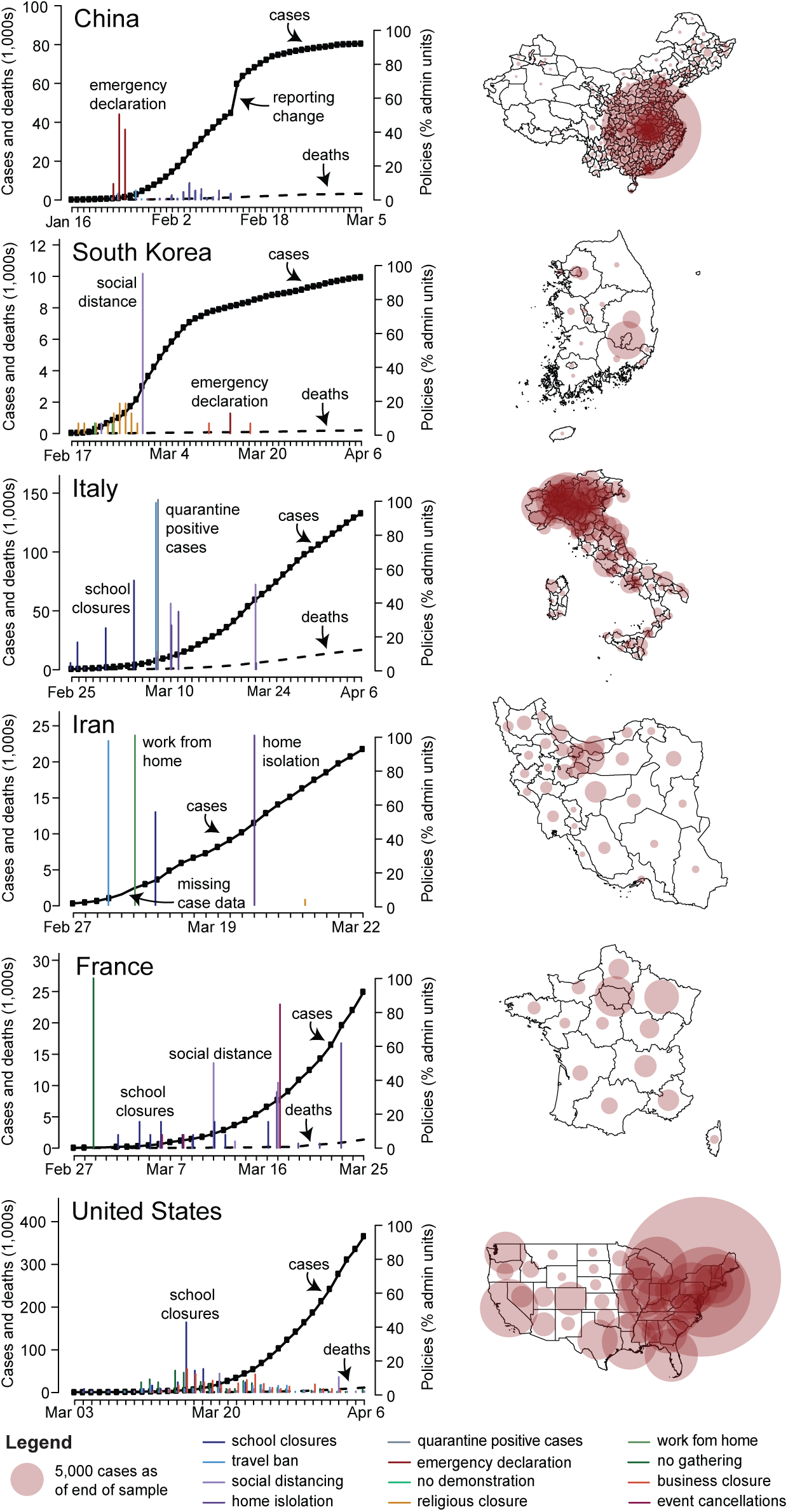
Data on COVID-19 infections and large-scale anti-contagion policies. Left: Daily cumulative confirmed cases of COVID-19 (solid black line, left axis) and deaths (dashed black line) over time. Vertical lines are deployments of anti-contagion policies, with height indicating the number of administrative units instituting a policy that day (right axis). For display purposes only, ≤ 5 policy types are shown per country and missing case data are imputed unless all sub-national units are missing. Right: Maps of cumulative confirmed cases by administrative unit on the last date of each sample.

## Methods Summary

We employ well-established “reduced-form” econometric techniques^5, 14^ commonly used to measure the effects of events^6, 15^ on economic growth rates. Similar to early COVID-19 infections, economic output generally increases exponentially with a variable rate that can be affected by policies and other conditions. Here, this technique aims to measure the total magnitude of the effect of *changes in policy*, without requiring explicit prior information about fundamental epidemiological parameters or mechanisms, many of which remain uncertain in the current pandemic. Rather, the collective influence of these factors is empirically recovered from the data without modeling their individual effects explicitly (see Methods). Prior work on influenza, ^16^ for example, has shown that such statistical approaches can provide important complementary information to process-based models.

To construct the dependent variable, we transform location-specific, subnational time-series data on infections into first-differences of their natural logarithm, which is the *per-day growth rate of infections* (see Methods). We use data from first- or second-level administrative units and data on active or cumulative cases, depending on availability (see Supplementary Information). We employ widely-used panel regression models^5, 14^ to estimate how the daily growth rate of infections changes over time within a location when different combinations of large-scale policies are enacted (see Methods). Our econometric approach accounts for differences in the baseline growth rate of infections across subnational locations, which may be affected by time-invariant characteristics, such as demographics, socio-economic status, culture, and health systems; it accounts for systematic patterns in growth rates within countries unrelated to policy, such as the effect of the work-week; it is robust to systematic under-surveillance specific to each subnational unit; and it accounts for changes in procedures to diagnose positive cases (see Methods and Supplementary Information).

## Results

We estimate that in the absence of policy, early infection rates of COVID-19 grow 43% per day on average across these six countries (Standard Error [SE]= 5%), implying a doubling time of approximately 2 days. Country-specific estimates range from 34% per day in the US (SE= 7%) to 68% per day in Iran (SE= 9%). We cannot determine if the high estimate for Iran results from true epidemiological differences, data quality issues (see Methods), the concurrence of the initial outbreak with a major religious holiday and pilgrimage (see Supplementary Notes), or sampling variability. Excluding Iran, the average growth rate is 38% per day (SE= 5%). Growth rates in all five other countries are independently estimated to be very near this value (Figure 2a). These estimated values differ from observed average growth rates because the latter are confounded by the effects of policy. These growth rates are not driven by the expansion of testing or increasing rates of case detection (see Methods and Extended Data Fig. 2) nor by data from individual regions (Extended Data Fig. 3).

**Figure 2:**
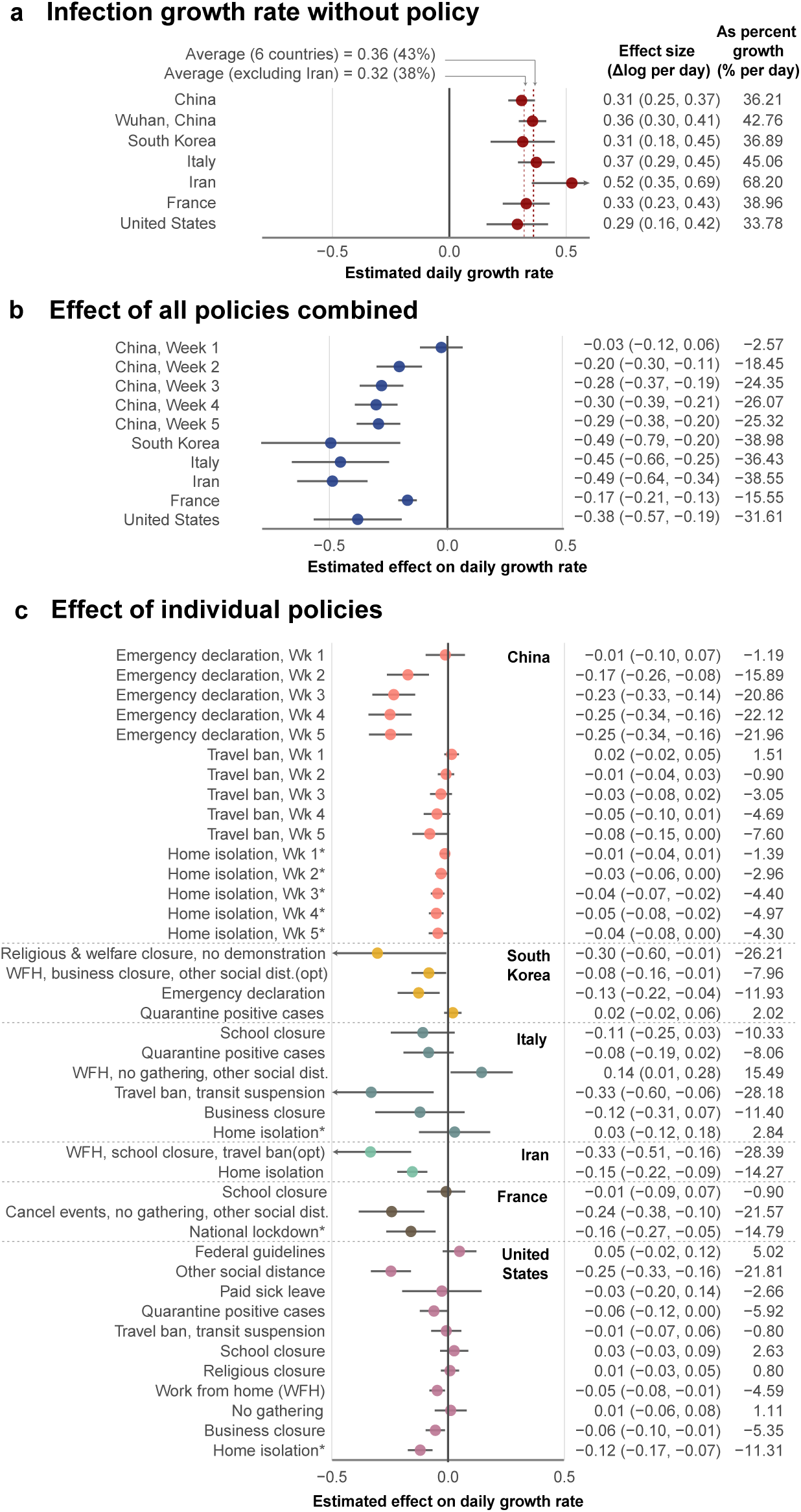
Empirical estimates of unmitigated COVID-19 infection growth rates and the effect of anti-contagion policies. Markers are country-specific estimates, whiskers are 95% CI. Columns report effect sizes as a change in the continuous-time growth rate (95% CI in parentheses) and the day-over-day percentage growth rate. (a) Estimates of daily COVID-19 infection growth rates in the absence of policy (dashed lines = averages with and without Iran, both excluding Wuhan-specific estimate). (b) Estimated combined effect of all policies on infection growth rates. (c) Estimated effects of individual policies or policy groups on the daily growth rate of infections, jointly estimated and ordered roughly chronologically within each country. *Reported effect of home isolation” includes effects of other implied policies (see Methods). China: N=3669; South Korea: N=595, Italy: N=2898, Iran: N=548, France: N=270, US: N=1238.

Some prior analyses of pre-intervention infections in Wuhan suggest slower growth rates (doubling every 5-7 days)^17, 18^ using data collected before national standards for diagnosis and case definitions were first issued by the Chinese government on January 15, 2020.^19^ However, case data in Wuhan from before this date contain multiple irregularities: the cumulative case count decreased on January 9; no new cases were reported during January 9-15; and there were concerns that information about the outbreak was suppressed^20^ (see Supplementary Table 2). When we remove these problematic data, utilizing a shorter but more reliable pre-intervention time series from Wuhan (January 16–21), we recover a growth rate of 43% per day (SE= 3%, doubling every 2 days) consistent with results from all other countries except Iran (Figure 2a, Supplementary Table 3).

During the early stages of an epidemic, a large proportion of the population remains susceptible to the virus, and if the spread of the virus is left uninhibited by policy or behavioral change, exponential growth continues until the fraction of the susceptible population declines meaningfully.^11, 13, 21, 22^ After correcting for estimated rates of case-detection, ^23^ we compute that the minimum susceptible fraction across administrative units in our sample is 72% of the total population (Cremona, Italy) and 87% of units would likely be in a regime of uninhibited exponential growth (> 95% susceptible) if policies were removed on the last date of our sample.

Consistent with predictions from epidemiological models, ^2, 10, 24^ we find that the combined effect of policies within each country reduces the growth rate of infections by a substantial and statistically significant amount (Figure 2b, Supplementary Table 3). For example, a locality in France with a baseline growth rate of 0.33 (national average) that fully deployed all policy actions used in France would be expected to lower its daily growth rate by -0.17 to a growth rate of 0.16. In general, the estimated total effects of policy packages are large enough that they can in principle offset a large fraction of, or even eliminate, the baseline growth rate of infections—although in several countries, many localities have not deployed the full set of policies. Overall, the estimated effects of all policies combined are generally insensitive to withholding regional (i.e. state- or province-level) blocks of data from the sample (Extended Data Fig. 3).

In China, only three policies were enacted across 116 cities early in a seven week period, providing us with sufficient data to empirically estimate how the effects of these policies evolved over time without making assumptions about the timing of these effects (see Methods and Fig. 2b). We estimate that the combined effect of these policies reduced the growth rate of infections by −0.026 (SE= 0.046) in the first week following their deployment, increasing substantially in the second week to -0.20 (SE= 0.049), and essentially stabilizing in the third week near −0.28 (SE= 0.047). In other countries, we lack sufficient data to estimate these temporal dynamics explicitly and only report the average pooled effect of policies across all days following their deployment (see Methods). If other countries have transient responses similar to China, we would expect effects in the first week following deployment to be smaller in magnitude than the average effect we report. In Extended Data Fig. 5a and Supplementary Methods Section 3, we explore how our estimates would change if we impose the assumption that policies cannot affect infection growth rates until after a fixed number of days; however, we do not find evidence this improves model fit.

The estimates above (Figure 2b) capture the superposition of all policies deployed in each country, i.e., they represent the average effect of policies that we would expect to observe if all policies enacted anywhere in each country were implemented simultaneously in a single region of that country. We also estimate the effects of individual policies or clusters of policies (Figure 2c) that are grouped based on either their similarity in goal (e.g., library and museum closures) or timing (e.g., policies deployed simultaneously). Our estimates for these individual effects tend to be statistically noisier than the estimates for all policies combined. Some estimates for the same policy differ between countries, perhaps because policies are not implemented identically or because populations behave differently. Nonetheless, 22 out of 29 point estimates indicate that individual policies are likely contributing to reducing the growth rate of infections. Seven policies (one in South Korea, two in Italy, and four in the US) have point estimates that are positive, six of which are small in magnitude (< 0.1) and not statistically different from zero (5% level). Consistent with greater overall uncertainty in these dis-aggregated estimates, some in China, South Korea, Italy, and France are somewhat more sensitive to withholding regional blocks of data (Extended Data Fig. 4), but remain broadly robust to assuming a constant delayed effect of all policies (Extended Data Fig. 5b).

Based on these results, we find that the deployment of anti-contagion policies in all six countries significantly and substantially slowed the pandemic. We combine the estimates above with our data on the timing of the 1, 717 policy deployments to estimate the total effect of all policies across the dates in our sample. To do this, we use our estimates to predict the growth rate of infections in each locality on each day, given the actual policies in effect at that location on that date (Figure 3, blue markers). We then use the same model to predict what counterfactual growth rates would be on that date if the effects of all policies were removed (Figure 3, red markers), which we call the “no-policy scenario.” The difference between these two predictions is our estimated effect that all deployed policies had on the growth rate of infections. During our sample, we estimate that all policies combined slowed the average growth rate of infections by −0.252 per day (SE= 0.045, p< 0.001) in China, −0.248 (SE= 0.089, p< 0.01) in South Korea, −0.24 (SE= 0.068, p< 0.001) in Italy, −0.355 (SE= 0.063, p< 0.001) in Iran, −0.123 (SE= 0.019, p< 0.001) in France and -0.084 (SE= 0.03, p< 0.01) in the US. These results are robust to modeling the effects of policies without grouping them (Extended Data Fig. 6a and Supplementary Table 4) or assuming a delayed effect of policy on infection growth rates (Supplementary Table 5).

**Figure 3:**
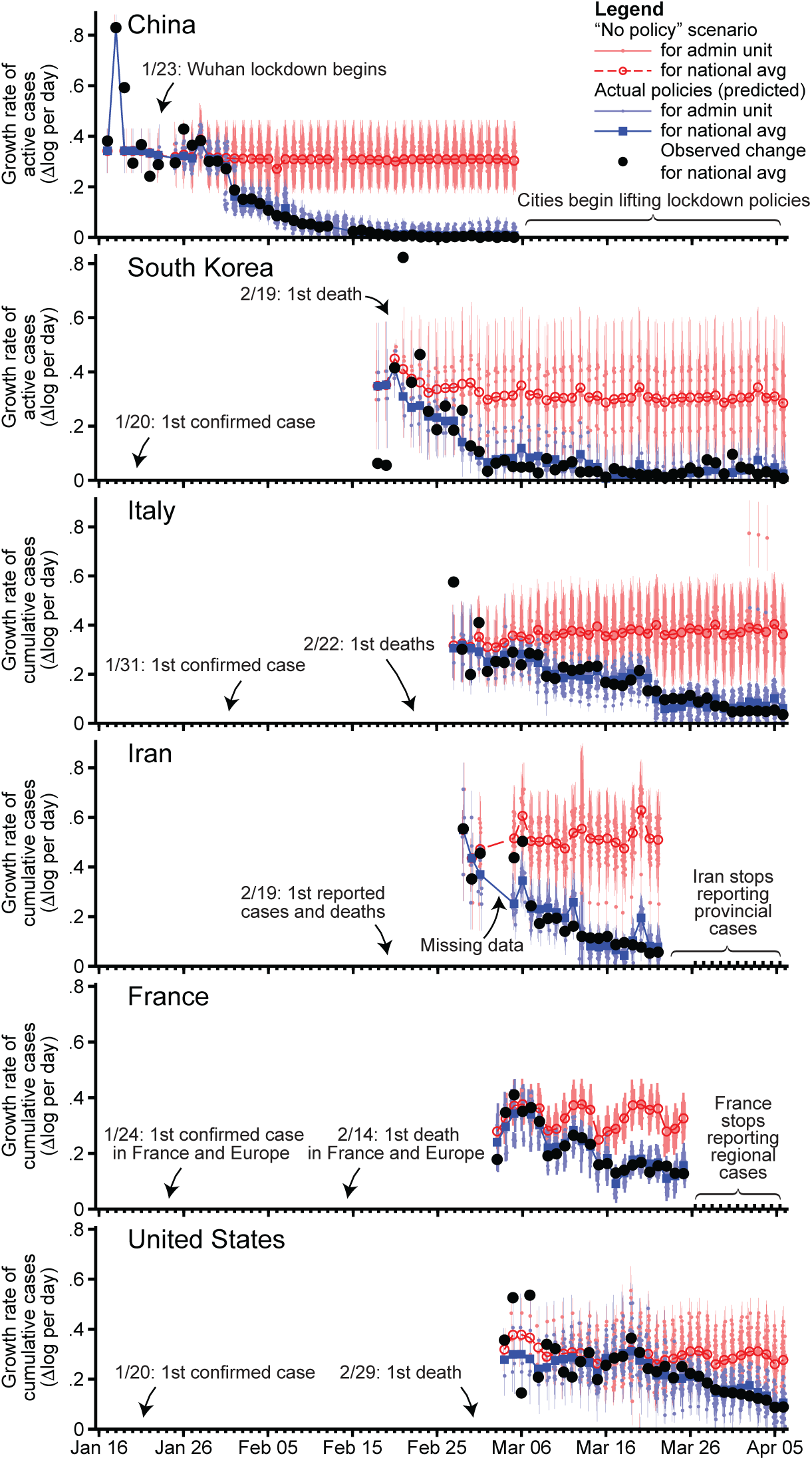
Estimated infection growth rates based on actual anti-contagion policies and in a “no policy” counterfactual scenario. Predicted daily growth rates of active (China, South Korea) or cumulative (all others) COVID-19 infections based on the observed timing of all policy deployments within each country (blue) and in a scenario where no policies were deployed (red). The difference between these two predictions is our estimated effect of actual anti-contagion policies on the growth rate of infections. Small markers are daily estimates for sub-national administrative units (vertical lines are 95% CI). Large markers are national averages. Black circles are observed daily changes in log(infections), averaged across administrative units. Sample sizes are the same as Figure 2.

The number of COVID-19 infections on a date depends on the growth rate of infections on all prior days. Thus, persistent reductions in growth rates have a compounding effect on infections, until growth is slowed by a shrinking susceptible population. To provide a sense of scale for our results, we integrate the growth rate of infections in each locality from Figure 3 to estimate cumulative infections, both with actual anti-contagion policies and in the no-policy counterfactual scenario. To account for the declining susceptible population in each administrative unit, we couple our econometric estimates of the effects of policies with a Susceptible-Infected-Removed (SIR) model^11, 13^ that adjusts the susceptible population in each administrative unit based on estimated case-detection rates^23, 25^ (see Methods). This allows us to extend our projections beyond the initial exponential growth phase of infections, a threshold that many localities cross in our no-policy scenario.

Our results suggest that ongoing anti-contagion policies have already substantially reduced the number of COVID-19 infections observed in the world today (Figure 4). Our central estimates suggest that there would be roughly 37 million more cumulative confirmed cases (corresponding to 285 million more total infections, including the confirmed cases) in China, 11.5 million more confirmed cases in South Korea (38 million total infections), 2.1 million more confirmed cases in Italy (49 million total infections), 5 million more confirmed cases in Iran (54 million total infections), 1.4 million more confirmed cases in France (45 million total infections), and 4.8 million more confirmed cases (60 million total infections) in the US had these countries never enacted any anti-contagion policies since the start of the pandemic. The magnitudes of these impacts partially reflect the timing, intensity, and extent of policy deployment (e.g., how many localities deployed policies), and the duration for which they have been applied. Several of these estimates are subject to large statistical uncertainties (see intervals in Figure 4). Sensitivity tests (Extended Data Fig. 7) that assume a range of plausible alternative parameter values relating to disease dynamics, such as incorporating a Susceptible-Exposed-Infected-Removed (SEIR) model, suggest that interventions may have reduced the severity of the outbreak by a total of 55-66 million confirmed cases over the dates in our sample (central estimates). Sensitivity tests varying the assumed infection-fatality ratio (Supplementary Table 6) suggest a corresponding range of 46-77 million confirmed cases (490-580 million total infections).

**Figure 4:**
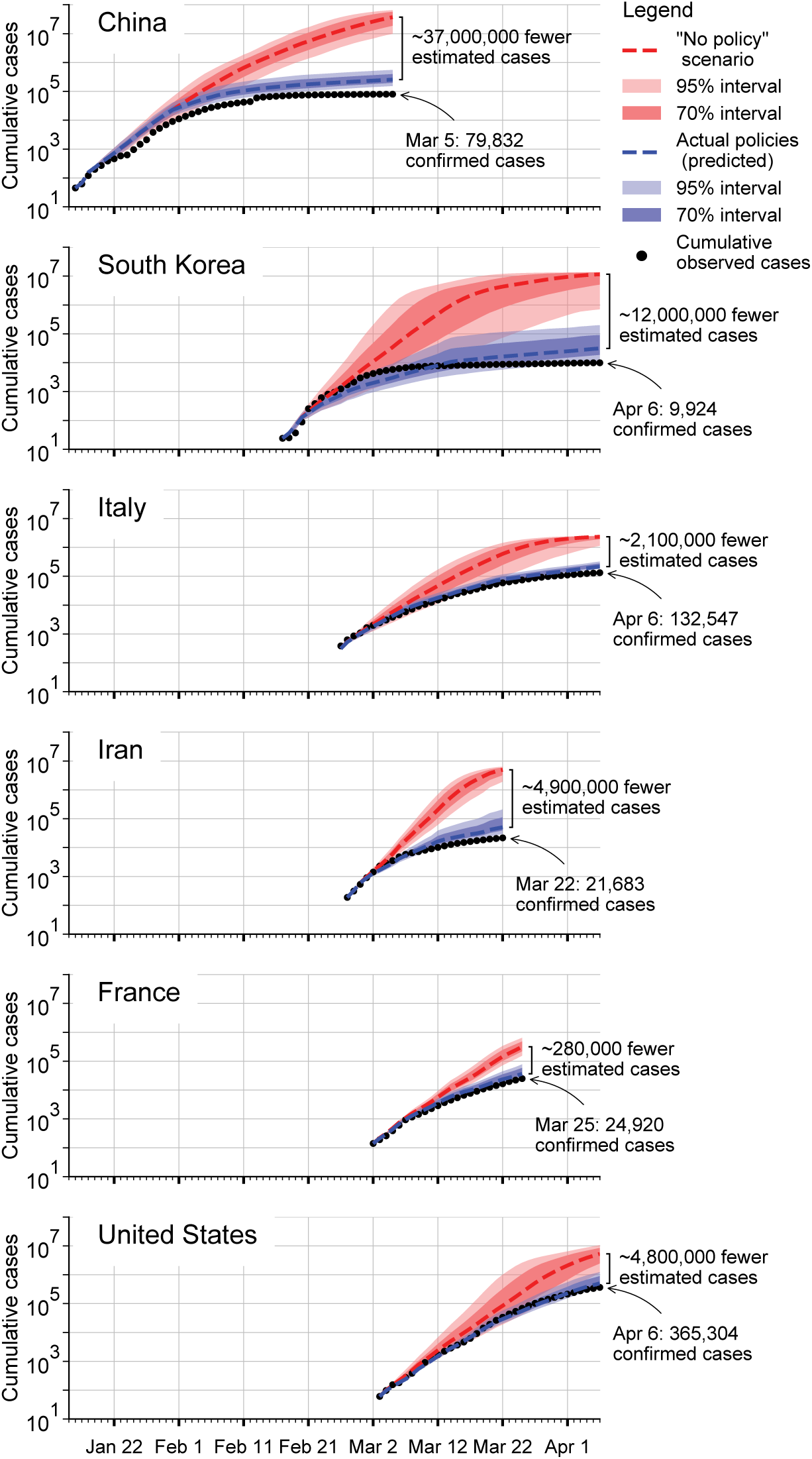
Estimated cumulative confirmed COVID-19 infections with and without anti-contagion policies. The predicted cumulative number of confirmed COVID-19 infections based on actual policy deployments (blue) and in the no-policy counterfactual scenario (red). Shaded areas show uncertainty based on 1, 000 simulations where empirically estimated parameters are resampled from their joint distribution (dark = inner 70% of predictions; light = inner 95%). Black dotted line is observed cumulative infections. Infections are not projected for administrative units that never report infections in the sample, but which might have experienced infections in a no-policy scenario.

## Discussion

Our empirical results indicate that large-scale anti-contagion policies are slowing the COVID-19 pandemic. Because infection rates in the countries we study would have initially followed rapid exponential growth had no policies been applied, our results suggest that these policies have provided large health benefits. For example, we estimate that there would be roughly 465× the observed number of confirmed cases in China, 17× in Italy, and 14× in the US by the end of our sample if large-scale anti-contagion policies had not been deployed. Consistent with process-based simulations of COVID-19 infections, ^2, 4, 8, 9, 22, 26^ our analysis of existing policies indicates that seemingly small delays in policy deployment likely produced dramatically different health outcomes.

While the limitations of available data pose challenges to our analysis, our aim is to use what data exist to estimate the first-order impacts of unprecedented policy actions in an ongoing global crisis. As more data become available, related findings will become more precise and may capture more complex interactions. Furthermore, this analysis does not account for interactions between populations in nearby localities, ^13^ nor mobility networks.^3, 4, 8, 9^ Nonetheless, we hope these results can support critical decision-making, both in the countries we study and in the other 180+ countries where COVID-19 infections have been reported.^7^

A key advantage of our reduced-form “top down” statistical approach is that it captures the real-world behavior of affected populations without requiring that we explicitly model underlying mechanisms and processes. This is useful in the current pandemic where many process-related parameters remain uncertain. However, our results cannot and should not be interpreted as a substitute for “bottom up” process-based epidemiological models specifically designed to provide guidance in public health crises. Rather, our results complement existing models, for example, by helping to calibrate key model parameters. We believe both forward-looking simulations and backward-looking empirical evaluations should be used to inform decision-making.

Our analysis measures changes in local infection growth rates associated with changes in anticontagion policies. A necessary condition for this association to be interpreted as the plausibly causal effect of these policies is that the timing of policy deployment is independent of infection growth rates.^14^ This assumption is supported by established epidemiological theory^11, 13, 27^ and evidence, ^28, 29^ which indicate that infections in the absence of policy will grow exponentially early in the epidemic, implying that pre-policy infection growth rates should be constant over time and therefore uncorrelated with the timing of policy deployment. Further, scientific guidance to decision-makers early in the current epidemic explicitly projected constant growth rates in the absence of anti-contagion measures, limiting the possibility that anticipated changes in natural growth rates affected decision-making.^2, 22, 30, 31^ In practice, policies tended to be deployed in response to high total numbers of cases (e.g. in France), ^32^ in response to outbreaks in other regions (e.g. in China, South Korea, and Iran), ^33^ after delays due to political constraints (e.g. in the US and Italy), and often with timing that coincided with arbitrary events, like weekends or holidays (see Supplementary Notes for detailed chronologies).

Our analysis accounts for documented changes in COVID-19 testing procedures and availability, as well as differences in case-detection across locations; however, unobserved trends in case-detection could affect our results (see Methods). We analyze estimated case-detection trends^23^ (Extended Data Fig. 2), finding that this potential bias is small, possibly elevating our estimated no-policy growth rates by 0.022 (7%) on average.

It is also possible that changing public knowledge during the period of our study affects our results. If individuals alter behavior in response to new information unrelated to anti-contagion policies, such as seeking out online resources, this could alter the growth rate of infections and thus affect our estimates. If increasing availability of information reduces infection growth rates, it would cause us to overstate the effectiveness of anti-contagion policies. We note, however, that if public knowledge is increasing in response to policy actions, such as through news reports, then it should be considered a pathway through which policies alter infection growth, not a form of bias. Investigating these potential effects is beyond the scope of this analysis, but it is an important topic for future investigations.

Finally, our analysis focuses on confirmed infections, but other outcomes, such as hospitalizations or deaths, are also of policy interest. Future work on these outcomes may require additional modeling approaches because they are relatively more context-and state-dependent. Nonetheless, we experimentally implement our approach on the daily growth rate of hospitalizations in France, where hospitalization data is available at the granularity of this study. We find that the total estimated effect of anti-contagion policies on the growth rate of hospitalizations is similar to our estimates for infection growth rates (Extended Data Fig. 6c).

## Data Availability

All data and code used in this analysis are available at https://github.com/bolliger32/gpl-covid. Updates are posted at http://www.globalpolicy.science/covid19.

## Methods

### Data Collection and Processing

We provide a brief summary of our data collection processes here (see the Supplementary Notes for more details, including access dates). Epidemiological, case definition/testing regime, and policy data for each of the six countries in our sample were collected from a variety of in-country data sources, including government public health websites, regional newspaper articles, and crowd-sourced information on Wikipedia. The availability of epidemiological and policy data varied across the six countries, and preference was given to collecting data at the most granular administrative unit level. The country-specific panel datasets are at the region level in France, the state level in the US, the province level in South Korea, Italy and Iran, and the city level in China. Due to data availability, the sample dates differ across countries: in China we use data from January 16 - March 5, 2020; in South Korea from February 17 - April 6, 2020; in Italy from February 26 - April 6, 2020; in Iran from February 27 - March 22, 2020; in France from February 29 - March 25, 2020; and in the US from March 3 - April 6, 2020. Below, we describe our data sources.

#### China

We acquired epidemiological data from an open source GitHub project^34^ that scrapes time series data from Ding Xiang Yuan. We extended this dataset back in time to January 10, 2020 by manually collecting official daily statistics from the central and provincial (Hubei, Guangdong, and Zhejiang) Chinese government websites. We compiled policies by collecting data on the start dates of travel bans and lockdowns at the city-level from the “2020 Hubei lockdowns” Wikipedia page^35^ and various other news reports. We suspect that most Chinese cities have implemented at least one anti-contagion policy due to their reported trends in infections; as such, we dropped cities where we could not identify a policy deployment date to avoid miscategorizing the policy status of these cities. Thus our results are only representative for the sample of 116 cities for which we obtained policy data.

#### South Korea

We manually collected and compiled the epidemiological dataset in South Korea, based on provincial government reports, policy briefings, and news articles. We compiled policy actions from news articles and press releases from the Korean Centers for Disease Control and Prevention (KCDC), the Ministry of Foreign Affairs, and local governments’ websites.

#### Iran

We used epidemiological data from the table “New COVID-19 cases in Iran by province”^36^ in the “2020 coronavirus pandemic in Iran” Wikipedia article, which were compiled from data provided on the Iranian Ministry of Health website (in Persian). We relied on news media reporting and two timelines of pandemic events in Iran^37,36^ to collate policy data. From March 2-3, Iran did not report subnational cases. Around this period the country implemented three national policies: a recommendation against local travel (3/1), work from home for government employees (3/3), and school closure (3/5). As the effects of these policies cannot be distinguished from each other due to the data gap, we group them for the purpose of this analysis.

#### Italy

We used epidemiological data from the GitHub repository^38^ maintained by the Italian Department of Civil Protection (Dipartimento della Protezione Civile). For policies, we primarily relied on the English version of the COVID-19 dossier “Chronology of main steps and legal acts taken by the Italian Government for the containment of the COVID-19 epidemiological emergency” written by the Dipartimento della Protezione Civile, ^39^ and Wikipedia.^40^

#### France

We used the region-level epidemiological dataset provided by France’s government website^41^ and supplemented it with numbers of confirmed cases by region on France’s public health website, which was previously updated daily through March 25.^42^ We obtained data on France’s policy response to the COVID-19 pandemic from the French government website, press releases from each regional public health site, ^43^ and Wikipedia. ^44^

#### United States

We used state-level epidemiological data from usafacts.org, ^45^ which they compile from multiple sources. For policy responses, we relied on a number of sources, including the U.S. Centers for Disease Control (CDC), the National Governors Association, as well as various executive orders from county-and city-level governments, and press releases from media outlets.

#### Policy Data

Policies in administrative units were coded as binary variables, where the policy was coded as either 1 (after the date that the policy was implemented, and before it was removed) or 0 otherwise, for the affected administrative units. When a policy only affected a fraction of an administrative unit (e.g., half of the counties within a state), policy variables were weighted by the percentage of people within the administrative unit who were treated by the policy. We used the most recent population estimates we could find for countries’ administrative units (see the Population Data section in the Appendix). In order to standardize policy types across countries, we mapped each country-specific policy to one of the broader policy category variables in our analysis. In this exercise, we collected 168 policies for China, 59 for South Korea, 214 for Italy, 23 for Iran, 59 for France, and 1, 194 for the United States (see Supplementary Table 1). There are some cases where we encode policies that are necessarily in effect whenever another policy is in place, due in particular to the far-reaching implications of home isolation policies. In China, wherever home isolation is documented, we assume a local travel ban is enacted on the same day if we have not found an explicit local travel ban policy for a given locality. In France, we assume home isolation is accompanied by event cancellations, social distancing, and no-gathering policies; in Italy, we assume home isolation entails no-gathering, local travel ban, work from home, and social distancing policies; in the US, we assume shelter-in-place orders indicate that non-essential business closures, work from home policies, and no-gathering policies are in effect. For policy types that are enacted multiple times at increasing degrees of intensity within a locality, we add weights to the variable by escalating the intensity from 0 pre-policy in steps up to 1 for the final version of the policy (see the Policy Data section in the Appendix).

#### Epidemiological Data

We collected information on cumulative confirmed cases, cumulative recoveries, cumulative deaths, active cases, and any changes to domestic COVID-19 testing regimes, such as case definitions or testing methodology. For our regression analysis (Figure 2), we use active cases when they are available (for China and South Korea) and cumulative confirmed cases otherwise. We document quality control steps in the Appendix. Notably, for China and South Korea we acquired more granular data than the data hosted on the Johns Hopkins University (JHU) interactive dashboard;^48^ we confirm that the number of confirmed cases closely match between the two data sources (see Extended Data Fig. 1). To conduct the econometric analysis, we merge the epidemiological and policy data to form a single data set for each country.

### Econometric analysis

#### Reduced-Form Approach

The reduced-form econometric approach that we apply here is a “top down” approach that describes the behavior of aggregate outcomes *y* in data (here, infection rates). This approach can identify plausibly causal effects^5, 14^ induced by exogenous changes in independent policy variables *z* (e.g., school closure) without explicitly describing all underlying mechanisms that link *z* to *y*, without observing intermediary variables *x* (e.g., behavior) that might link *z* to *y*, or without other determinants of y unrelated to *z* (e.g., demographics), denoted *w*. Let *f*(·) describe a complex and unobserved process that generates infection rates *y*:

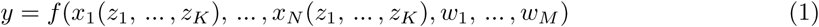

Process-based epidemiological models aim to capture elements of *f*(·) explicitly, and then simulate how changes in *z*, *x*, or *w* affect *y*. This approach is particularly important and useful in forward-looking simulations where future conditions are likely to be different than historical conditions. However, a challenge faced by this approach is that we may not know the full structure of *f*(·), for example if a pathogen is new and many key biological and societal parameters remain uncertain. Crucially, we may not know the effect that large-scale policy (*z*) will have on behavior (*x*(*z*)) or how this behavior change will affect infection rates (*f*(·)).

Alternatively, one can differentiate Equation 1 with respect to the *k*^th^ policy *z_k_*:

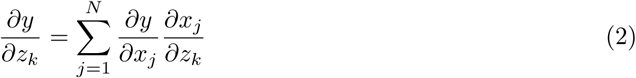

which describes how changes in the policy affects infections through all *N* potential pathways mediated by *x*_1_, …, *x_N_*. Usefully, for a fixed population observed over time, empirically estimating an average value of the local derivative on the left-hand-side in Equation 2 does not depend on explicit knowledge of *w*. If we can observe *y* and *z* directly and estimate changes over time 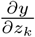: with data, then intermediate variables *x* also need not be observed nor modeled. The reduced-form econometric approach^5, 14^ thus attempts to measure 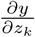 directly, exploiting exogenous variation in policies *z*.

#### Model

Active infections grow exponentially during the initial phase of an epidemic, when the proportion of immune individuals in a population is near zero. Assuming a simple Susceptible-Infected-Recovered (SIR) disease model (e.g., ref. [^11^]), the growth in infections during the early period is

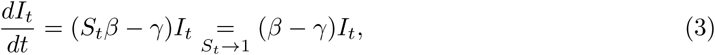

where *I_t_* is the number of infected individuals at time *t*, *β* is the transmission rate (new infections per day per infected individual), *γ* is the removal rate (proportion of infected individuals recovering or dying each day) and *S* is the fraction of the population susceptible to the disease. The second equality holds in the limit *S* → 1, which describes the current conditions during the beginning of the COVID-19 pandemic. The solution to this ordinary differential equation is the exponential function

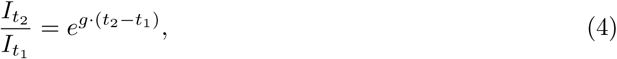

where *I_tl_* is the initial condition. Taking the natural logarithm and rearranging, we have

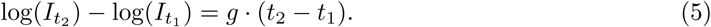

Anti-contagion policies are designed to alter *g*, through changes to *β*, by reducing contact between susceptible and infected individuals. Holding the time-step between observations fixed at one day (*t*_2_ − *t*_1_ = 1), we thus model g as a time-varying outcome that is a linear function of a time-varying policy

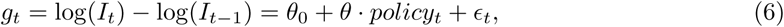

where *θ*_0_ is the average growth rate absent policy, *policy_t_* is a binary variable describing whether a policy is deployed at time *t*, and *θ* is the average effect of the policy on growth rate *g* over all periods subsequent to the policy’s introduction, thereby encompassing any lagged effects of policies. *∊_t_* is a mean-zero disturbance term that captures inter-period changes not described by *policy_t_*. Using this approach, infections each day are treated as the initial conditions for integrating Equation 4 through to the following day.

We compute the first differences log(*I_t_*) − log(*I_t−_*_1_) using active infections where they are available, otherwise we use cumulative infections, noting that they are almost identical during this early period (except in China, where we use active infections). We then match these data to policy variables that we construct using the novel data sets we assemble and apply a reduced-form approach to estimate a version of Equation 6, although the actual expression has additional terms detailed below.

#### Estimation

To estimate a multi-variable version of Equation 6, we estimate a separate regression for each country *c*. Observations are for subnational units indexed by *i* observed for each day *t*. Because not all localities began testing for COVID-19 on the same date, these samples are unbalanced panels. To ensure data quality, we restrict our analysis to localities after they have reported at least ten cumulative infections.

A necessary condition for unbiased estimates is that the timing of policy deployment is independent of natural infection growth rates, ^14^ a mathematical condition that should be true in the context of a new epidemic. In established epidemiological models, including the standard SIR model above, early rates of infection within a susceptible population are characterized by constant exponential growth. This phenomenon is well understood theoretically, ^13^, ^13, 27,46^ has been repeatedly documented in past epidemics^28, 29,47^ as well as the current COVID-19 pandemic^11,^, ^12^ and implies constant infection growth rates in the absence of policy intervention. Thus, we treat changes in infection growth rates as conditionally independent of policy deployments since the correlation between a constant variable and any other variable is zero in expectation.

We estimate a multiple regression version of Equation 6 using ordinary least squares. We include a vector of subnational unit-fixed effects *θ*_0_ (i.e., varying intercepts captured as coefficients to dummy variables) to account for all time-invariant factors that affect the local growth rate of infections, such as differences in demographics, socio-economic status, culture, and health systems.^5^ We include a vector of day-of-week-fixed effects *δ* to account for weekly patterns in the growth rate of infections that are common across locations within a country, however, in China, we omit day-of-week effects because we find no evidence they are present in the data – perhaps due to the fact that the outbreak of COVID-19 began during a national holiday and workers never returned to work. We also include a separate single-day dummy variable each time there is an abrupt change in the availability of COVID-19 testing or a change in the procedure to diagnose positive cases. Such changes generally manifest as a discontinuous jump in infections and a re-scaling of subsequent infection rates (e.g., See China in Figure 1), effects that are flexibly absorbed by a single-day dummy variable because the dependent variable is the first-difference of the logarithm of infections. We denote the vector of these testing dummies *μ*.

Lastly, we include a vector of *P_c_* country-specific policy variables for each location and day. These policy variables take on values between zero and one (inclusive) where zero indicates no policy action and one indicates a policy is fully enacted. In cases where a policy variable captures the effects of collections of policies (e.g., museum closures and library closures), a policy variable is computed for each, then they are averaged, so the coefficient on this type of variable is interpreted as the effect if all policies in the collection are fully enacted. There are also instances where multiple policies are deployed on the same date in numerous locations, in which case we group policies that have similar objectives (e.g., suspension of transit and travel ban, or cancelling of events and no gathering) and keep other policies separate (i.e., business closure, school closure). The grouping of policies is useful for reducing the number of estimated parameters in our limited sample of data, allowing us to examine the impact of subsets of policies (e.g. Fig. 2c). However, policy grouping does not have a material impact on the estimated effect of all policies combined nor on the effect of actual policies, which we demonstrate by estimating a regression model where no policies are grouped and these values are recalculated (Supplementary Table 4, Extended Data Fig. 6).

In some cases (for Italy and the US), policy data is available at a more spatially granular level than infection data (e.g., city policies and state-level infections in the US). In these cases, we code binary policy variables at the more granular level and use population-weights to aggregate them to the level of the infection data. Thus, policy variables may take on continuous values between zero and one, with a value of one indicating that the policy is fully enacted for the entire population. Given the limited quantity of data currently available, we use a parsimonious model that assumes the effects of policies on infection growth rates are approximately linear and additively separable. However, future work that possesses more data may be able to identify important nonlinearities or interactions between policies.

For each country, our general multiple regression model is thus

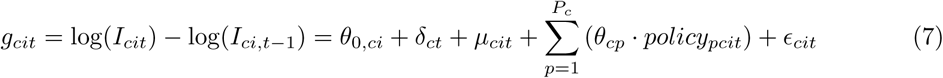

where observations are indexed by country c, subnational unit *i*, and day *t*. The parameters of interest are the country-by-policy specific coefficients *θ_cp_*. We display the estimated residuals *∊_cit_* in Extended Data Fig. 10, which are mean zero but not strictly normal (normality is not a requirement of our modeling and inference strategy), and we estimate uncertainty over all parameters by calculating our standard errors robust to error clustering at the day level.^14^ This approach allows the covariance in *∊_cit_* across different locations within a country, observed on the same day, to be nonzero. Such clustering is important in this context because idiosyncratic events within a country, such as a holiday or a backlog in testing laboratories, could generate nonuniform country-wide changes in infection growth for individual days not explicitly captured in our model. Thus, this approach non-parametrically accounts for both arbitrary forms of spatial auto-correlation or systematic misreporting in regions of a country on any given day (we note that it generates larger estimates for uncertainty than clustering by *i*). When we report the effect of all policies combined (e.g., Figure 2b) we are reporting the sum of coefficient estimates for all policies 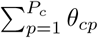, accounting for the covariance of errors in these estimates when computing the uncertainty of this sum.

Note that our estimates of *θ* and *θ*_0_ in Equation 7 are robust to systematic under-reporting of infections, a major concern in the ongoing pandemic, due to the construction of our dependent variable. This remains true even if different localities have different rates of under-reporting, so long as the rate of under-reporting is relatively constant. To see this, note that if each locality i has a medical system that reports only a fraction *ψi* of infections such that we observe *Ĩ_it_* = *ψ_i_I_it_* rather an actual infections *I_it_*, then the left-hand-side of Equation 7 will be

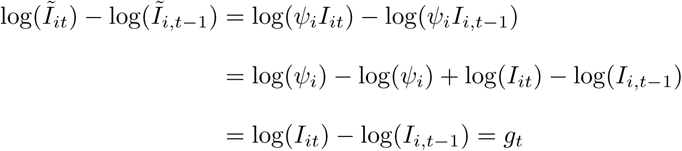

and is therefore unaffected by location-specific and time-invariant under-reporting. Thus systematic under-reporting does not affect our estimates for the effects of policy *θ*. As discussed above, potential biases associated with non-systematic under-reporting resulting from documented changes in testing regimes over space and time are absorbed by region-day specific dummies *μ*.

However, if the rate of under-reporting within a locality is changing day-to-day, this could bias infection growth rates. We estimate the magnitude of this bias (see Extended Data Fig. 2), and verify that it is quantitatively small. Specifically, if *Ĩ_it_* = *ψ_it_I_it_* where *ψ_it_* changes day-to-day, then

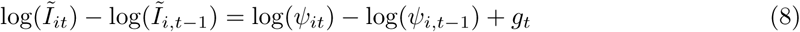

where log(*ψ_it_*)−log(*ψ_ijt−_*_1_) is the day-over-day growth rate of the case-detection probability. Disease surveillance has evolved slowly in some locations as governments gradually expand testing, which would cause *ψ_it_* to change over time, but these changes in testing capacity do not appear to significantly alter our estimates of infection growth rates. In Extended Data Fig. 2, we show one set of epidemiological estimates^23^ for log(*ψ_it_*) − log(*ψ_i, t−_*_1_). Despite random day-to-day variations, which do not cause systematic biases in our point estimates, the mean of log(*ψ_it_*) − log(*ψ_i, t_*_−1_) is consistently small across the different countries: 0.05 in China, 0.064 in Iran, 0.019 in South Korea, −0.058 in France, 0.031 in Italy, and 0.049 in the US. The average of these estimates is 0.026, potentially accounting for 7.3% of our global average estimate for the no-policy infection growth rate (0.36). These estimates of log(*ψ_it_*) − log(*ψ_i, t−_*_1_) also do not display strong temporal trends, alleviating concerns that time-varying under-reporting generates sizable biases in our estimated effects of anti-contagion policies.

#### Transient dynamics

In China, we are able to examine the transient response of infection growth rates following policy deployment because only three policies were deployed early in a seven-week sample period during which we observe many cities simultaneously. This provides us with sufficient data to estimate the temporal structure of policy effects without imposing assumptions regarding this structure. To do this, we estimate a distributed-lag model that encodes policy parameters using weekly lags based on the date that each policy is first implemented in locality i. This means the effect of a policy implemented one week ago is allowed to differ arbitrarily from the effect of that same policy in the following week, etc. These effects are then estimated simultaneously and are displayed in Fig. 2 (also Supplementary Table 3). Such a distributed lag approach did not provide statistically meaningful insight in other countries using currently available data because there were fewer administrative units and shorter periods of observation (i.e. smaller samples), and more policies (i.e. more parameters to estimate) in all other countries. Future work may be able to successfully explore these dynamics outside of China.

As a robustness check, we examine whether excluding the transient response from the estimated effects of policy substantially alters our results. We do this by estimating a “fixed lag” model, where we assume that policies cannot influence infection growth rates for *L* days, recoding a policy variable at time *t* as zero if a policy was implemented fewer than *L* days before *t*. We re-estimate Equation 7 for each value of *L* and present results in Extended Data Fig. 5 and Supplementary Table 5.

#### Alternative disease models

Our main empirical specification is motivated with an SIR model of disease contagion, which assumes zero latent period between exposure to COVID-19 and infectiousness. If we relax this assumption to allow for a latent period of infection, as in a Susceptible-Exposed-Infected-Recovered (SEIR) model, the growth of the outbreak is only asymptotically exponential.^11^ Nonetheless, we demonstrate that SEIR dynamics have only a minor potential impact on the coefficients recovered by using our empirical approach in this context. In Extended Data Figs. 8 and 9 we present results from a simulation exercise which uses Equations 9-11, along with a generalization to the SEIR model^11^ to generate synthetic outbreaks (see Supplementary Methods Section 2). We use these simulated data to test the ability of our statistical model (Equation 7) to recover both the unimpeded growth rate (Extended Data Fig. 8) as well as the impact of simulated policies on growth rates (Extended Data Fig. 9) when applied to data generated by SIR or SEIR dynamics over a wide range of epidemiological conditions.

### Projections

#### Daily growth rates of infections

To estimate the instantaneous daily growth rate of infections if policies were removed, we obtain fitted values from Equation 7 and compute a predicted value for the dependent variable when all *P_c_* policy variables are set to zero. Thus, these estimated growth rates 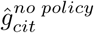 capture the effect of all locality-specific factors on the growth rate of infections (e.g., demographics), day-of-week-effects, and adjustments based on the way in which infection cases are reported. This counterfactual does not account for changes in information that are triggered by policy deployment, since those should be considered a pathway through which policies affect outcomes, as discussed in the main text. Additionally, the “no-policy” counterfactual does not model previously unobserved changes in behavior that might occur if fundamentally new behaviors emerge even in the absence of government intervention. When we report an average no-policy growth rate of infections (Figure 2a), it is the average value of these predictions for all observations in the original sample. Location-and-day specific counterfactual predictions (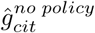), accounting for the covariance of errors in estimated parameters, are shown as red markers in Figure 3.

#### Cumulative infections

To provide a sense of scale for the estimated cumulative benefits of effects shown in Figure 3, we link our reduced-form empirical estimates to the key structures in a simple SIR system and simulate this dynamical system over the course of our sample. The system is defined as the following:

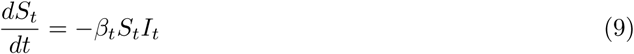

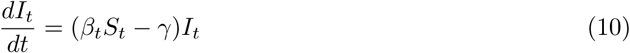

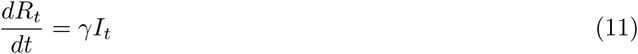

where *S_t_* is the susceptible population and *R_t_* is the removed population. Here *β_t_* is a time-evolving parameter, determined via our empirical estimates as described below. Accounting for changes in *S* becomes increasingly important as the size of cumulative infections (*I_t_* + *R_t_*) becomes a substantial fraction of the local subnational population, which occurs in some no-policy scenarios. Our reduced-form analysis provides estimates for the growth rate of active infections (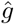) for each locality and day, in a regime where *S_t_* ≈ 1. Thus we know

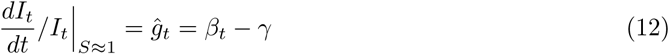

but we do not know the values of either of the two right-hand-side terms, which are required to simulate Equations 9-11. To estimate *γ*, we note that the left-hand-side term of Equation 11 is

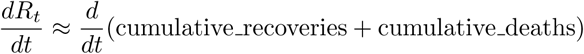

which we can observe in our data for China and South Korea. Computing first differences in these two variables (to differentiate with respect to time), summing them, and then dividing by active cases gives us estimates of *γ* (medians: China=0.11, Korea=0.05). These values differ slightly from the classical SIR interpretation of *γ* because in the public data we are able to obtain, individuals are coded as “recovered” when they no longer test positive for COVID-19, whereas in the classical SIR model this occurs when they are no longer infectious. We adopt the average of these two medians, setting *γ* = .08. We use medians rather than simple averages because low values for *I* induce a long right-tail in daily estimates of *γ* and medians are less vulnerable to this distortion. We then use our empirically-based reduced-form estimates of *ĝ* (both with and without policy) combined with Equations 9-11 to project total cumulative cases in all countries, shown in Figure 4. We simulate infections and cases for each administrative unit in our sample beginning on the first day for which we observe 10 or more cases (for that unit) using a time-step of 4 hours. Because we observe confirmed cases rather than total infections, we seed each simulation by adjusting observed *I_t_* on the first day using country-specific estimates of case detection rates. We adjust existing estimates of case under-reporting^23^ to further account for asymptomatic infections assuming an infection-fatality ratio of 0.075%.^25^ We assume *R_t_* = 0 on the first day. To maintain consistency with the reported data, we report our output in confirmed cases by multiplying our simulated *I_t_* + *R_t_* values by the aforementioned proportion of infections confirmed. We estimate uncertainty by resampling from the estimated variance-covariance matrix of all regression parameters. In Extended Data Fig. 7, we show sensitivity of this simulation to the estimated value of *γ* as well as to the use of a Susceptible-Exposed-Infected-Recovered (SEIR) framework. In Supplementary Table 6, we show sensitivity of this simulation to the assumed infection-fatality ratio (see Supplementary Methods Section 1).

## Data and Code Availability

The datasets generated during and/or analysed during the current study are available at https://github.com/bolliger32/gpl-covid (DOI: 10.5281/zenodo.3832367). For easier replication, we have also created a CodeOcean “capsule” - which contains a pre-built computing environment in addition to the source code and data. This is available at https://codeocean.com/capsule/1887579/tree/v1 (DOI: I0.24433/C0.6625287.v2). Future updates and/or extensions to data or code will be listed at http://www.globalpolicy.science/covid19.

## End Notes

## Acknowledgements

We thank Brenda Chen for her role initiating this work and Avi Feller for his feedback. Funding: SAP, EK, PL, JT are supported by a gift from the Tuaropaki Trust. TC is supported by an AI for Earth grant from National Geographic and Microsoft. DA, AH, IB are supported through joint collaborations with the Climate Impact Lab. KB is supported by the Royal Society Te Aparangi Rutherford Postdoctoral Fellowship. HD and ER are supported by the National Science Foundation Graduate Research Fellowship under Grant No. DGE 1106400 and 1752814, respectively. Opinions, findings, conclusions or recommendations expressed in this material are those of the authors and do not reflect the views of supporting organizations.

## Author Contributions

SH conceived of and led the study. All authors designed analysis, interpreted results, designed figures, and wrote the paper. China: LYH, TW collected health data, LYH, TW, JT collected policy data, LYH cleaned data. South Korea: JL Collected health data, TC, JL collected policy data, TC cleaned data. Italy: DA collected health data, PL collected policy data, DA cleaned data. France: SAP collected health data, SAP, JT, HD collected policy data, SAP cleaned data. Iran: AH collected health data and policy data, AH, DA cleaned data. USA: ER, KB collected health data, EK collected policy data, ER, DA, KB cleaned data. IB collected geographic and population data for all countries. SH designed the econometric model. SH, SAP, JT conducted econometric analysis for all countries. KB, IB, AH, ER, EK designed and implemented epidemiological models and projections. SAP, KB, IB, JT, AH, EK designed and implemented robustness checks. HD created Fig. 1, TC created Fig. 2, JT created Fig. 3, ER created Fig. 4, DA created SI Table 1, LYH, JL created SI Table 2, JT created SI Table 3, JT created SI Table 4, SAP, JT created SI Table 5, KB created SI Table 6, LYH created ED Figs. 1-2, SAP created ED Figs. 3-5, JT created ED Fig. 6, KB created ED Fig. 7, IB created ED Figs. 8-9, JT created ED Fig. 10. DA, IB, PL managed policy data collection and quality control. IB, TC managed the code repository. IB, PL ran project management. EK, TW, JT, PL managed literature review. LYH, EK, TW managed References. PL managed the Extended Data and Appendix.

## Ethics Declaration

The authors declare no conflicts of interest.

## Additional Information

Supplementary Information is available for this paper. Correspondence and requests for materials should be addressed to Solomon Hsiang.

## Extended Data for The Effect of Large-Scale Anti-Contagion Policies on the COVID-19 Pandemic”

All data and code are available at https://github.com/bolliger32/gpl-covid.

Additional resources and updates are available at www.globalpolicy.science/covid19.

For repository related questions, please contact.

For data related questions, please contact.

This document last updated: May 16, 2020. Data last updated: April 6, 2020.

**Extended Data Figure 1.**
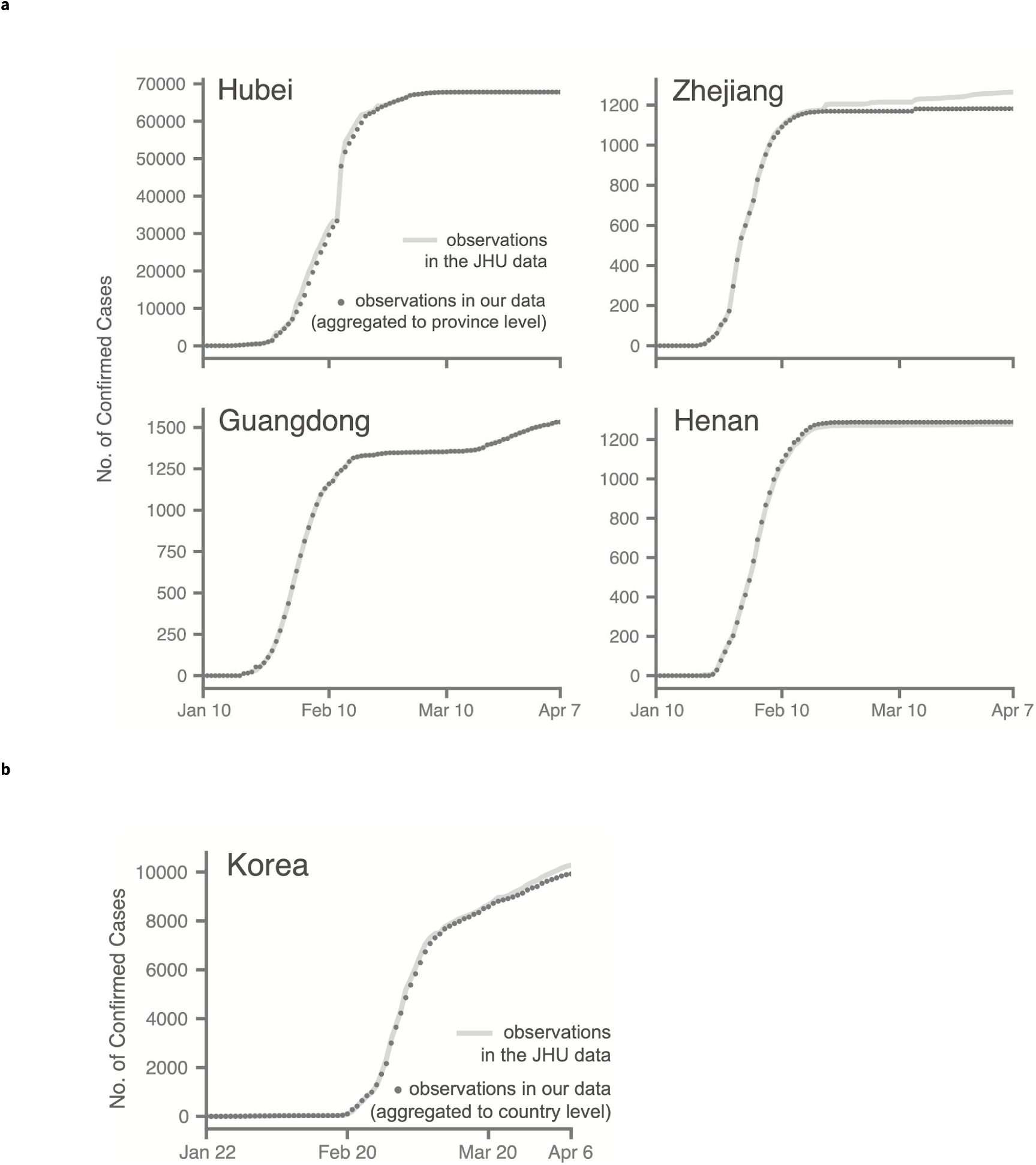
Validating disaggregated epidemiological against aggregated data from the Johns Hopkins Center for Systems Science and Engineering. Comparison of cumulative confirmed cases from a subset of regions in our collated epidemiological dataset to the same statistics from the 2019 Novel Coronavirus COVID-19 (2019-nCoV) Data Repository by the Johns Hopkins Center for Systems Science and Engineering (JHU CSSE).^1^ We conduct this comparison for Chinese provinces and South Korea, where the data we collect are from local administrative units that are more spatially granular than the data in the JHU CSSE database. **a**, In China, we aggregate our city-level data to the province level, and **b**, in Korea we aggregate province-level data up to the country level. Small discrepancies, especially in later periods of the outbreak, are generally due to imported cases (international or domestic) that are present in national statistics but which we do not assign to particular cities (in China) or provinces (in Korea).

**Extended Data Figure 2.**
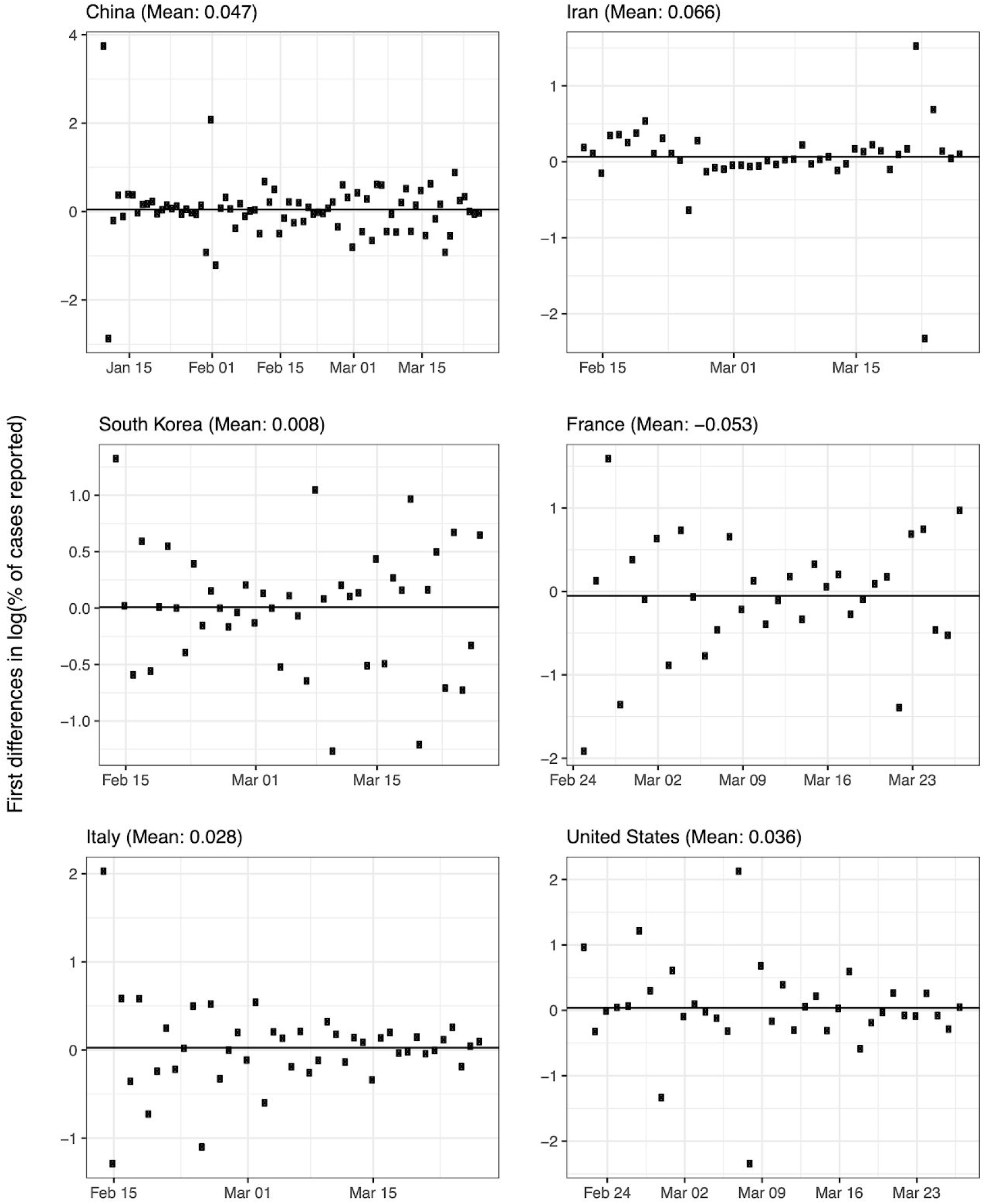
Estimated trends in case detection over time within each country. Systematic trends in case detection may potentially bias estimates of no-policy infection growth rates (see Equation 8). We estimate the potential magnitude of this bias using data from the Centre for Mathematical Modelling of Infectious Diseases.^2^ Markers indicate daily first-differences in the logarithm of the fraction of estimated symptomatic cases reported for each country over time. The average value over time (solid line and value denoted in panel title) is the average growth rate of case detection, equal to the magnitude of the potential bias. For example, in the main text we estimate that the infection growth rate in the United States is 0.29 (Figure 2A), of which growth in case detection might contribute 0.049 (this figure). Sample sizes are 75 in China, 41 in Iran, 40 in South Korea, 29 in France, 40 in Italy, and 32 in the US.

**Extended Data Figure 3.**
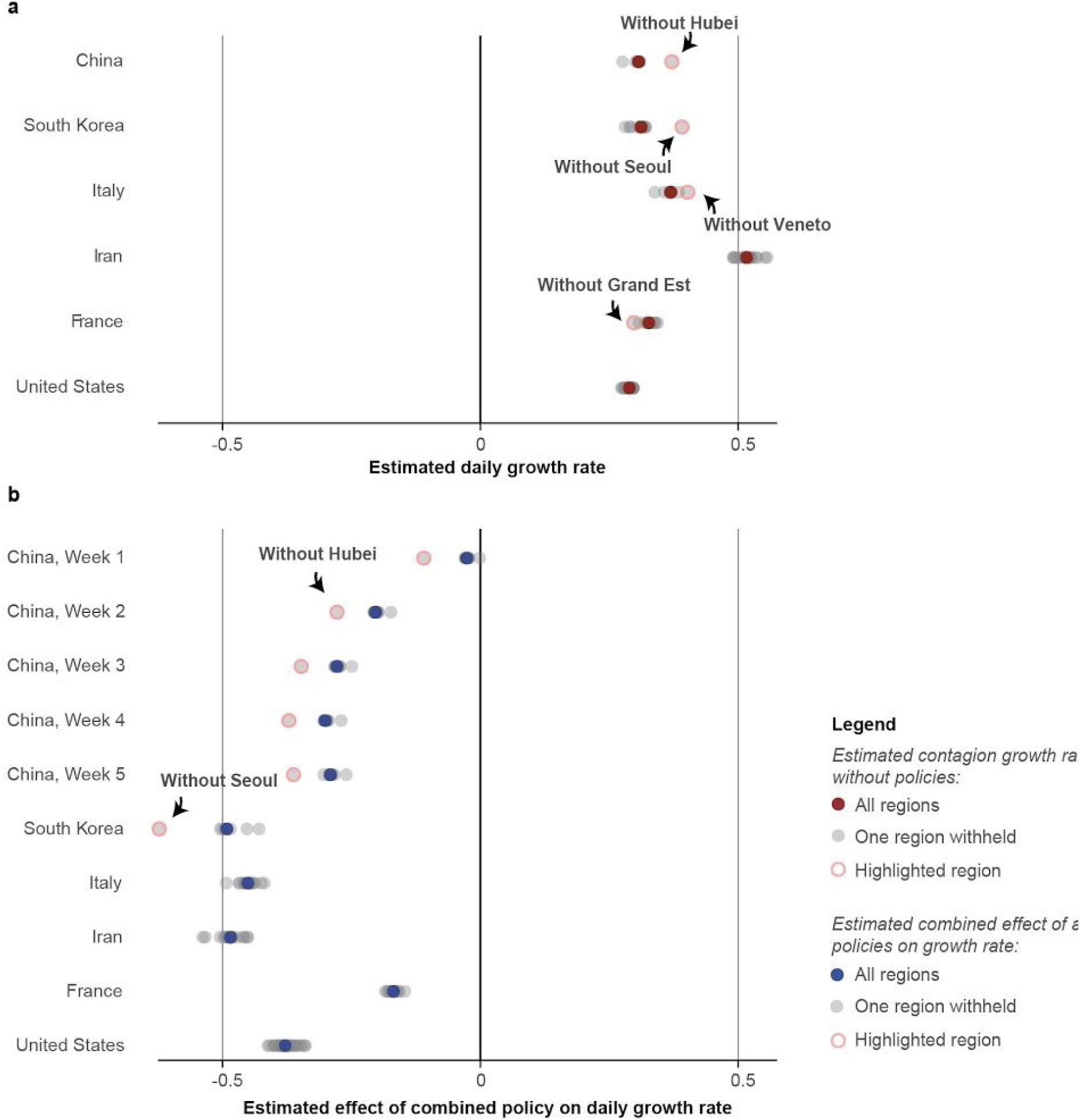
Robustness of the estimated no-policy growth rate of infections and the combined effect of policies to withholding blocks of data from entire regions. For each country, we re-estimated Eq. 7 using real data *k* times, each time withholding one of the *k* first-level administrative regions (“Adm1,” i.e. state or province) in that country. Each gray circle is either (**a**) the estimated no-policy growth rate or (**b**) the total effect of all policies combined, from one of these *k* regressions. Red and blue circles show estimates from the full sample, identical to results presented in panels A and B of Figure 2, respectively. For each country panel, if a single region is influential, the estimated value when it is withheld from the sample will appear as an outlier. Some regions that appear influential are highlighted with an open pink circle. As in Figure 2B of the main text, we estimate a distributed lag model for China and display each of the estimated weekly lag effects (red circle is the same “without Hubei” sample for lags). The full sample includes 3, 684 observations in China, 595 in South Korea, 2, 898 in Italy, 548 in Iran, 270 in France, and 1, 238 in the US.

**Extended Data Figure 4.**
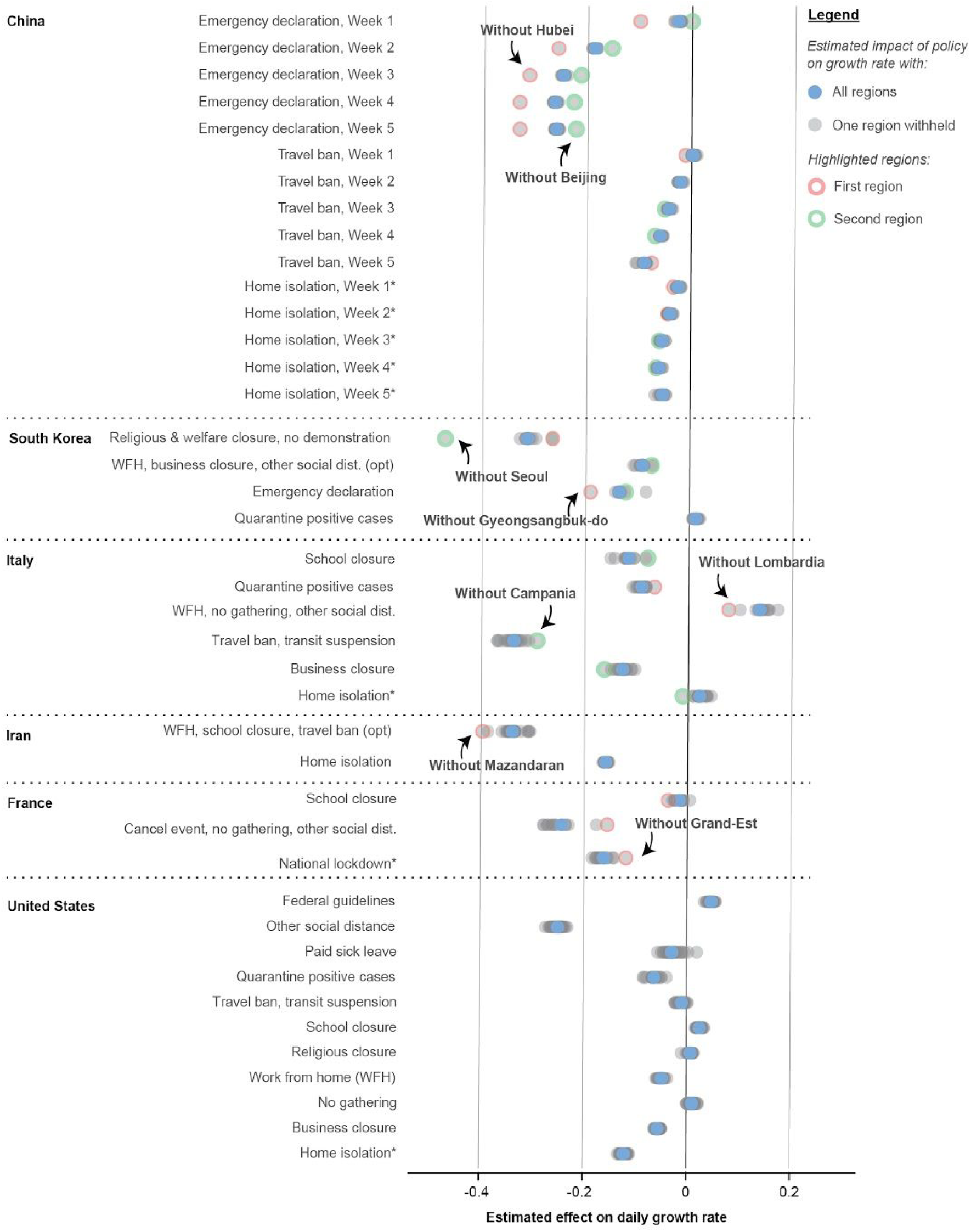
Robustness of the estimated effects of individual policies to withholding blocks of data from entire regions. Same as Extended Data Figure 3, but for individual policies (analogous to Figure 2C in the main text). In cases where two regions are influential, a second region is highlighted with an open green circle. The full sample includes 3, 669 observations in China, 595 in South Korea, 2, 898 in Italy, 548 in Iran, 270 in France, and 1, 238 in the US.

**Extended Data Figure 5.**
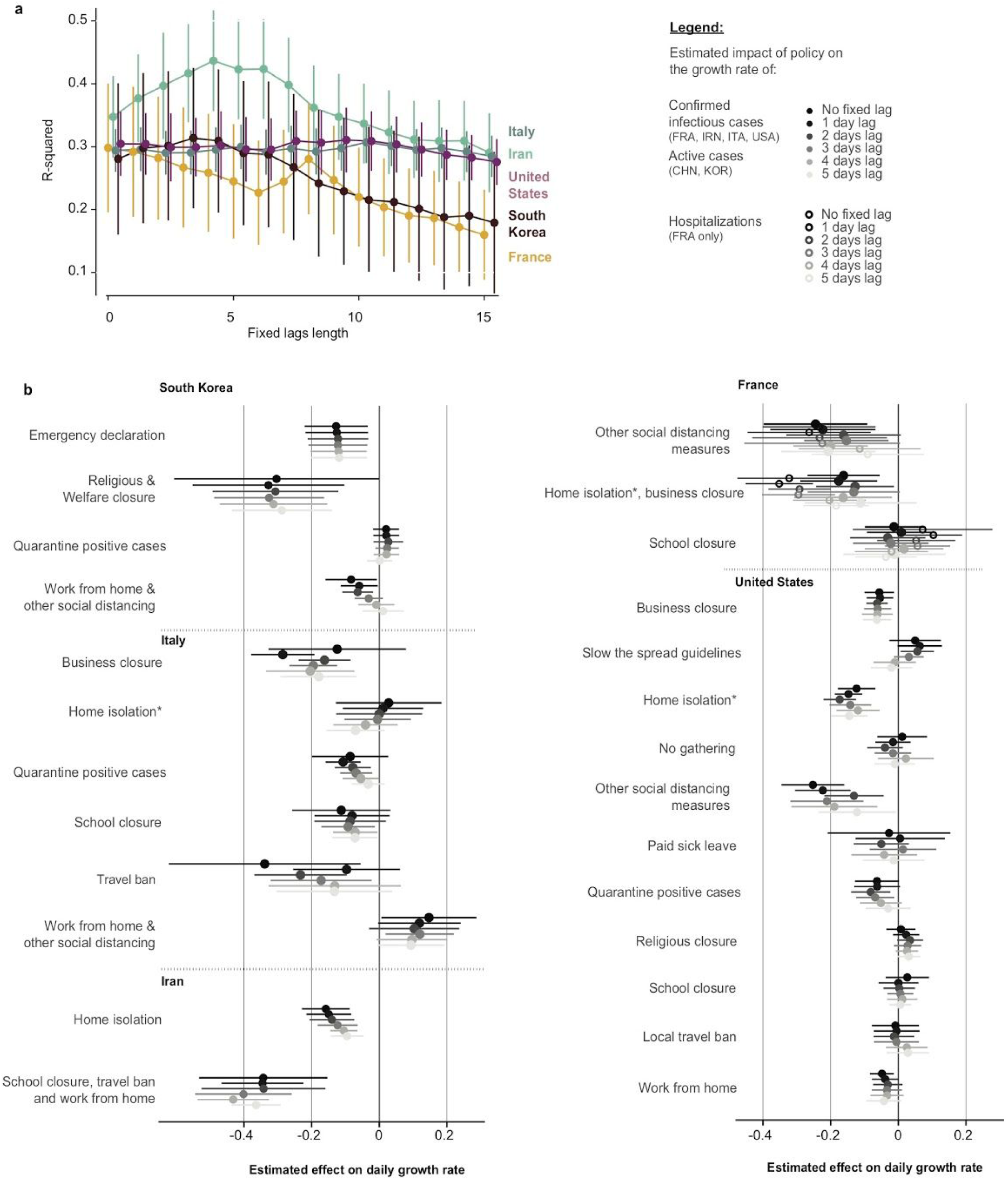
Evidence supporting models where policies affect infection growth rates in the days following deployment. Existing evidence has not demonstrated whether policies should affect infection growth rates in the days immediately following deployment. It is therefore not clear *ex ante* whether the policy variables in Eq. 7 should be encoded as “on” immediately following a policy deployment. We estimate “fixed-lag” models in which a fixed delay between a policy’s deployment and its effect is assumed (see Supplementary Methods Section 3). If a delay model is more consistent with real world infection dynamics, these fixed lag models should recover larger estimates for the impact of policies and exhibit better model fit. **a**, R-squared values associated with fixed-lag lengths varying from zero to fifteen days. Center values represent the R squared value in our sample, whiskers are 95% CI computed through resampling with replacement. In-sample fit generally declines or remains unchanged if policies are assumed to have a delay longer than four days. **b**, Estimated effects for no lag (the model reported in the main text) and for fixed-lags between one and five days. Center values represent the point estimate, error bars are 95% CI. Estimates generally are unchanged or shrink towards zero (e.g. *Home isolation* in Iran), consistent with mis-coding of post-policy days as no-policy days. The sample size is 595 in South Korea, 2, 898 in Italy, 548 in Iran, 270 in France, and 1, 238 in the US.

**Extended Data Figure 6.**
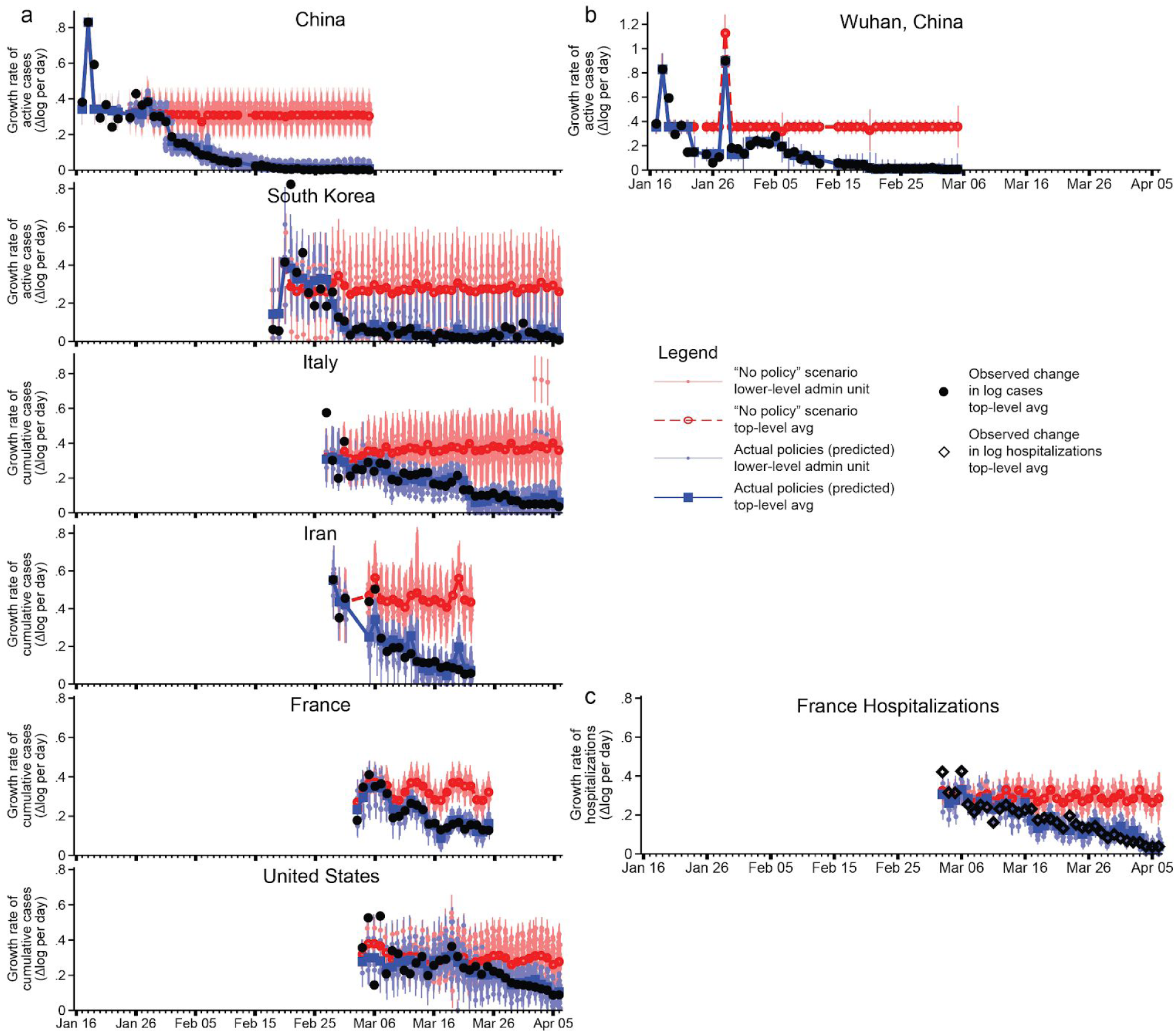
Estimated infection or hospitalization growth rates with actual anti-contagion policies and in a “no policy” counterfactual scenario. **a**, The estimated daily growth rates of active (China, South Korea) or cumulative (all others) infections based on the observed timing of all policy deployments within each subnational unit (blue) and in a scenario where no policies were deployed (red). Identical to Figure 3 in the main text, but using an alternative disaggregated encoding of policies that does not group any policies into policy packages. The sample size is 3, 669 in China, 595 in South Korea, 2, 898 in Italy, 548 in Iran, 270 in France, and 1, 238 in the US. **b**, Same as Figure 3 in the main text, but Eq. 7 is implemented for a single example administrative unit, Wuhan, China. The sample size is 46 observations. **c**, Same as Figure 3 in the main text, but using hospitalization data from France rather than cumulative cases (the French government stopped reporting cumulative cases after March 25, 2020). The sample size is 424 observations. For all panels, the difference between the with- and no-policy predictions is our estimated effect of actual anti-contagion policies on the growth rate of infections (or hospitalizations). The markers are daily estimates for each subnational administrative unit (vertical lines are 95% confidence intervals). Black circles are observed changes in log(*infections*) (or diamonds for log(*hospitalizations*)), averaged across observed administrative units.

**Extended Data Figure 7.**
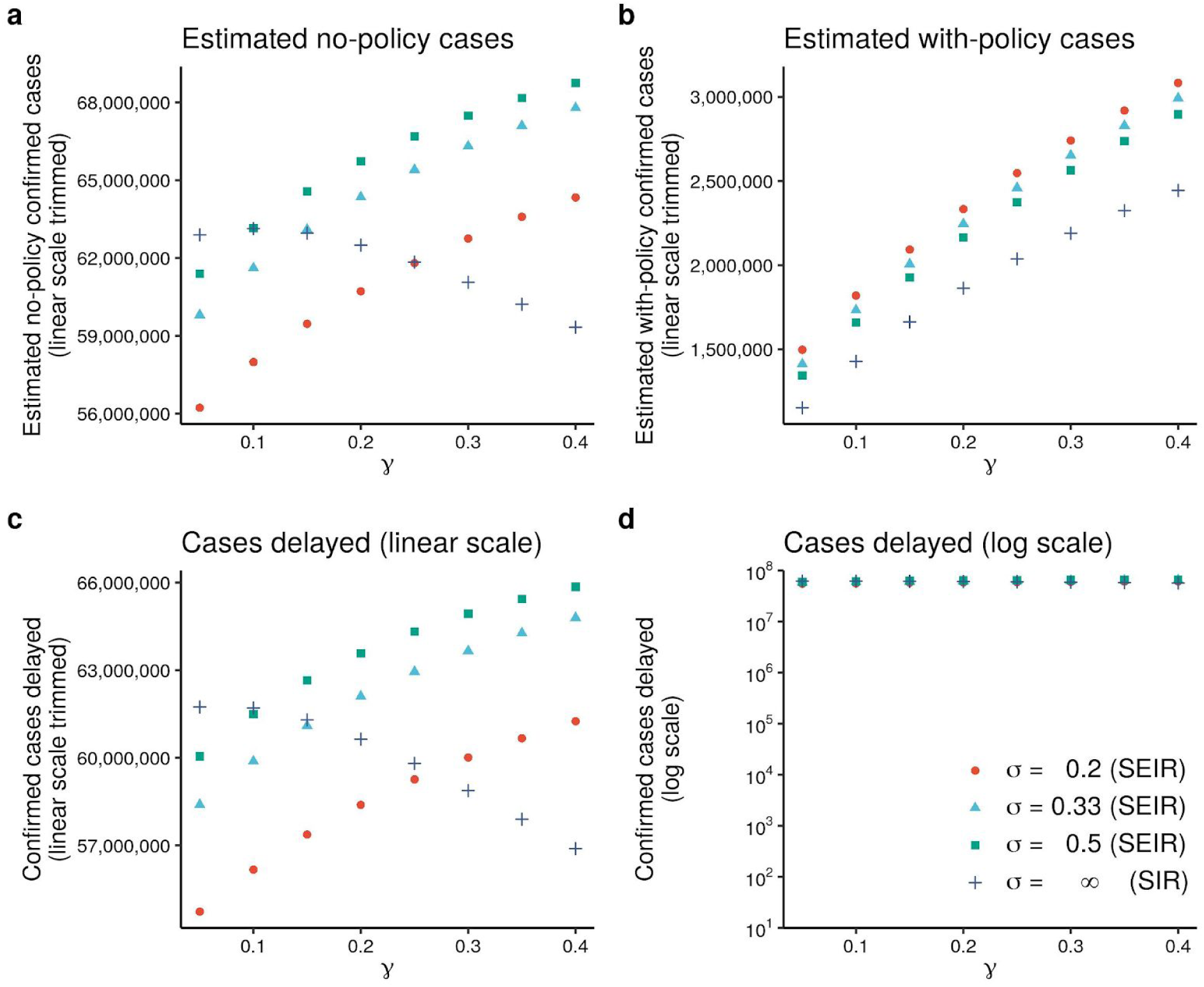
Sensitivity of estimated averted/delayed infections to the choice of γ and σ in an SIR/SEIR framework. This figure displays the sensitivity of total averted/delayed cases presented in Figure 4 of the main text to alternative modeling assumptions. We compute total cases across the respective final days in our samples for the six countries presented in our analysis. The figure displays how these totals vary with eight values of *γ* (0.05-0.4) and four values of *σ* (0.2, 0.33, 0.5, ∞), where the final value of *σ* (∞) corresponds to the SIR model. **a**, The simulated total number of infections under no policy. **b**, Same, but using actual policies. **c**, The difference between (**a**) and (**b**), which is the total number of averted/delayed infections. **d**, Same as (**c**), but on a logarithmic scale similar to Figure 4 in the main text (**a** - **c** are on a linear scale, trimmed to show details). Figure 4 in the main text uses *γ* **=** 0.079, which we calculate using empirical recovery/death rates in countries where we observe them (China and South Korea, see Methods). If we assume a 14-day delay between infected individuals becoming non-infectious and being reported as “recovered” in the data, we would calculate *γ* **=** 0.18. Figure 4 in the main text assumes *σ* = ∞.

**Extended Data Figure 8.**
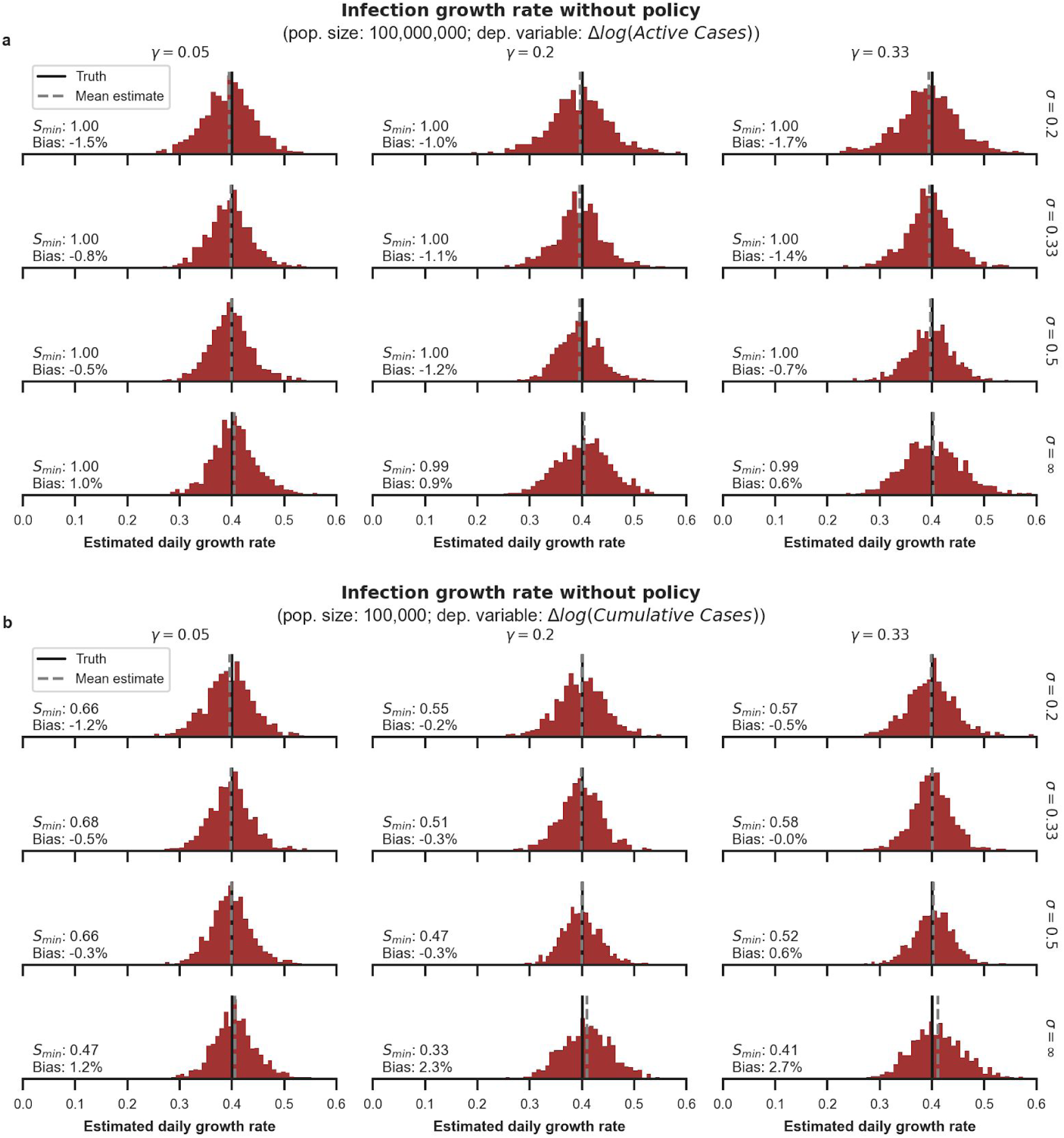
Simulating reduced-form estimates for the no-policy growth rate of infections for different population regimes and disease dynamics. We examine the performance of reduced-form econometric estimators through simulations in which different underlying disease dynamics are assumed (see Supplementary Information Section 3). Each histogram shows the distribution of econometrically estimated values across 1, 000 simulated outbreaks. Estimates are for the no-policy infection growth rate (analogous to Figure 2a) when three different policies are deployed at random moments in time. The black line shows the correct value imposed on the simulation and the red histogram shows the distribution of estimates using the regression in Eq. 7, applied to data output from the simulation. The grey dashed line shows the mean of this distribution. The 12 subpanels describe the results when various values are assigned to the mean infectious period (*γ^−^*^1^) and mean latency period (*σ^−^*^1^) of the disease. “*σ = ∞”* is equivalent to SIR disease dynamics. In each panel, *S_min_* is the minimum susceptible fraction observed across all 1, 000 45-day simulations shown in each panel. In the real datasets used in the main text, after correcting for country-specific under-reporting, *S_min_* across all units analyzed is 0.72 and 95% of the analyzed units finish with *S_min_* > 0.91. “Bias” refers to the distance between the dashed grey and black line as a percentage of the true value. **a**, Simulations in near-ideal data conditions in which we observe active infections within a large population (such that the susceptible fraction of the population remains high during the sample period, similar to those in our data for Chongqing, China). **b**, Simulations in a non-ideal data scenario where we are only able to observe cumulative infections in a small population (similar to those in our sample for Cremona, Italy).

**Extended Data Figure 9.**
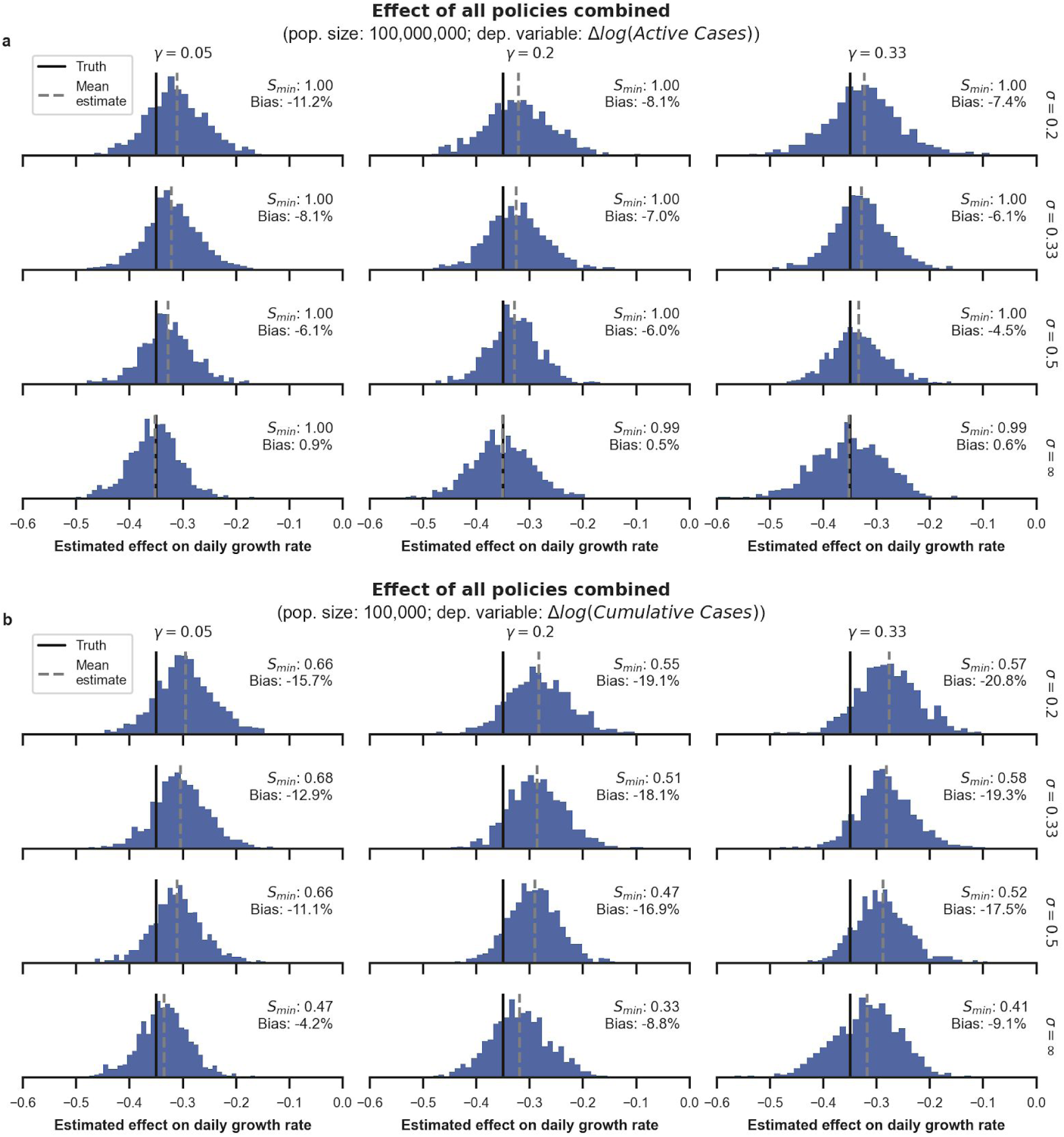
Simulating reduced form estimates for anti-contagion policy effects for different population regimes and assumed disease dynamics. Same as Extended Data Figure 8, but estimates are for the combined effect of three different policies (analogous to Figure 2b) that are deployed at random moments in time.

**Extended Data Figure 10.**
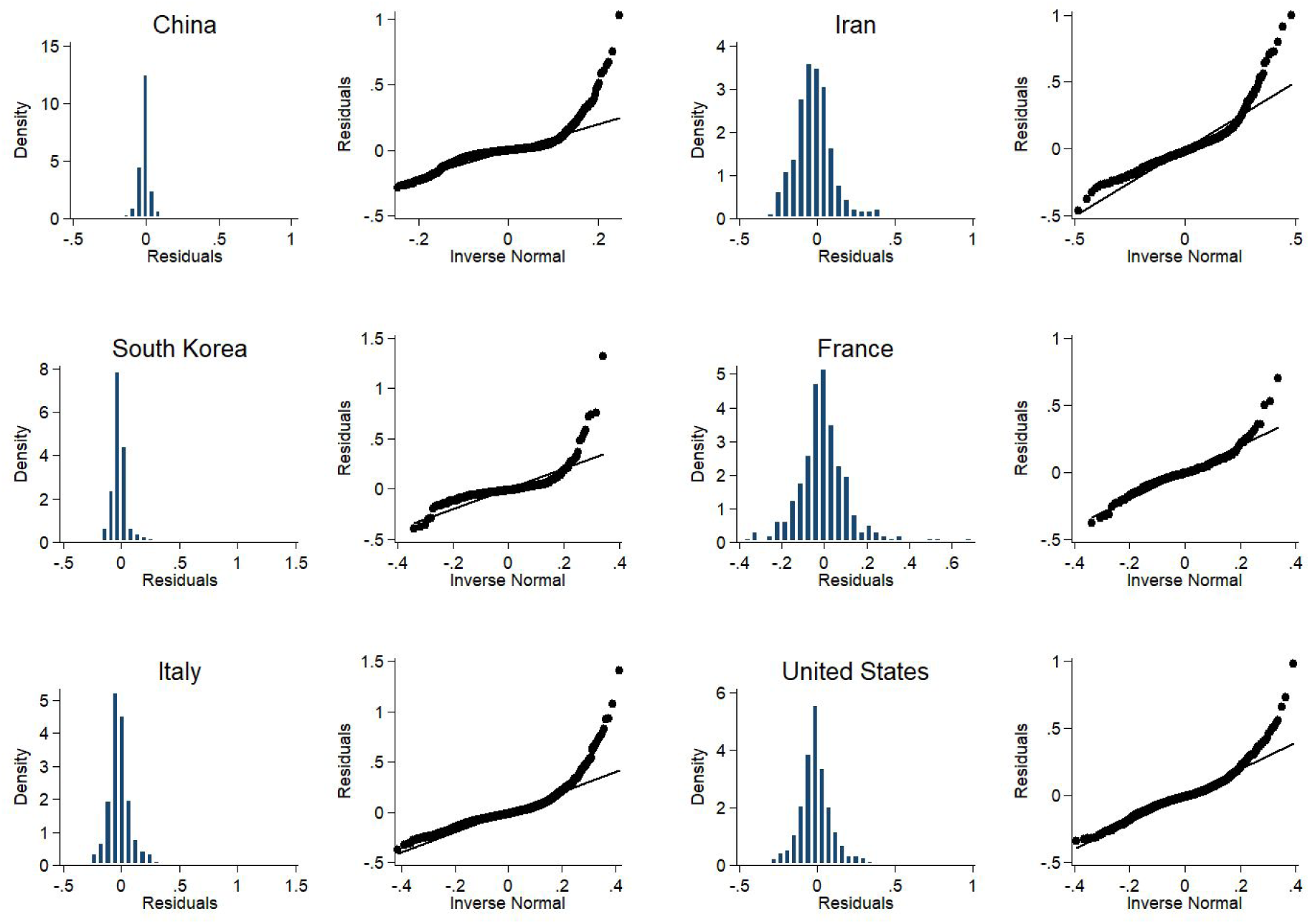
Regression residuals for the growth rates of COVID-19 by country. These plots show the estimated residuals from Equation 7 for each country-specific econometric model. Histograms (left) show the estimated unconditional probability density function. Quantile plots (right) show quantiles of the cumulative density function (y-axis) plotted against the same quantiles for a Normal Distribution. For additional details, see Fig. 3 and the Econometric Analysis section of Methods.

## Supplementary Information for “The Effect of Large-Scale Anti-Contagion Policies on the COVID-19 Pandemic”

The following sections provide additional information to support the article and to document our data collection and processing procedures.

All data and code are available at https://github.com/bolliger32/gpl-covid.

Additional resources and updates are available at www.globalpolicy.science/covid19.

For repository related questions, please contact.

For data related questions, please contact.

This document last updated: May 16, 2020. Data last updated: April 6, 2020.

The Supplementary Information contains three sections: Supplementary Notes, Supplementary Methods, and Supplementary Tables.

The Supplementary Notes section details the basic accounting of policy deployment decisions in each of the six countries we analyze, and describes the data acquisition and processing procedure for the epidemiological and policy data used in this paper. The sources for both types of data come from a variety of in-country data sources, which include government public health websites, regional newspaper articles, and Wikipedia crowd-sourced information. We have supplemented this data with international data compilations. A list of the epidemiological and policy data compiled for this analysis can be found here.

The Supplementary Methods section describes sensitivity analyses and simulations performed to verify the robustness of our model, including: the sensitivity of our counterfactual projections to varying epidemiological parameters; and the sensitivity of our regression estimates to alternative lag structures, withholding of data, and differing policy groupings.

The Supplementary Tables section contains tables detailing:

1. Number of unique anti-contagion policies tabulated by administrative division of each country.
2. Epidemiological data in Wuhan prior to policy intervention, and estimates of the initial infection growth rate and case doubling times.
3. Full regression results from our main regression estimating the effect of policy on growth rates.
4. Estimates of the effect of policies on infection growth rate using a more disaggregated grouping of policies.
5. Estimates of the effect of policies on infection growth rate assuming a variety of lag structures.
6. Estimates of the cumulative number of confirmed cases averted assuming a variety of country-specific underreporting ratios.

## Supplementary Notes

### Chronology of Policy Implementation

In the main article, we analyze how infection growth rates change when anti-contagion policies are deployed. In order for these estimated associations to be interpreted as plausibly causal, it must be the case that changes in policy are not correlated with changes in infection growth rates that would have occured in the absence of policy actions. For locations and periods where policy^1^ was actually deployed, it is not possible to directly observe how growth rates would have changed in the absence of policy actions.^2^ In general, it is well-established that in the absence of policy, early rates of infection within a susceptible population follow almost-perfect exponential growth.^3, 4, 5, 6, 7, 8, 9, 10^ This fact would suggest that in the absence of policy, infection growth rates would be constant^11, 12^ and, therefore, changes in the infection growth rate could not be correlated with the timing of policy deployments (since correlation with a constant variable is zero). However, it is nonetheless also worthwhile to consider how the timing of policy actions were determined in practice. In this section, we provide a basic accounting of policy deployment decisions in each of the six countries we analyze, focusing on the timing of major events and reported motivations for notable policy deployments. These accounts are intended to familiarize readers with the general decision-making context of each country, although they are not exhaustive and are not intended to capture the full complexity of decision-making in the ongoing pandemic.

As stated in the main text, policies were generally not initially deployed with any reference to observed or anticipated natural changes in growth rates. In general, policies tended to be deployed in response to high total numbers of cases or to outbreaks in other regions, after long delays due to political constraints, and often with timing that coincided with arbitrary events, like the start and end of the week, or holidays. For example, epidemic planning in France explicitly tied policy actions to case-counts in specific regions, and epidemiological guidance in the US explicitly recommended policy actions depending on when thresholds in total case counts were passed. In South Korea, policy actions across the country were initially triggered by an idiosyncratic outbreak in a Shincheonji Church in Daegu, and in Iran they were triggered by an outbreak in Qom coinciding with a religious pilgrimage. In China and Italy, nation-wide policy actions were deployed relatively swiftly by the central government in response to a regional outbreak, with limited variation across locations that could be correlated with changes in local growth rates. In China, Italy, and the US, policies tended to be deployed during the weekend or on a Monday or Friday, with the preferred day of the week varying across these countries.

### China

In China, most anti-contagion policies were deployed in a centralized manner. On January 27, 2020, towards the beginning of the outbreak in Wuhan, the central government extended the Chinese New Year national holiday that originally ended on January 30 to February 2, 2020, preventing all schools and businesses across the country from resuming operations. The holiday was later further extended within various provinces to end in mid February.

The main policy instruments that cities deployed were: level 1 emergency declaration, travel bans, and home isolation. The timing of these policies varied across cities, but was still strongly influenced by provincial government mandates. Indeed, cities routinely cite documents from upper-level governments when issuing statements or policies. From January 23 to January^13^ 29, 2020, all provincial governments declared Level 1 emergency responses.^14^ The declaration often included the closing of entertainment venues, banning of public gatherings, and extensive temperature monitoring at airports, railway stations and highway checkpoints.^15^ In addition to these measures, cities started to implement travel bans, limiting travelling in or out of the cities, as well as home isolation policies, which limited residents from leaving their home for non-essential activities. The first travel ban was enacted in Wuhan, on January 23, 2020, with all the cities in Hubei province adopting the same policy shortly afterwards.

Many Chinese cities appeared to enact policies in response to outbreaks in other localities or other political considerations that did not simply reflect the spread of infections across regions. For example, Tian et al. (2020)^16^ examined the policy responses in China during the first 50 days of the pandemic and found that about half of the cities that implemented transmission control measures did so before the first local case was reported (see Figure S2 in Tian et al. (2020)). In another example, many cities in Hubei did not implement a home isolation policy until February 10, 2020, lagging behind many other cities in other provinces, even though the count of cases was highest in Hubei. A disproportionate fraction of policies were deployed on Fridays in China (36.2%).

### South Korea

South Korea confirmed its first imported coronavirus case from Wuhan, China on January 20, 2020.^17^ By January 29, 2020, there were four confirmed cases, all originating from Wuhan. In response, the Korean Center for Disease Control began systematically screening all travelers arriving from China.^18^

On February 18, 2020, South Korea confirmed its 31st case in Daegu Metropolitan City, an individual later identified as a member of the Shincheonji Church.^19^ Over the next two days, the number of cases more than doubled, almost half of them linked to the Shincheonji Church.^20^ While the outbreak was localized in Daegu, both Daegu and Gyeongsangbuk-do (the province that surrounds Daegu) immediately shut down all Shincheonji-affiliated places of worship. The outbreak in Daegu triggered most other provinces to follow suit by closing all religious facilities associated with Shincheonji. This included provinces with significantly lower infection rates at that point in time, such as Jeollabuk-do, Jeollanam-do, Gwangju and even Jeju, an island-province far removed from the epicenter of the outbreak.^21^

The Shincheonji outbreak prompted the central government to activate the highest health alert level on February 23, 2020.^22^ This was a turning point in the coronavirus response across the country, ^23, 24, 25, 26, 27^ with South Korean President Moon Jae-in declaring,

> *“The [coronavirus] situation [in the country] is entirely different after the Shincheonji outbreak…*.
>
> *The central government, local governments, health officials and medical personnel and the entire people must wage an all-out, concerted response to the [COVID-19] problem.”*

By February 26, 2020, there were over 1, 100 confirmed cases, approximately half of which were attributable to the Shincheonji Church.^28^ The identification of the superspreader linked to the Shincheonji Church and the consequent virus outbreak resulted in a large number of governmental countermeasures deployed at the end of February. As the number of confirmed cases surpassed 7, 000 in Daegu and some cities in Gyeongsangbuk-do in mid-March, the South Korean government declared a state of emergency for those regions hardest hit by the virus.^29^ Throughout the country, additional policies were adopted, including strengthened travel restrictions, wider testing regimes, recommendations for social distancing, and guidelines for certain business operations.

On March 19, 2020, as the number of confirmed cases in other countries and the risk of importing the pathogen increased, the central government extended screening measures to all inbound travelers arriving from every country, ^30, 31^ requiring them to report their symptoms daily for 14 days via a self-diagnosis mobile application.^32^ As the situation in other countries worsened, the government intensified its efforts in limiting the spread of the virus from abroad. On March 22, 2020, inbound travelers from Europe were subjected to a home-quarantine for 14 days.^33^ This policy was revised on March 27, 2020 to also include all symptomatic travelers arriving from the US.^34^ Shortly after, on April 1, 2020, this policy was expanded to include inbound travelers arriving from all countries.^35^

### Italy

The first confirmed cases in Italy were imported cases detected on January 31, 2020.^36^ By mid February, the virus had spread to Lombardy, and later Veneto, in Northern Italy.^37^ The hardest hit municipalities within these regions (Bertonico, Casalpusterlengo, Castelgerundo, Castiglione d’Adda, Codogno, Fombio, Maleo, San Fiorano, Somaglia, and Terranova dei Passerini in Lombardy, and Vo’Eugane in Veneto) were the first areas in Italy to be placed under lockdown on February 22, 2020.

The Italian government made the decision at the national level to close schools across the country on March 5, 2020.^40^ On March 8, 2020, Prime Minister Giuseppe Conte placed Lombardy and 14 other northern provinces under lockdown, and placed restrictions on the rest of the country (such as suspending any public or private gatherings and other social distancing measures).^41^ He then placed the entire country under lockdown on March 10, 2020, ^42^ with increasing restrictions on commercial activities passed on March 11, 2020.^43^ Lastly on March 22, 2020, additional commercial activities and industries were closed, and people’s ability to travel across the country were further curtailed.^44, 45^

Throughout this period, Italian policymakers weighed implementing restrictive anti-contagion policies against maintaining individual freedoms, and were delayed in reaching consensus on when to implement policies due to coordinating across the different branches of government.^46^

A majority (62.7%) of all policies in Italy were deployed on a Sunday.

### Iran

The first major outbreak in Iran was connected to a major Shia pilgrimage in the city of Qom that brought travelers from Iran and throughout the Middle East to visit the Fatima Masumeh shrine, which often involves kissing or touching the shrine.^47^ The virus, initially centered in Qom and neighboring Tehran, spread rapidly throughout the country. Early in the pandemic, government communication and policy responses appeared mixed.^48^ It has been reported that, as the virus spread, factions within the government competed for control over the nation’s coronavirus response, leading to implementation of policy that was not strongly coordinated.

Throughout this period, the Iranian government increased the stringency of its response as the count of confirmed cases increased. The first two cases and deaths were reported simultaneously on February 19, 2020. The next day, school closures were announced in the province of Qom and travel in the region was discouraged. On February 22, 2020, the government closed schools in 14 provinces and closed down major gathering sites such as football matches and theaters. By March 5, 2020 schools were closed nationwide and government employees were required to work from home. Home isolation was overseen by the military on March 13, 2020, described by The Guardian as a “near-curfew [following] growing exasperation among MPs that calls for Iranian citizens to stay at home had been widely ignored, as people continued to travel before the Nowruz New Year holidays.”^50^ Military implementation of home isolation was reportedly hindered due in part to pushback from the civilian government.^49^

### France

The ORSAN (*organisation de la réponse du système de santé en situations sanitaires exceptionnelles)* plan, ^51^ originally designed to deal with the 2009 H1N1 epidemic, is a predefined, structured emergency plan that guides epidemic responses in France. The ORSAN plan, which is implemented by President Emmanuel Macron and Prime Minister Édouard Philippe based on the advice of the *Conseil scientifique Covid-19*, divides the epidemic outbreak into three distinct phases (“*stades”)* accompanied by different anti-contagion policies:^52^

*Stade 1:* The phase when the virus has not prevalently spread within the French population. Policies at this point aim to track infected individuals and identify the source of contagion in order to prevent future clusters. *Stade 1* was declared on February 25, 2020. *Stade 2:* The phase when the virus has spread to a limited number of regions within the country. During this phase, policies deployed are designed to slow the propagation of the virus within these regions. For instance, understock, localities may close schools or limit access to elderly care facilities on a case-by-case basis. *Stade2 was* declared on February 29, 2020 when the cumulative number of cases within the country reached 100 cases, and the national government prohibited large gatherings of over 5, 000 people. On March 9, 2020, this national policy was subsequently changed to limiting gatherings to no more than 1, 000 people. *Stade 3:* The phase when every region of the country is affected by the virus. Policies are intended to mitigate the impact of the epidemic. *Stade 3* officially started on March 14, 2020 with a speech from President Emmanuel Macron. He announced a nationwide school closure starting the following Monday. The next day the government decided to close all non-essential business and the national lockdown began on March 17, 2020.

The central government relied heavily on the *Conseil scientifique Covid-19* for the management of the crisis. The *Conseil scientifique Covid-19’s* analysis was largely based on epidemiological models inspired by the Imperial College model (Ferguson et al, 2006; Luca et al, 2018; Ferguson et al, 2005).^53^ According to the *Conseil scientifique Covid-19*’s first report, the goal of anti-contagion measures was to limit the number of individuals who would require hospitalization.^54^

### United States

Although the first confirmed COVID-19 case in the US was documented on January 20, 2020, ^55^ many jurisdictions across the country did not implement mandatory policies at the county or state level until early-to mid-March, with some states delaying more restrictive policy action until even later. At the national level, both policy action and public engagement surrounding COVID-19 moved slowly. On January 24, 2020, the head of the National Institute of Allergy and Infectious Diseases, Anthony Fauci, commented that:

> “*We don’t want the American public to be worried about this because their risk is low.”*^56^

On January 28, 2020, Alex Azar, the secretary of Health and Human Services gave a Press Briefing in which the tone remained largely the same:

> “*As of today, the CDC has reported 5 cases of the novel coronavirus infection here in the United States. China has now reported more than 4, 500 cases. Americans should know that this is a potentially very serious public health threat, but, at this point, Americans should not worry for their own safety.”*^57^

However, on January 31, 2020, Azar declared a public health emergency for the US.^58^ The first national policy action, implemented on February 2, 2020, suspended entry into the United States of foreign nationals who had visited China in the previous 14 days and mandated quarantine for Americans who had recently visited hard hit regions of China.^59^ The federal government followed this Presidential Proclamation with a similar ban on February 29, 2020, prohibiting entry for non-U.S. citizens who had been in Iran.^60^ On March 11, 2020, the federal administration further suspended arrivals by halting travel from the Schengen Area in Europe.^61^

On March 16, 2020 the White House announced “15 Days to Slow the Spread,” a set of social distancing guidelines that urged Americans to work from home, engage in remote schooling, avoid unnecessary shopping trips, refrain from visiting restaurants and bars, and not gather in groups of more than 10 people.^62^ It was widely reported that these federal guidelines drew on a report by the Imperial College COVID-19 Response Team (Ferguson et al, 2020) that recommended non-pharmaceutical interventions to slow the spread of the virus.^63^ This report assumed that infection was “seeded in each country at an exponentially growing rate” (page 4) and provided guidance to policymakers that “suppression policies are best triggered early in the epidemic, with a cumulative total of 200 ICU cases per week being the latest point at which policies can be triggered” and still keep peak ICU demand below surge limits.^64^

Besides the federal travel bans and social distancing guidelines, the majority of US anti-contagion policies were enacted in a decentralized manner by state governors. Washington State, the first state to suffer an outbreak, declared a state of emergency on February 29, 2020 in response to the first in-state COVID-19 death.^65^ A few weeks later, Washington Governor Jay Inslee passed “the immediate two-week closure of all restaurants, bars, and entertainment and recreational facilities, as well as additional limits on large gatherings” starting on March 16, 2020, where his “announcement comes after the recent spike in numbers of COVID-19 cases in the state and across the country.”^66^ A week later, on March 24, 2020, Governor Inslee announced a state-wide shelter-in-place order. In his announcement, he wrote: “We have now confirmed that more than 2, 000 Washingtonians have contracted the virus. There are likely thousands more that have not yet been diagnosed. COVID-19 has taken more than 100 lives in our state, a number that will also continue to rise…So, tonight, I am issuing a ‘Stay at Home’ order to fight this virus…This includes a ban on all gatherings, and closures of many businesses.”^67^

In California, another state with early confirmed cases of COVID-19, the Health and Human Services Agency in the California Department of Public Health issued on March 7, 2020 “statewide guidance to help both school and public health officials inform their decision making” on whether to close schools and school districts.^68^ They included different scenarios each with a separate course of action, where scenarios changed based on whether there were two or more confirmed cases in the community; one or more confirmed case(s) by a student or teacher at the school; or if multiple schools within the school district had a student or teacher test positive. The recommendations for some of these scenarios included closing the school or the entire school district. On March 19, 2020, Newsom announced the state-wide shelter in place order. The governor said his decision to deploy the policy came after epidemiological models that projected the spread of COVID-19 in California.^69^

New York State, which would become the center of the US outbreak, confirmed its first case on March 1, 2020. Governor Andrew Cuomo responded by saying that the general risk to New Yorkers “remains low” and that “we are fully coordinated, and we are fully mobilized, and we are fully prepared to deal with the situation as it develops.”^70^ On March 9, 2020, during an interview with Katy Tur of MSNBC, when asked “What about schools? Are you considering closing them? What will it take to close them?” Cuomo responded:

> *“We close them on a case-by-case basis…You have higher numbers in certain areas [of the state], lower in others. If the numbers are low, God bless. If the numbers are high, take action. As I mentioned before where we have a cluster in Westchester that has more cases than New York City. So how do you handle that hotbed, that hot spot, as they call it? Closing schools, closing gatherings, etcetera, extraordinary efforts where you have higher density of cases.”*

Only a few days later, on March 16, 2020, New York and neighboring states Connecticut and New Jersey, announced that they would limit social and recreational gathering to 50 people^71^ per national CDC recommendations issued the day before.^72^ The three states also coordinated the implementation of business closures (including restaurants, bars, movie theaters and casinos) effective on March 16, 2020.^73^ The decision to implement these uniform restrictions all at once across the three neighboring states was to “prevent ‘state shopping’ where residents of one state travel to another and vice versa.”^74^

In other states that have subsequently been affected by the virus, policies to close businesses and schools, reduce gatherings and cancel events, and reduce travel have been implemented. In some cases, the timing of these policies was determined by a holiday or in response to a high number of confirmed cases in-state. For example Ohio (on March 15, 2020), ^75^ Michigan (on March 16, 2020), ^76^ Illinois (on March 16, 2020)^77^ and Massachusetts (on March 17, 2020)^78^ responded preemptively to the St. Patrick’s Day holiday (on March 17, 2020) by ordering the closures of bars and restaurants. In Georgia, the closure of public K-12 schools starting on March 18, 2020, the limiting of large gatherings and closing of bars on March 23, 2020, implementing a statewide shelter in place on April 2, 2020, were deployed in response to “the number of COVID-19 cases in Georgia [which has] continue[d] to rise,” as stated in the Executive Orders issued by Governor Bill Kemp.^79^ On March 18, 2020, Kentucky closed all businesses that interfaced with the public, such as recreational facilities, hair salons, and concert venues, and recommended against visiting senior, psychiatric, or acute care centers.^80^

Policies across the US were deployed on all days of the week, but a disproportionate number were deployed on Mondays (25.0%) and Fridays (23.8%) relative to other days of the week (10.2% on average).

## Epidemiological Data

The epidemiological datasets and sources used in this paper are described below. The main health variables of interest are:

1. *cum_confirmed_cases:* The total number of confirmed positive cases in the administrative area since the first confirmed case.
2. *cum_deaths:* The total number of individuals that have died from COVID-19.
3. *cum_recoveries:* The total number of individuals that have recovered from COVID-19.
4. *cum_hospitalized:* The total number of hospitalized individuals.
5. *cum_hospitalized_symptom:* The total number of symptomatic hospitalized individuals.
6. *cum_intensive_care*: The total number of individuals that have received intensive care.
7. *cum_home_confinement:* The total number of individuals that have been self-quarantined in their homes as a result of a positive test.
8. *active_cases:* The number of individuals who currently still test positive on the date of the observation.
9. *active_cases_new:* The number of new active cases since the previous date.
10. *cum_tests:* The total number of tests (includes both positive and negative results) conducted in an administrative unit.

Additional metadata accompanying the health outcome variables:

1. *date:* The date of observation.
2. *adm0_name:* The IS03 (country) code to which this observation belongs.
3. *adml_name:* The name of the Adm1 region (typically state or province) to which this observation belongs.
4. *adm2_name\* If the dataset contains observations at the Adm2 level, then this is the name of the Adm2 region to which this observation belongs (e.g. counties in the United States).
5. *adm[l, 2]_id:* Any alphanumeric ID scheme to identify different administrative units (e.g. FIPS code in the United States).
6. *lat:* The latitude of the centroid of the administrative unit.
7. *lon:* The longitude of the centroid of the administrative unit.
8. *policies_enacted:* The number of active policies that are in place for the administrative unit as of that date. This variable is not population weighted.
9. *testing_regime:* A categorical variable used to identify when an administrative region changed their COVID-19 testing regime. This is zero-indexed, with the ordering only indicating chronological progression (there is no external meaning to Regime 2 vs. Regime 1 vs. Regime 0, and there is no consistency enforced for coding across countries). For example, if China changes their testing regime twice, all observations prior to the first regime change would be coded *testing_regime=0*, all observations in between the two changes would be coded *testing_regime=l*, and all observations after the second change would be coded *testing_regime=2*.
10. *population:* The population of the administrative unit.
11. *popjsjmputed:* A binary variable equal to 1 if the population is imputed, and 0 otherwise. Used for imputing the population of some cities in China.

### Data Imputation

In certain instances where health outcome observations are missing or suffer from data quality issues, we have imputed to fill in the missing values. Imputed health outcome variables are denoted by *[health_outcome]_imputed*. We do not use any imputed health data in our analyses; it is only used for display purposes.

We impute by:

1. Taking the natural log of the non-missing observations pertaining to that health outcome variable.
2. Linearly interpolating over the missing dates for that health outcome variable.
3. Exponentiating the interpolated values back into levels and rounding to the nearest integer.

### China

We have collated a city-level time series health outcome dataset in China for 339 cities from January 10, 2020 to April 7, 2020.

For data from January 24, 2020 onwards, we relied on the public dataset Ding Xiang Yuan^81^ (DXY) that reports daily statistics across Chinese cities. Since DXY only publishes the most recent (cross-sectional) statistics (and not the historical data), we used the time series dataset scraped from DXY in an open source GitHub project.^82^ The web scraper program checks for updates at least once a day for the statistics published on DXY and records any changes in the number of cumulative confirmed cases, cumulative recoveries or cumulative deaths.

We assumed that no updates to the statistics meant there had been no new cases. We dropped a small number of cases that had been recorded but not assigned to a specific city (many of these cases are imported ones from other cities). We also dropped confirmed cases in prison populations (we assumed the spread of COVID-19 in prisons was not affected by the implementation of city-level lockdowns or travel ban policies).

For city level health outcomes prior to January 24, 2020, we manually collected official daily statistics from the central^83^ and provincial (Hubei, ^84^ Guangdong, ^85^ and Zhejiang^86^) Chinese government websites. We did not collect city level health outcomes recorded prior to January 24, 2020 in provinces that had fewer than ten confirmed cases at that date. We made this decision since our analysis dropped observations with fewer than ten cumulative confirmed cases to prevent noisy data during the early transmission phase from disproportionately biasing the estimated results.

After merging the two datasets, we conducted a few quality checks:

1. We checked that cumulative confirmed cases, cumulative recoveries, and cumulative deaths were increasing overtime. In instances when cumulative outcomes decreased overtime, we assumed that the recent numbers were more reliable, and treated the earlier number of cumulative cases as missing (this was often due to data entry errors or cases where patients that were reported to have been diagnosed with COVID-19, but were later found out to actually have tested negative). The magnitude of these errors was relatively small. We filled in any missing data with the imputation methodology described in the health data overview section.
2. We validated our city-level dataset by aggregating observations up to the provincial level and comparing the time trends from the aggregated dataset to that of the provincial dataset collated by Johns Hopkins University.^87^ We confirmed that the two datasets matched very closely (see Figure A2 Panel A).

#### Testing Regime Changes

During our sample period starting January 16, 2020, the criteria for being diagnosed with COVID-19 changed five times in China.^88^ On January 18, 2020, China began using the reverse transcription polymerase chain reaction (RT-PCR) test in addition to genome sequencing to confirm the SARS-CoV-2 infection in suspected cases.^89^ China also no longer required failure in antibiotic treatment and began considering patients who were not exposed to markets in Wuhan but had contact with symptomatic persons from Wuhan.^90^ On January 28, 2020, China began considering patients not necessarily linked to Wuhan with at least two out of the previous three required clinical manifestations.^91^ On February 13, 2020, China created a separate “clinically confirmed” case definition for the Hubei province, which counted patients who met clinical criteria through chest imaging and may not have had epidemiological links or a positive PCR test.^92^ On February 20, 2020, China reversed this decision and removed the separate “clinically confirmed” case definition for Hubei.^93^ On March 4, 2020, China expanded the possible laboratory confirmation tests for SARS-CoV-2 to include serology.^94^ We included this information in the dataset because it could have potentially changed the levels and short-term growth rates of the number of confirmed cases.

The testing regime date changes are encoded within the data cleaning script.

### South Korea

We have collated a provincial-level time series health outcome dataset in South Korea from January 20, 2020 to April 6, 2020.

Most provinces in South Korea have been publishing data on their number of confirmed coronavirus cases. Seoul, ^95^ Daegu, ^96^ Gyeongsangbuk-do, ^97^ Jeollabuk-do, ^98^ and Sejong^99^ provinces have been reporting the number of confirmed cases on a daily basis. For these provinces, we recorded this published health data.

Given that the province of Gangwon-do^100^ does not report provincial-level health data, we refer to the daily number of new cases reported by each of its counties (Chuncheon-si, ^101^ Wonju-si, ^102^ Gangneung-si, ^103^ Taebaek-si, ^104^ Sokcho-si, ^105^ and Samcheok-si^106^). As a result, we manually collected the number of new confirmed cases from each county’s webpage and aggregated the numbers to the provincial level.

The remaining provinces (Gyeonggi-do, ^107^ Incheon, ^108^ Busan, ^109^ Ulsan, ^110^ Gwangju, ^111^ Chungcheongnam-do, ^112^ Chungcheongbuk-do, ^113^ Gyeongsangnam-do, ^114^ Jeju, ^115^ and Jeollanam-do^116^) did not explicitly publish the number of cumulative confirmed cases. However, they did publish patient-level data, including the date when patients had tested positive. For these provinces, we constructed the measure of cumulative confirmed cases by counting the number of daily confirmed cases and adding it to the previous date’s total.

Most provinces did not publish the number of deaths. Instead, we checked the daily policy briefings posted on the government homepages mentioned in the footnotes and manually collected mortality data. In instances when mortality data were not found in the briefings, we obtained the mortality data from other sources, such as through social media sources (e.g. Facebook) and blogs maintained by local governments. Lastly, we supplemented these sources with mortality data reported in news articles.

#### Testing regime changes

We collected information on testing regime changes using press releases from the Korean Center for Disease Control and Prevention (KCDC). In the press release menu, the KCDC uploaded daily briefing announcements which contained information on testing criteria and changes to its testing regime.^117^ Initially, the South Korean government only tested people who: 1) demonstrated respiratory symptoms within 14 days after visiting Wuhan South China Seafood Wholesale Market and 2) those who had pneumonia symptoms within 14 days after returning from Wuhan.^118^

As the outbreak spread, the KCDC broadened the criteria for testing. Starting January 28, 2020, the agency isolated 1) those who had fever or respiratory symptoms upon returning from Hubei province and 2) those who had symptoms of pneumonia upon returning from mainland China.^119, 120^ We coded this as the first change in the testing regime.

The second testing regime change occurred on February 4, 2020, when the KCDC announced that people who had had any “routine contacts” with confirmed cases were required to self quarantine for a 14-day period. The agency defines two categories of contacts: close contacts and routine contacts. The former is defined as a person who has been within two meters of, in the same room as, or exposed to any respiratory secretions of an infected individual. The latter refers to whether the individual conducted any activity in the same place and at the same time as the infected person. Prior to this regime change, the KCDC separated those two cases and applied different quarantine policies; starting February 4, 2020, any routine contacts were also required to be self-quarantined.^121^

Shortly thereafter, South Korea aggressively expanded the scope of their testing. Starting February 7, 2020, the KCDC broadened the definition of suspected cases to 1) anyone who developed a fever or respiratory symptoms within 14 days after returning from China, 2) anyone who developed a fever or respiratory symptoms within 14 days after being in close contact with a confirmed case, and 3) anyone suspected of contracting COVID-19 based on their travel history to affected countries and their clinical symptoms.^122^ Moreover, the KCDC announced that the test would be free for all suspected cases and confirmed cases.^123^ As a result of these efforts, KCDC announced that they would begin to test 3, 000 people daily, a marked increase from only 200 people a day previously.^124^

The KCDC revised their guidelines on February 20, 2020 in order to test more people. Their press release stated: “Suspected cases with a medical professional’s recommendation, regardless of travel history, will get tested. Additionally, those who are hospitalized with unknown pneumonia will also be tested. Lastly, anybody in contact with a diagnosed individual will need to self-isolate, and will only be released when they test negative on the thirteenth day of isolation.”^125^

As the number of patients grew rapidly, the KCDC decided to focus on more vulnerable groups. In their February 29, 2020 press release, the agency stated: “The KCDC has asked local government and health facilities to focus on tests and treatment, especially targeting those aged 65+ and those with underlying conditions who need early detection and treatment.” This change was coded as our next testing regime change in the dataset.^126^

On March 22, 2020, the KCDC began conducting COVID-19 diagnostic testing for every inbound traveler entering from Europe. This was coded as another testing regime change. Of the 1, 442 inbound travelers from Europe arriving March 22, 2020, 152 were symptomatic and were quarantined and tested at an airport quarantine facility. The remaining 1, 290 travelers were asymptomatic and were moved to a temporary living facility to be tested.^127^

On March 27, 2020, this policy was expanded, where all inbound travellers from the US with symptoms (regardless of nationality) were required to be tested at the airport.^128^ We code this as our final testing regime change.

The data on the testing regime date changes are in the *“KOR_policy_data_sources.csv.”*

### Italy

We have collated a regional and provincial level time series health outcome dataset in Italy from February 24, 2020 to April 7, 2020.

This data came from the GitHub repository maintained by the Italian Department of Civil Protection *(Dipartimento della Protezione Civile)*. Health outcomes included the number of confirmed cases, the number of deaths, the number of recoveries, and the number of active cases. These figures have been updating daily at 5 or 6 pm (Central European Time). The regional-level dataset was pulled directly from *“dati-regioni/dpc-covid19-ita-regioni.csv,”* and the provincial-level dataset was pulled from *“dati-province/dpc-covid19-ita-province. csv.”*

#### Testing regime changes

The testing regime change in Italy occurred when the Director of Higher Health Council announced on February 26, 2020 that COVID-19 testing would only be performed on symptomatic patients, as the majority of the previous tests performed were negative.

The data on the testing regime date changes are in the *“ITA_policy_data_sources.csv.”*

### Iran

We have collated a provincial-level time series health outcome dataset in Iran from February 19, 2020 to March 22, 2020 (however health data are not reported by the Iranian Ministry of Health for March 2-3, 2020).

The Iranian government had been announcing its new daily number of COVID-19 confirmed cases at the provincial level on the Ministry of Health’s website. This data has been compiled daily in the table “New COVID-19 cases in Iran by province”^129^ located in the “2020 coronavirus pandemic in Iran” article on Wikipedia.

We spot-checked the data in the Wikipedia table against the Iranian Ministry of Health announcements^130^ using a combination of Google Translate and a comparison^131^ of the numbers in the announcements (which were written in Persian script) to the Persian numbers.

#### Testing regime changes

On March 6, 2020, the Ministry of Health announced^132^ a national coronavirus plan, which included contacting families by phone to identify potential cases, along with the disinfecting of public places. The plan was to begin in the provinces of Qom, Gilan, and Isfahan, and then would be rolled out nationwide. On March 13, 2020, the government announced a military-enforced home isolation policy throughout the nation.^133^ This announcement included nationwide disinfecting of public places. While a follow-up announcement of the March 6 high testing regime stating its complete rollout was not found, the March 13 announcement did reference the implementation of the public spaces component of the earlier plan across the country. We thus assumed that the high testing regime had also been fully rolled out on March 13, 2020.

#### Testing Regime Changes

The data on the testing regime date changes are in the *“IRN_policy_data_sources.csv.”*

### France

We have collated a regional-level time series confirmed cases dataset in France from February 15, 2020 to March 25, 2020, and regional-level time series hospitalization data from March 3, 2020 to April 6, 2020.

We used the number of confirmed COVID-19 cases by *région* from France’s government website.^134^ The sources listed for this dataset were the French public health website, ^135^ the Ministry of Solidarity and Health, ^136^ French newspapers that reported government information, ^137^ and regional public health websites.^138^ Given that these data were not published on a daily basis, we supplemented the dataset by scraping the number of confirmed cases by *région* on the French public health website through March 25, 2020, which is the last date the subnational case data are made publicly available.^139^

Hospitalization data come from the same source^140^ (*Santé Publique France)* as the case data. *Santé Publique France* announced they would stop posting regional-level case data because they were not reliable, and only provide hospitalization data instead.

### Testing Regime Changes

The one testing regime change in France occurred on March 13, 2020 with the beginning of the epidemic “stade 3”, when the government started to give severe cases in hospitals priority for testing. ^141^ The testing regime date changes are encoded within the data cleaning script.

### United States

We have collated a state-level time series health outcome dataset in the United States from January 22, 2020 to April 7, 2020.

The data come from the Github repository associated with the usafacts.org interactive dashboard. As of the time of writing, the data are available <u>here</u>. The repository and dashboard are updated essentially in real-time, at least daily.

#### Testing regime changes

To determine the testing regime, we used estimated daily counts of the cumulative number of tests conducted in every state, as aggregated by the largely crowd sourced effort named “The Covid Tracking Project” (covidtracking.com). We estimated the total number of tests as the sum of confirmed positive and negative cases. For some states and some days, there have been no negative case counts, in which case we utilize just the confirmed positive cases. We also ensured that the confirmed number of positive cases agreed with the counts in the John Hopkins University COVID-19 >4dataset.^142^

We programmatically determined possible testing regime changes by filtering for any consecutive days during which the testing rate increased at least 250% from one day to the next, and where this jump was an increase of at least 150 total tests over one day. After visually inspecting the candidates, we confirmed that the automatically detecting testing regime changes represent visually distinguishable changes in testing rates. The testing regime date changes are encoded within the data cleaning script.

## Policy Data

The policy events, datasets, and sources used in this paper are described below. For each country, the relevant country-specific policies identified were then mapped to a harmonized policy categorization used across all countries.

The policy categories are by default coded as binary variables, where *[policy_variable]* = 0 before the policy is implemented in that area, and *[policy_variable]* = 1 on the date the policy is implemented (and for all subsequent dates until the policy is lifted). There are instances when the value of the policy variable is between 0 and 1; for further details, refer to the Policy Intensity subsection.

The main policy categories identified across the six different countries fall into four broad classes:

1. Restricting travel:

a. *travel_ban_local*: A policy that restricts people from entering or exiting the administrative area (e.g county or province) treated by the policy.
b. *travel_ban_intl_in*: A policy that either bans foreigners from specific countries from entering the country, or requires travelers coming from abroad to self-isolate upon entering the country.
c. *travel_ban_intl_out*: A policy that suspends international travel to specific foreign countries that have high levels of COVID-19 outbreak.
d. *travel_ban_country_list:* A list of countries for which the national government has issued a travel ban or advisory. This information supplements the policy variable *travel_ban_intl_out*.
e. *transit_suspension:* A policy that suspends any non-essential land-, rail-, or water-based passenger or freight transit.
2. Distancing through cancellation of events and suspension of educational/commercial/religious activities:

a. *school_closure:* A policy that closes school and other educational services in that area.
b. *business_closure:* A policy that closes offices, non-essential businesses, and non-essential commercial activities in that area. Non-essential services are defined by area. This policy also includes the limiting of business hours and reducing restaurant and bar operations.
c. *religious_closure:* A policy that prohibits gatherings at a place of worship, specifically targeting locations that are epicenters of the COVID-19 outbreak. See the section on Korean policy for more information on this policy variable.
d. *work_from_home*: A policy that requires people to work remotely. This policy may also include encouraging workers to take holiday/paid time off.
e. *event_cancel:* A policy that cancels a specific pre-scheduled large event (e.g. parade, sporting event, etc). This is different from prohibiting all events over a certain size.
f. *no_gathering:* A policy that prohibits any type of public or private gathering, (whether cultural, sporting, recreational, or religious). Depending on the country, the policy can prohibit a gathering above a certain size, in which case the number of people is specified by the “*no_gathering_size”* variable.
g. *no_gathering_inside:* A policy that specifically prohibits indoor gatherings. See the section on French policy for more information on this policy variable.
h. *no_demonstration:* A policy that prohibits protest-specific gatherings. See the section on Korean policy for more information on this policy variable.
i. *social_distance* A policy that encourages people to maintain a safety distance (often between one to two meters) from others. This policy differs by country, but includes other policies that close cultural institutions (e.g. museums or libraries), or encourage establishments to reduce density.
j. *welfare_services_closure:* A policy that mandates the closure of social welfare facilities, specifically mental rehabilitation facilities, social welfare centers, and homeless use facilities. See the section on Korean policy for more information on this policy variable.
3. Quarantine and lockdown:

a. *pos_cases_quarantine:* A policy that mandates that people who have tested positive for COVID-19, or subject to quarantine measures, have to confine themselves at home. The policy can also include encouraging people who have fevers or respiratory symptoms to stay at home, regardless of whether they tested positive or not.
b. *homejsolation:* A policy that prohibits people from leaving their home regardless of their testing status. For some countries, the policy can also include the case when people have to stay at home, but are allowed to leave for work-or health-related purposes. This policy is also a superset of other, less stringent policies. As such, the estimates of the full home isolation effect encapsulates the effect one would observe if none of the less stringent policies were yet in place when home isolation was implemented.
4. Additional policies

a. *emergency_declaration:* A decision made at the city/municipality, county, state/provincial, or federal level to declare a state of emergency. This allows the affected area to marshal emergency funds and resources as well as activate emergency legislation.
b. *paid_sick_leave:* A policy where employees receive pay while they are not working due to the illness.

### Optional policies

In the cases when the aforementioned policies are optional, we denote this as *[policy_variable]_opt*.

### Population weighting of policy variables

In cases where only a portion of the administrative unit (e.g. half of the counties within the state) are affected by the implementation of the policy, we weight the policy variable by the percentage of population within the administrative unit that is treated by the policy. This is denoted as *[policy_variable]_popwt*, and the value that this variable can take on is a continuous number between 0 and 1. Sources for the population data are detailed in a later section.

### Policy intensity

*policy_intensity* is a continuous value between 0 and 1 that modulates the intensity/restrictiveness of a policy. By default this value is 0 when the policy has not been implemented and 1 when the policy is implemented (i.e. the policy variables are treated as indicator variables). However, in instances when a policy has evolved overtime, then less restrictive implementations of the policy are weighted by a *policy_intensity* value that is between 0 and 1, and the most restrictive version of the policy has a value of 1.

For simplicity, if a given policy has undergone one version change, then *the policy_intensity* of the less restrictive edition is equal to 0.5, and the value of the more restrictive edition is equal to 1. If there have been two version changes, then *the policy_intensity* of the less restrictive edition is equal to 0.33, the value of the second most restrictive edition is equal to 0.67, and the value of the most restrictive edition is equal to 1, etc.

Additionally, in certain countries, a type of broad policy category (e.g. *business_closure)* could have multiple sub-policies that fall within this category (e.g. closing of restaurants/bars, closing of gyms/casinos/recreational facilities, and closing of non-food/non-recreational facilities). In this case, the policies are treated additively, where each of the three sub-policies have a *policy_intensity* weight of 0.33, and if an administrative unit has implemented only two of three of the policies on a given date, then the *business_closure* variable would equal 0.66.

Lastly, for some countries, the deployment of a certain policy (e.g. *home_solation*) implies that other policies are also turned on (e.g. *travel_ban_local* or *work_from_home*), even if there is no separate policy that explicitly states this. The relationships between policies are country-specific, and these assumptions are encoded in our country-by-country policy data dictionary, which can be found in the GitHub repository at *data/raw/multi_country/policy_implication_rules.json*.

In the instances when the implementation of a certain policy implies that other policies are also in place, these secondary policies are only drawn from the set of policies that have already been deployed within that country. For example, in Italy when *home_solation* is enacted for an administrative unit on a specific date, this implies that *school_closure* and *work_from_home* should also be turned on for this administrative unit on this date, since these two policies have been implemented within the country. However, this would not imply that the policy variable *no_demonstrations* (a policy that has been implemented in other countries within our sample, but not in Italy) is turned on, since this policy had not been deployed in any Italian administrative unit prior to this specific date.

More specifically, we compute *policy_intensity* using this approach:

1. For non-population-weighted policy variables: For a given policy category on a specific date (e.g. *business_closure* on March 15, 2020), take the maximum of the mandatory policy intensities for all units lower (e.g*.Adm0)* than, equal to, and higher (*e.g.Adm2*) than the analysis unit (e.g. *Adm1)*. Assign this maximum *policy_intensity* value to the unit of analysis. If there is no mandatory version of the policy that applies to the unit of analysis, then take the maximum of the optional policy intensities and assign it to the optional policy variable for the analysis unit.
2. For population-weighted policy variables: Take the maximum of the mandatory policy intensities for all units lower (e.g. *Adm0)* than and equal to the analysis unit (e.g. *Adm1)*, and assign that as the default mandatory intensity for all units higher *(e.g.Adm2)*. If the policy is not mandatory at the analysis or lower unit, then assign the maximum of the optional *policy_intensity* value as the default optional intensity for all higher units. For any higher unit that has a specific policy, assign the appropriate version (mandatory or optional) of the policy variable at that higher unit the maximum of that intensity and the default intensity, with mandatory always taking priority over optional. For all that don’t have a specific policy, assign them the default intensity (again, assigning this to the optional or mandatory version as appropriate). Then calculate the population-weighting at the analysis unit level (e.g. Adm1), separately for both optional and mandatory variables. Each higher unit should only have a non-zero intensity for optional or for mandatory (or neither), but not both.
3. For broadly defined policy variables like *social_distance* that could encompass a variety of country-specific policies: The *policy_intensity* assignment differs by country. If the specific policies employed at the various administrative levels are the same policy, then the approach in (1) is used. If they are different policies within the same broad category, then we add instead of taking the maximum, allow for both optional and mandatory policies, and and otherwise follow the approach of (1). This addition is appropriate across different administrative divisions because of (1).

### China

We obtain data on China’s policy response to the COVID-19 pandemic by culling data on the start dates of travel bans and lockdowns at the city-level from the “2020 Hubei lockdowns” Wikipedia page, ^143^ and various news reports.

To combat the spread of COVID-19, the Chinese government imposed Level 1 emergency declarations, travel restrictions and quarantine measures. The lockdown of the city of Wuhan, the origin of the pandemic, occurred on January 23, 2020. Immediately following the Wuhan lockdown, neighboring cities followed suit, banning travel into and out of their borders, shutting down businesses, and placing residents under household quarantine. The same policy measures were implemented in cities across China for the next three weeks.

Starting around the same time as the initial lockdown, from January 23 to January 29, 2020, all provincial governments declared Level 1 emergency responses.^144^ The declaration often included the closing of entertainment venues, banning of public gatherings, and extensive temperature monitoring at airports, railway stations and highway checkpoints.^145^

Some lockdowns occurred during the national Chinese New Year holiday (January 24–30, 2020) when schools and most workers were on break. On January 27, 2020, China extended the official holiday to February 2, 2020, while many additional provinces delayed resuming work and opening schools for even longer.^146^ The Chinese New Year holiday is analogous to containment policies such as school closures and restrictions on non-essential work. We do not specifically estimate the effect of this holiday extension, as most cities were in lockdown during the extended holiday, and a lockdown is a more restrictive containment measure. A lockdown requires all residents to stay home, except for medical reasons or essential work, and only allows one person from each household to go outside once every one to five days (exact policy varied by city).

### South Korea

We obtained data on South Korea’s policy response to the COVID-19 pandemic from various news sources, as well as press releases from the Korean Centers for Disease Control and Prevention (KCDC), the Ministry of Foreign Affairs, and local governments’ websites. The policy variables coded in the dataset are: *welfare_services_closure, business_closure_opt, emergency_declaration, no_demonstration, religious_closure, event_cancel, school_closure, social_distance_opt, travel_ban_intl_in_opt, travel_ban_intl_out_opt,, work_from_home_opt*, and *pos_cases_quarantine*.

On February 28 2020, the KCDC recommended the closure of 14 types of social welfare facilities to reduce the spread of infection among vulnerable groups in the population.^147^ These include childcare centers, vocational rehabilitation centers for the disabled, senior citizen centers, mental rehabilitation facilities, and homeless use facilities. We code this in the variable *welfare_services_closure*. Even though it was technically a recommendation, we did not code this policy as optional because a majority of facility types listed in the press release (senior citizen centers, job centers, childcare centers, etc.) are under public administration, so these facilities likely would have followed recommendations. Indeed, some news articles have reported that all children’s centers in Busan are closed^148^ as well as over 3, 600 facilities in Seoul.^149^

We created another variable, *business_closure_opt*, which applies to two provinces: Seoul and Gyeonggi-do. On March 11, 2020, the mayor of Seoul advised that popular commercial establishments such as karaoke places, clubs, and cyber cafes be closed.^150^ Seven days later, the governor of Gyeonggi-do issued an executive order limiting the usage of commonly frequented commercial establishments and requiring a higher standard of cleanliness.^151^ We coded this as an optional business closure given that the policy discourages usage of these facilities but did not explicitly order them to shut down.

Daegu and Gyeongsangbuk-do have been two of the regions hardest hit by COVID-19. The government of South Korea declared an emergency for those two areas on March 15, 2020.^152^ We incorporated this information into the variable *emergency_declaration*.

The variable “ *no_demonstration* reflects the efforts of some regions limitingany protests callingfor slowing the spread of the outbreak. On February 24, 2020, Incheon stopped a protest in front of the Incheon Metropolitan City Hall.^153^ Two days later, Seoul prohibited protests in downtown areas where massive demonstrations used to take place.^154^

Many province level COVID-19 policies have targeted religious gatherings at Shincheonji Church of Jesus, since its religious gatherings have been linked to the explosion in the number of cumulative confirmed cases. Provincial governments tried to shut down Shincheonji-related places of worship, and the related policy implementation is encoded in the variable *religious_closure*. The regions which utilized this policy option are: Daegu, ^155^ Gyeongsangbuk-do, ^156^ Seoul, ^157^ Jeju, ^158^ Gyeonggi-do, ^159^ Jeollanam-do, ^160^ Gyeongsangnam-do, ^161^ Incheon, ^162^ Ulsan, ^163^ Busan, ^164^ Jeollabuk-do, ^165^ Chungcheongbuk-do, ^166^ Gwangju, ^167^ Chungcheongnam-do, ^168^ and Daejeon.^169^

Many provinces have also canceled public events organized by local administrative agencies. We code this policy in the variable *event_cancel*. The regions which exercised this policy are: Seoul, ^170^ Daegu, ^171^ Gangwon-do, ^172^ Chungcheongbuk-do, ^173^ Chungcheongnam-do, ^174^ Sejong, ^175^ Daejeon, ^176^ Gyeongsangbuk-do,^177^ Gyeongsangnam-do, ^178^ Jeju, ^179^ Gyeonggi-do, ^180^ Ulsan, ^181^ Gwangju, ^182^ Busan, ^183^ Incheon, ^184^ Jeollanam-do, ^185^ and Jeollabuk-do.^186^

The policy variable *school_closure* has been turned on for the entirety of the Korean time series dataset. This is because all schools were already on vacation during the beginning of the outbreak, and the government then postponed their start dates. At the time of writing, the Ministry of Education announced that schools would be kept closed until April 3, 2020.^187^ Therefore, this policy variable is always equal to 1 in the dataset.

*social_distance_opt* has been turned on from February 29, 2020, when KCDC recommended social distancing as one of the main tools to deal with the outbreak. In their press release, they recommended that people maintain personal hygiene and practice ‘social distancing’ until the beginning of March, an important point of this outbreak.^188^ In the case of Daegu, the hardest-hit region in the country, we coded the variable as 1 starting from February 22, 2020, based on the statement, “It is recommended for residents in Daegu to minimize gathering events and outdoor activities.”^189^

The first travel restriction for incoming travelers *(travel_ban_intl_in_opt)* was implemented on January 28, 2020. It is worth noting that it was not a total prohibition of incoming visitors; rather, it means inbound travellers were subject to COVID-19 specific emergency measures. KCDC mentioned that starting on January 28, 2020 “any travellers departing] from China [would] be a subject to strengthened screening and quarantine measures.”^190^ On February 12, 2020, KCDC broadened the list of countries subject to the stricter measures to include Hong Kong and Macau.^191^ Subsequently, KCDC added Italy and Iran (on March 11, 2020);^192^ France, Germany, Spain, UK, and Netherlands (on March 15, 2020);^193^ and any remaining European countries (March 15, 2020)^194^ to their country list. On March 19, 2020, the policy was expanded to include all travelers arriving at port regardless of country of origin.^195^

This restriction was not limited to inbound travellers. The government also issued advisories on countries where the number of infections had increased, which has been encoded as the variable *travel_ban_intl_out_opt*. The first outbound travel alert due to COVID-19 was announced on January 28, 2020: The Ministry of Foreign Affairs (MOFA) issued a Level 2 (Yellow) alert for any travel to mainland China, Hong Kong, and Macau.^196^ Later, MOFA added Italy on February 28, 2020, ^197^ Japan on March 9, 2020, ^198^ and all European countries on March 16, 2020.^199^ On March 18, 2020, KCDC strongly called for the cancellation or delay of all international travel on non-urgent matters.^200^ It should be noted that the Level 2 alert does not enable the government to prohibit travel to these destinations, which is why the policy was coded as optional.

There are four types of travel advisories distributed by the South Korean government: Level 1, Navy; Level 2, Yellow; Level 3, Red; and Level 4, Black.^201^ Travel under the Level 4 alert is prohibited, and the government utilizes legal instruments to enforce the restriction. If people leave the country under the black alert, they will be subject to fines up to ten million KRW, or imprisonment up to a year. However, there is no enforcement instrument for the advisories up to Level 3. In that sense, we stated above that the banning policy does not mean prohibiting travel. Nevertheless, we coded the yellow alert as the first travel ban in our dataset, since Level 2 alerts are issued relatively rarely, such as during a significant demonstration^202^ or military coup.^203^ As a result, we coded the Level 2 alert due to COVID-19 into the dataset for the policy analysis.

The policy variable *“work_from_home_optional* indicates when KCDC began recommending that people work from home. On March 15, 2020, the KCDC press release stated: “Since contact with confirmed cases in an enclosed space increases the possibility of transmission, it is recommended to work at home or adjust desk locations so as to keep a certain distance among people in the office. More detailed guidelines for local governments and high-risk working environments will be distributed soon.”^204^

On March 22, 2020, the KCDC announced that all inbound travelers from Europe would be tested at the airport and subject to quarantine measures.^205^ Korean citizens and long-term visitors returning from abroad needed to home-quarantine for 14 days (even if they test negative for COVID-19), while short-term visitors would be actively monitored. Inbound travelers with no symptoms were required to stay at temporary facilities while awaiting their test results.^206^ We coded this as the policy variable *pos_cases_quarantine* modulated *by policyjntensity* = 0.25. When this policy was expanded on March 27, 2020 to include all symptomatic travelers arriving from the US, ^207^ we coded this variable with a *policy_intensity =* 0.5. On April 1, 2020, these quarantine measures were extended to include inbound travelers arriving from all countries, with exceptions allowed only for limited cases (diplomatic missions etc.).^208^ This variable was then coded with *policyjntensity* = 0.75. Lastly, starting on April 5, 2020, the KCDC announced that inbound travelers who fail to comply with quarantine regulations are subject to imprisonment of up to 1 year or a fine of up to 10 million won for the violation of the Infectious Disease Control and Prevention Act. In addition, persons of foreign nationality who fail to comply may be subject to measures including deportation and entry ban in accordance with the Immigration Act.^209^ We then coded this variable with *the policyjntensity =* 1.

### Italy

We have obtained data on Italy’s policy responses to the COVID-19 pandemic primarily from the English version of the COVID-19 dossier “Chronology of main steps and legal acts taken by the Italian Government for the containment of the COVID-19 epidemiological emergency”^210^ written by the Department of Civil Protection (*Dipartimento della Protezione Civile)*, most recently updated on March 12, 2020. This dossier details the majority of the municipal, regional, provincial, and national policies rolled out between the start of the pandemic to present-day. We have supplemented these policy events with news articles that detail which administrative areas were specifically impacted by the additional policies.

The first major policy rollout was on February 23, 2020, when 11 municipalities across two provinces in Northern Italy were placed on lockdown. These policies included closing schools, cancelling public and private events and gatherings, closing museums and other cultural institutions, closing non-essential commercial activities, and prohibiting the movement of people into or out of the municipalities.

The second major policy rollout was on March 1, 2020, when two provinces and three regions in Northern Italy were placed on partial lockdown. These policies also included closing schools, cancelling public and private events and gatherings, closing museums, closing non-essential commercial activities, as well as limiting the number of people at places of worship, restricting operating hours of bars and restaurants, and encouraging people to work remotely.

The third major policy roll-out was on March 5, 2020, when all schools across the country were closed.

The fourth major policy roll-out was on March 8, 2020 when the region of Lombardy and 13 provinces in Northern Italy were placed on lockdown. These policies included the cancellation of public and private events and gatherings, closing of museums, encouraging people to work remotely, limiting the number of people at places of worship, restricting opening hours of bars and restaurants, mandating quarantine of people who tested positive for COVID-19, prohibiting the movement of people into or out of the affected area, and restricting movement within the affected area to only work or health-related purposes. Commercial activities were still allowed, as long as they maintained a safety distance of one meter apart per person within the establishment. All civil and religious ceremonies, including weddings and funerals, were suspended. During this same policy roll-out, the rest of the country faced less stringent policies: cancelling public and private events, closing museums, and requiring restaurants and commercial establishments to maintain a safety distance of one meter apart per person within the establishment.

The fifth major policy roll-out was announced on March 9, 2020, and went into effect on March 10, 2020, when lockdown policies applied to Northern Italy were rolled out to the entire country. Lastly, on March 11, 2020, the lockdown was changed to also cover the closing of any non-essential businesses and further restricted people from leaving their home.

After the death toll in Italy surpassed that of China on March 21, 2020, the Italian government increased the severity of their existing policies. Effective March 22, 2020, all non-essential industrial production and factories would be shut down across the country.^211^ Domestic travel was further restricted; people were not permitted to leave the municipality they were currently in except for urgent matters or emergencies.^212^ Lastly, in the hard-hit northern region of Lombardy, the regional government increased lockdown restrictions by banning all individual outdoor exercise or sporting activity.^213^

*Policy Intensity:* We have modified the policy intensity of three different policy variables: *home_isolation, business_closure*, and *travel_ban_local*.

The *home_isolation* policy underwent three policy revisions:

1. The least restrictive version of the policy applies to when people were allowed to leave the house for work, health, and essential reasons *(policy_intensity of home_isolation* = 0.33).
2. The moderate version of the policy applies to when people were allowed to leave the house only for health and essential reasons (which includes the ability to go outdoors for individual exercise/sporting activities) (*policy_intensity of home_isolation* = 0.67).
3. The most restrictive version of the policy applies to when people were allowed to leave the house only for health and essential reasons, but were no longer allowed to leave the house for individual exercise/sporting activities *(policy_intensity of home_isolation* = 1).

The *business_closure* policy underwent three policy revisions:

1. The least restrictive version of the policy applies to the limiting of restaurant hours (but other commercial activities were permitted) (*policy_intensity* of *business_closure* = 0.33),
2. the moderate version of the policy applies to the closing of all non-essential businesses, *(policy_intensity* of *business_closure* = 0.67),
3. and the most restrictive version of the policy applies to the closing of all non-essential industrial production and factories, in addition to the closing of non-essential businesses (*policy_intensity* of *business_closure* = 1).

Lastly, the *travel_ban_local* policy underwent two policy revisions:

1. The least restrictive version of the policy applies to when people were not allowed to enter/exit the affected administrative area, *(policy_intensity* of *travel_ban_local* = 0.5),
2. and the most restrictive version of the policy applies to a more restrictive ban on domestic travel that mandated that people had to stay in the municipality they were currently in (*policy_intensity* of *travel_ban_local* = 1).

### Iran

For Iran’s policy response to the COVID-19 pandemic, we relied on news media reporting as the primary source of policy information (mostly due to translation restrictions). We also relied on two timelines of pandemic events in Iran to help guide the policy search.^214, 215^

The first major outbreak in Iran was connected to a major Shia pilgrimage in the city of Qom that brought Shiite pilgrims from Iran and throughout the Middle East, where they came to kiss the Fatima Masumeh shrine. It is possible that the disease was brought to Qom by a merchant traveling from Wuhan, China.^216^ In addition, it is believed that the Iranian government knew of the COVID-19 outbreak prior to its February 21, 2020 parliamentary elections, but downplayed the risks associated with the disease as not to suppress voter turnout (given concerns that a low turnout would reflect poorly on its legitimacy).^217^ The disease, initially centered in Qom and neighboring Tehran, spread rapidly throughout the country.

As the number of cases grew, the Iranian government started to increase the stringency of its response. The first case was reported on February 19, 2020 (two individuals who both were reported to have died that day). The next day, school closures were announced in the province of Qom and travel in the region was discouraged. By February 22, 2020 the government closed schools in 14 provinces and closed down major gathering sites such as football matches and theaters. By March 5, 2020 schools were closed nationwide and government employees were required to work from home. Home isolation was implemented by the military on March 13, 2020, which the media described as “the near-curfew follows growing exasperation among MPs that calls for Iranian citizens to stay at home had been widely ignored, as people continued to travel before the Nowruz New Year holidays.”^218^

### France

We obtain data on France’s policy response to the COVID-19 pandemic from the French government website, press releases from each regional public health site, and Wikipedia.

The French government website contains a timeline of all national policy measures.^219^ Each regional public health agency (*l’Agence Régionale de Santé)* in France posts press releases with information on the policies the region or departements within the region will implement to mitigate the spread and impact of the COVID-19 outbreak.^220^ The Wikipedia page on the 2020 coronavirus pandemic in France has collated information on the major policy measures taken in response to the COVID-19 pandemic.^221^

Starting February 29, 2020, France banned mass gatherings of more than 5, 000 people nationwide, while some major sporting events were cancelled and a handful of schools closed to mitigate the spread of the virus. As more COVID-19 cases were confirmed during the following week, additional sporting events were canceled, more schools decided to close, and certain cities and *départements* limited mass gatherings to no more than 50 people, excluding shops, business, restaurants, bars, weddings, and funerals. Some régions closed early childhood establishments (e.g. nurseries, daycare centers) and prohibited visitors to elderly care facilities. On March 8, 2020, France banned mass gatherings of more than 1, 000 people nationwide. Other schools, cities, and départements followed suit with additional school closures and limiting mass gatherings. On March 11, 2020, France prohibited all visits to elder care establishments. Starting March 16, 2020, France closed all schools nationwide. Between March 17, 2020 - March 23, 2020, governments at both the national level and région level implemented more restrictive lockdown policies, which included shelter-in-place measures,^222^ the closing of public places,^223^ and banning of outside markets and severely restricting movement outside of the house.^224^

We have coded various policies that cancel events and large gatherings as such: any cancellations of professional sporting and other specific pre-scheduled events as the policy variable *event_cancel*. The *no_gathering* policy variable represents policy measures that banned all events or mass gatherings of a certain size, e.g. no gatherings of over 1, 000 people. The *social_distance* policy variable includes measures preventing visits to elder care establishments, closures of public pools and tourist attractions, and teleworking plans for workers.

### United States

For the United States’ policy response to the COVID-19 pandemic, we relied on a number of sources, including the U.S. Center for Disease Control (CDC), the National Governors Association, individual state health departments, as well as various press releases from county and city-level government or media outlets. The CDC has posted and continually updated a Community Mitigation Framework that encompasses both mandatory and recommended policies at a national level.^225^,^226^ The CDC’s framework along with the guidelines released by the White House on March 16, 2020 named “15 Days to Slow the Spread” included a set of social distancing guidelines that urged Americans to work from home, engage in remote schooling, avoid unnecessary shopping trips, refrain from visiting restaurants and bars, and not gather in groups of more than 10 people.^227^ These guidelines were interpreted by individual governors as they each declared their own States of Emergency at various subsequent dates, and later released their own community mitigation plans or executive orders.

Some of the first states to release such plans included Massachusetts, California, Florida, Washington, and New York, which included both mandatory and optional policies to prevent the COVID-19 spread. ^228^ In addition the National Governors Association has served as a resource for individual states’ policies in response to COVID-19, updating each states’ policy rollout timelines as well as providing links to states’ Executive Orders and other official policy documentation.^229^ To supplement both national and state level policies and recommendations, data was collected, when possible, for cities and counties that have also taken on the role of providing guidance and implementing policies to mitigate the spread of COVID-19.

There have been a wide range in regulatory responses since the first case of COVID-19 was announced in Washington State on January 14, 2020. As a result, the CDC and the Department of State began releasing guidance to those at risk of being exposed to the virus. The initial recommendations included travel warnings for specific countries with confirmed cases and sustained COVID-19 spread. Over the course of our dataset, these warnings increased in intensity, changing from warning against inbound and outbound travel to specific countries in both Europe and Asia to warning against travel at all.^230^ International travel restrictions were coded as *travel_ban_int_out* for outbound travel, and *travel_ban_int_in* for inbound travel, with lists of the places to and from which travel was restricted also included. On March 31, 2020, the US changed its global travel warning to Level 4, the highest warning level, which the US Department of State defines as avoiding “all international travel due to the global impact of COVID-19.”^231^ In addition to the national travel restrictions, individual states also implemented local travel bans, coded as travel_ban_local as the spread continued to grow, such that anyone entering specific states in which this policy was in effect were required to self-quarantine for 14 days. This ultimately reflected the national policy as well, in that people could still technically travel under a Level 4 warning, but upon arrival to the US, they would be put in a mandatory quarantine for 14 days.

In addition to travel restrictions, as COVID-19 prevalence increased within the US, the CDC began to release additional guidance for healthcare workers, individuals at higher risk, as well as for state-level action (e.g. travel or social distancing policies),^232^ and these policies have largely been implemented at the state-level rather than at the national level.

Social distancing guidelines were among the first and most widely implemented policies across states, but there exists a fair amount of variation in the specifics of each policy between states. As a result, *social_distancing* encompasses a wide range of mandatory and optional guidelines that all were focused on keeping people apart. Specifically, this policy variable was coded as a number of subcategories to capture the fact that it was implemented in a piecemeal manner within and across states.^233^,^234^

These are the policies included within each of the *social_distance* subcategories:

- *Isolate certain populations:* recommend or mandate the isolation of populations such as the elderly, immunocompromised or those who have recently returned from a cruise
- *If outside the home, must abide by social distancing standards:* require a six foot minimum distance from others outside the home, maintain distance when riding public transportation, ask that businesses restrict the number of people within storefront at a time as well as restricting certain types of activities that involve physical interaction with customers (e.g.., bagging groceries, taking cash payment)
- *Mandate mask wearing:* require people to wear a mask outside the home
- *Close public facilities:* close libraries, museums, flea markets, historic sites, memorials, and polling locations
- *Close outdoor facilities:* close beaches, state parks, public parks, public toilets, lakes, and campgrounds.
- *Social distance restriction of visitation to certain facilities:* restrict visitation to prisons, long term care facilities, child care facilities, and homeless shelters, stop elective medical and veterinary procedures, and bar short term rental accommodations.
- *Suspend non-critical state operations/government services:* close government buildings, stop in person meetings of people working for the state, suspend court operations, waive or extend licensing, and permit certain types of work to be carried out remotely, when normally could not (e.g.; notaries, police work, licensing)

Along the same line of social distancing policies, a separate variable was coded as *no_gathering* to represent policy measures that banned all events or mass gatherings of a certain size, i.e. no gatherings over a certain number of people (where this number has varied by region).

In addition, many governors mandated statewide school closures at the private and public K-12 and higher education levels, while others have left it up to each school district to decide.^235^ School closures have been coded as *school_closure* and once implemented, have been “turned on” for the remainder of our time series, as no schools have reopened since these policies have been implemented.

Business closures, coded as *business_closure*, have also been recommended or mandated at the state level. These policies have ranged from shutting down all non-essential businesses, reducing the number of hours a business may be in operation, severely restricting the number of customers that are allowed inside at one time, to prohibiting customers to enter a business, such as in the case of bars and restaurants, where they were only allowed to operate or take-out and delivery services.

When business closures have involved shutting down all non-essential operations, “essential” has been defined by each state but is largely similar between states, generally defining essential as food or healthcare providers, as well as basic government operations (i.e. trash collection, mail, water monitoring, etc). To support employees working remotely or staying home when sick, a number of states have also mandated paid sick leave for those who are affected by COVID-19, which has been coded as *paid_sick_leave*. There is a separate *work_from_home* category that includes measures that require businesses to allow employees to telework, if possible, such that no workers except for those who have essential functions are allowed to work in an office.

At the subnational level, many governors have implemented a statewide mandatory shelter-in-place policy, requiring all individuals to self-isolate within their home or place of residence and limit outdoor activity to essential functions only, which is defined by each state. Shelter-in-place laws have been coded as *home_isolation*, and generally are enacted alongside a number of other policies, including business closures, local travel bans, more restrictive gathering sizes, and enforceable social distance rules. Again similar to business closures, the definition of “essential function” has been updated in subsequent policy editions to be more detailed and often stricter, allowing for less activity out of the home.^236^

Both the *emergency_declaration* and *event_cancel* variables were included in the raw data, but were not included in the final analysis.

## Population Data

In order to construct population weighted policy variables and to determine the susceptible fraction of the population for disease projections under the realized and the no-policy counterfactual scenarios, we obtained the most recent estimates of population for each administrative unit included in our analysis. The sources of that population data are documented below.

### China

City-level population data have been extracted from a compiled dataset of the 2010 Chinese City Statistical Yearbooks. We matched the city level population dataset to the city level COVID-19 epidemiology dataset. As the two datasets use slightly different administrative divisions, we only matched 295 cities that exist in both datasets, and grouped the remaining 39 cities in our compiled epidemiology dataset into “other” for prediction purposes. Cities grouped into “other” because of mismatches have a total population of 114, 000, 000, or 8.5% of the total population in China.

For these 39 cities that we could not match, we imputed the population by taking the total remaining population (114, 000, 000) and divided it evenly between these remaining cities. We flag the imputed populations by using the binary variable *pop_is_imputed*.

### South Korea

We downloaded the number of population by provinces from a webpage administered by the Korean Statistical Information Service (KOSIS).^237^ The government agency recently updated the population information of February, 2020, which we used for our analysis.

### Italy

Region and province level population data come from the Italian National Institute of Statistics (Istat), estimating total population on January 1, 2019. The datasets for all Italian regions and provinces are scraped from <u>Istat’s website</u> in *get_adm_info.ipynb*.

### Iran

Province level population data for Iran comes from the 2016 Census, as listed on the *City Population* website.^238^ It is scraped in *get_adm_info.ipynb*.

### France

*Département-*level populations are obtained from the National Institute of Statistics and Economic database.^239^ We used the most up to date estimation of the population in France as of January 2020.

### United States

State- and county-level population data come from the 2017 American Community Surveys dataset, and is downloaded via the *census* Python package^240^ in *get_adm_info.ipynb*.

## Supplementary Methods

In our Supplementary Methods, we describe several sensitivity analyses performed to assess the robustness of our growth rate impacts and projections of cases averted/delayed.

This section is divided into five analyses:

1. Testing the sensitivity of projected averted/delayed cases to varying epidemiological parameters
2. Testing the sensitivity of our regression model to varying epidemiological parameters
3. Testing the sensitivity of our regression model to changes in lag structure
4. Testing the sensitivity of our regression model to withholding of data
5. Testing the sensitivity of our regression model to changes in policy groupings

## 1. Sensitivity of projected averted/delayed cases to the removal rate *γ* the use of an SEIR framework, and the infection-fatality ratio

We compute the empirical removal rate using aggregated data from the countries for which we observe active cases (i.e., China and South Korea) and estimate a value of *γ* = 0.079 (see the Methods section of the main text). This value measures the inverse of the mean duration from being *reported* as infected to being *reported* as recovered (or dead) and may differ from the fundamental epidemiological parameter describing the rate of removal from the pool of infectious individuals. While our estimate implies a recovery period (symptom onset to recovery) that is comparable to some estimates in the literature (median time of 19-23 days, varying based on age group, sex, severity, and mode of detection^241^), we test the extent to which our simulation results in Figure 4 depend on this value (Extended Data Figure 7). One motivation for this exercise is that there may be an unknown delay between the time when a patient becomes non-infectious and the time in which they are recorded in the aggregate data as recovered. For example, assuming a conservatively high value of 14 days for the average delay between “no longer infectious” and “confirmed recovered,” our empirical estimation yields *γ* = 0.18.

In addition, the use of an SIR framework may misrepresent the true underlying disease dynamics, and a more general SEIR framework, which includes representation of people exposed to the infection without yet being infectious, may produce more realistic simulations of cases averted/delayed by policy. We also test sensitivity to the use of the SEIR framework, as well as a key parameter in this alternative framework -- the assumed rate of transition from exposed to infectious (*σ)*.

We replicate the simulation underlying Figure 4 using an SEIR framework with values of *γ* = {0.05, 0.1, 0.15, 0.2, 0.25, 0.3, 0.35, 0.4} and *σ* ∊ {0.2, 0.33, 0.5, ∞}, with (*σ* = ∞ corresponding to the SIR framework we employ in the main simulation. We present our estimates of the total number of cases under both the no-policy and policy scenarios, as well as the total number of cases averted/delayed by policy. We sum simulated cases across all countries on the last dates of the countries’ respective samples. Note that the simulation uses the growth rates derived from our empirical model such that changes in *γ* and *σ* correspond to changes in the transmission rate *β · β* must vary with *γ* and *σ* as our data determine the underlying exponential growth rate. We show the results of this sensitivity analysis in Extended Data Figure 7.

Panels (**a**) and (**b**) respectively show the simulated number of cases in the no-policy and with-policy scenarios. The number of simulated no-policy cases is decreasing in *γ* for high *σ* and increasing in *γ* for low *σ*. The number of simulated no-policy cases is increasing in *σ* for low *γ* and nonmonotonic in *σ* for high *γ*. The number of simulated with-policy cases is increasing in *γ* and decreasing in *σ*. Panel (**c**) shows the number of cases averted due to policy and demonstrates that varying *σ* or *γ* can reduce our estimate of cases averted on the order of several million reported cases (up to 10%). Panel (**c**) shows that higher values of *γ* produce lower estimates of cases averted for the SIR model (*σ* = ∞), but increasing estimates of cases averted for the lower values of *σ*. Panel (**d**) plots the content of Panel (**c**) on the log scale used in Figure 4 of the main text for comparison. For our simulation value of *γ*, decreasing *σ* decreases our estimate of cases averted.

Overall, these results show that the order of magnitude of the number of cases averted is preserved within this reasonable range of potential *γ* and *σ* values.

We also compute country-specific estimates of infection underreporting to improve the outbreak simulations in Figure 4. To produce these estimates, we use code from Russell et al. and substitute the assumed case-fatality ratio (a key parameter in their model) for an infection-fatality ratio (IFR) of 0.75%.^242, 243^ Their analysis produces country-specific estimates of infection underreporting, which we use to scale our estimates of confirmed cases to estimate total infections. We test the sensitivity of our main results to this IFR assumption in Supplementary Table 6, where we also show results for the total number of confirmed cases and infections avoided/delayed using IFR assumptions of 0.5%and 1%. The table shows approximately a 70% increase in the estimated number of confirmed cases avoided/delayed for a doubling in the IFR. However, the estimated number of infections is relatively stable with approximately a 15% decline in the estimated number of infections for a doubling in the IFR.

## 2. Sensitivity of exponential regression model to varying epidemiological parameters

The model we use to estimate the impacts of policy on growth rates assumes exponential growth, which is typically valid for early-stage disease outbreaks. If growth is not exponential, there exists the potential for bias in estimated coefficients. There are three primary reasons why an early-stage outbreak could exhibit non-exponential growth in the absence of policy intervention:

1. The infection spread may progress quickly, lowering the susceptible fraction of the population to a degree that materially affects the infection spread, transitioning the outbreak away from the exponential “early stage” regime.
2. In a disease with a substantial latent period, the growth of infections is only asymptotically exponential.^244^ At any given moment in time, the instantaneous growth rate may differ from a steady-state exponential growth rate.
3. When analyzing growth in cumulative infections, as we do for countries where active infection data are unavailable, growth is similarly only asymptotically exponential.

In our dataset, 95% of administrative units have susceptible fractions above 0.93 on their last analysis day and all have susceptible fractions above 0.78, indicating that the first reason is unlikely to induce substantial bias in our results. When the transmission rate of a disease declines due to anti-contagion policy, the growth rate in infections decreases with a lag due to the dynamics associated with the latent period. Because of this, exponential models estimating the average treatment effect (ATE) of a policy may underestimate the true reduction seen from a policy because they include days in which the growth rate was still higher than the new steady state growth rate. Finally, in the early stages of an outbreak, the number of active cases will dominate the number of recovered/deceased patients and thus the differences in growth of active and cumulative cases is likely to be small.

To test the robustness of our regression approach, we construct simulated outbreaks in which we control demographic, policy, and epidemiological parameters. We then use a variant of the regression model (Eq. 7) from the main text to estimate the no-policy growth rates and the effects of each policy. In this simulation, we do not include any fixed effects to control for day-of-week (*δ*) and changing testing regime (*μ*) effects. These variables are not simulated as these are primarily measurement controls and their effects would be directly absorbed by the corresponding regression parameters if simulated. We compare the coefficient estimates to the “true” values used in the simulation.

To capture the impact of disease latency, we use an SEIR model framework to generate synthetic outbreaks. We simulate 12, 000 45-day outbreaks at hourly timesteps, starting with a no-policy exponential growth rate of 0.4 (similar to those estimated in our main analysis) and a single exposed individual. We adjust this asymptotic exponential growth rate to account for three synthetic policies that turn on at random times, each with a known effect (−0.05, −0.1, and −0.2). For each subset of 1, 000 simulations, we use one of four plausible values for the mean latency period, *σ^-^*^1^ (0, 2, 3, and 5 days), and one of three plausible values for the mean infectious period, *γ*^-1^ (3, 5, and 20 days). We choose a wide range of these variables due to substantial uncertainty over the epidemiological characteristics of the novel coronavirus,^245^ and a nonexistent latency period is included for comparison to an SIR-like data generating process. The mean transmission rate per infected person per day, β, is derived from the asymptotic growth rate, the mean latency period, and the mean infection period by solving for the eigenvalues of a SEIR system,^246^ which yields:

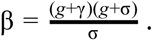

We apply exponential noise to β for each simulation and at each timestep, and contribute additional gaussian noise to σ and *γ* (standard deviations of 0.01 and 0.03, respectively). We add additional gaussian “measurement noise” to the instantaneous growth rates after simulation but before running our regression (standard deviation of 0.1). Cumulatively, this results in an average root-mean-squared-error (RMSE) in regressions across all 12, 000 simulations of ~0.11, which matches the RMSE of daily growth rate values across all six countries in our main analysis.

The dynamic model outputs a time series of susceptible (S), exposed (E), infectious (I), and removed (R) individuals, as a fraction of the total population. We use both I and I+R as the left-hand-side variables in our regression framework. The former corresponds to the analysis we run for countries in which we observe active cases and the latter to countries in which we observe only cumulative cases. We assume the majority of the “exposed but not infectious” population will not yet have been tested and will not appear in any of the datasets used in the main analysis. Our right-hand-side variables consist of the binary policy variables, allowing only for contemporaneous effects. This matches our main specification for all countries except China (where data availability allows for the estimate of lagged effects) and provides the most challenging environment in which to estimate the effect of policy in a dynamic system.

Results are presented in Extended Data Figures 8 and 9. While it is possible to simulate outbreaks consisting of innumerable parameter combinations and noise distributions, we display those that seem most relevant for evaluating the robustness of our main analysis. Our associated GitHub repository contains a Jupyter notebook for readers to further examine the effect of simulation configurations on regression model robustness.

Figures 8 and 9 are each split into two panels (a) and (b). Panel (a) of each figure shows simulations in near-ideal data conditions, in which we observe active infections within a large population. This means that the susceptible fraction of the population remains high during the entire sample period. For example, these conditions are similar to those in our real data for Chongqing, China. Panel (b) of each figure shows simulations in a non-ideal data scenario where we are only able to observe cumulative infections in a small population. In these simulations, the susceptible fraction declines to values as low as 33% of the population. For example, these conditions are similar to those in our real sample of data for Cremona, Italy.

Figure 8 demonstrates that our model recovers unbiased estimates of the no-policy growth rate under all conditions simulated. Because the growth rate prior to policy has likely approached its asymptotic rate by the time we begin our regressions, variance in our no-policy growth rate estimates comes from noise in the disease parameters and measurement. The ability to recover unbiased estimates of this value has important implications for our estimate of the total number of cases averted/delayed to date, as this number is primarily driven by the counterfactual number of cases we would expect to see in a world in which no anti-contagion policy was enacted.

Figure 9 demonstrates that our model recovers unbiased estimates of the cumulative effect for a disease with very short latency. As the latency period increases, the model begins to slightly underestimate the true effect of policy (i.e. it predicts a less negative value), due to the decay time over which a shock to transmission rate propagates to a new steady-state growth rate. The underestimate is reduced in situations where we are able to directly observe active infections and is increased when we can only observe cumulative infections. Note that statistical uncertainty in these estimated parameters dominates potential biases, even in “worst case” data conditions.

We conclude that biases (due to the use of an exponential model) in our estimates of the no-policy growth rate are essentially zero and are likely to be small and negative for our estimates of policy effectiveness. If present in the data, such biases would cause us to modestly understate the effectiveness of anti-contagion policies.

## 3. Sensitivity of estimates to changes in lag structure

Existing evidence has not demonstrated whether policies should affect infection growth rates in the days immediately following deployment. It is therefore not clear *ex ante* whether the policy variables in Equation 7 should be encoded as “on” immediately following a policy deployment. As a robustness check, we estimate “fixed-lag” models in which a fixed delay between a policy’s deployment and its effect is assumed. Specifically, we assume that policies cannot influence infection growth rates for *L* days, recoding a policy variable at time *t* as zero if a policy was implemented fewer than *L* days before *t*. We re-estimate Equation (7) for each value of *L* and present results in Extended Data Figure 5 and Supplementary Table 5. If a delay model is more consistent with real world infection dynamics, these fixed lag models should recover larger estimates for the impact of policies and exhibit better model fit.

Panel a of Extended Data Figure 5 displays the *R*^2^ associated with each country-level fixed lag model with fixed lag lengths ranging from no fixed lags up to a 15 day fixed lag. In-sample fit generally declines or remains unchanged if policies are assumed to have a delay longer than 4 days. Panel b of Extended Data Figure 5 plots the estimated effects for no lag (the model reported in the main text) and for fixed-lags between one and five days. Estimates generally are unchanged or shrink towards zero (e.g. *homejsolation* in Iran), consistent with miscoding of post-policy days as no-policy days.

In Supplementary Table 5, we show our estimates of the effect of all policy interventions in each country (analogous to the average difference between red and blue markers in Figure 3 of the main text) using a fixed lag of up to 5 days. The estimated effects are broadly consistent across different lag lengths; however, the magnitude of the effect size declines slightly with increasing lag lengths. If policies take several days to impact infection growth rates, we would expect effect sizes to increase rather than decrease with lag lengths. Our finding of declining effect sizes is more consistent with contamination of the control group, where policies are incorrectly encoded as zeros after they have been deployed.

## 4. Sensitivity of estimates to withholding of data

To ensure that the estimates from our regression model are robust to the withholding of data, we re-estimate our main model *k_c_* number of times for each country, where *k_c_* is the number of first level administrative units (Adm1, i.e. state or province) in country c. In each of the *k_c_* regressions for country c, we withhold data from one Adm1 unit when we estimate the effects of policy interventions on growth rates. The results of this exercise are displayed in Extended Data Figures 3 and 4.

## 5. Sensitivity of estimated growth rates to changes in policy groupings

In our main regression model, due to the limited length of our time series data and instances where multiple policies are deployed on the same date, we group certain policy interventions together. We group policies together that have similar objectives (e.g. travel_ban_local and *transit_suspension* would be one group, *event_cancel* and *no_gathering* would be another) and keep certain policies separate (i.e. *business_closure, school_closure, home_isolation)* where possible.

To test the sensitivity of our results to the grouping of policy interventions, we also estimate a model where the policies are estimated without grouping. Extended Data Figure 6 panel a shows the estimated infection growth rates and no-policy counterfactual growth rates using the model with disaggregated policies. Additionally, in Supplementary Table 4, we compare the effect of policy interventions in each country when the effect of all policies are estimated separately (“Disaggregated Model”) and when they are grouped into policy packages as in our preferred specification (“Main Specification”). We find the estimated impact of policy interventions on case growth rates is robust to this alternative specification.

## Supplementary Tables

**Supplementary Table 1.**
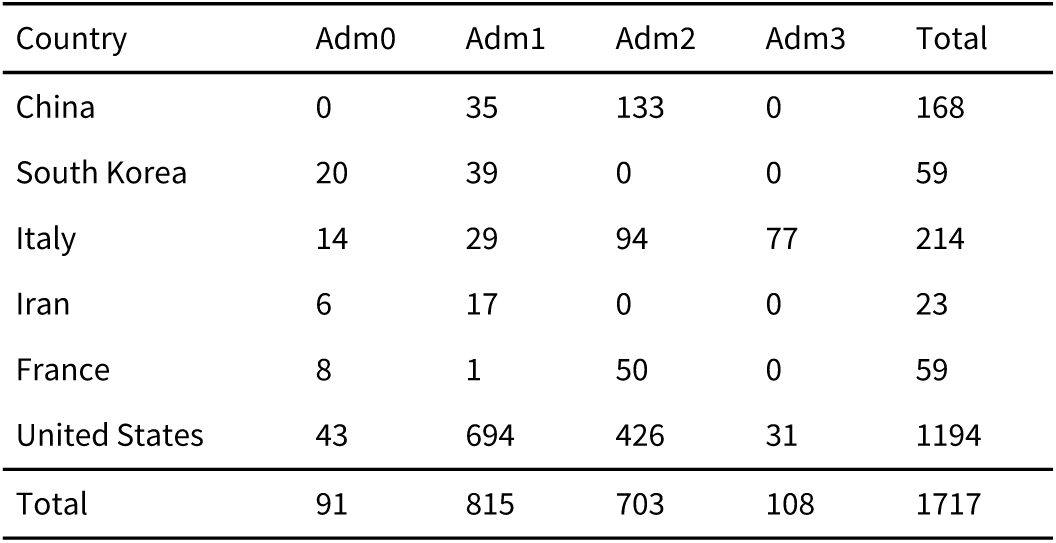
Number of unique anti-contagion policies in this study tabulated by administrative divisions of each country.

**Supplementary Table 2.** Estimates of no-policy infection growth rates and case doubling time using different samples from Wuhan, China. This table reports the raw epidemiological data in Wuhan, China and our estimates of infection growth rates and case doubling times prior to the city-wide lockdown on January 22, 2020, utilizing different sample periods to estimate these values. Column (1) provides the dates of observations. Columns (2) and (3) show raw official epidemiological data: the number of cumulative cases and the number of active cases *(= cumulative cases - recoveries - deaths)*, respectively. We show data from Wu et al. (2020) which match our data in the overlapping periods; in the interest of space, we do not display raw data used in Li et al. (2020), but they are similar with small differences. Columns (4) and (5), report estimates of pre-policy infection growth rates and case doubling times (in days) using different start dates, respectively. Specifically, we set the sample start date as the date in column (1) and the end date of the sample (used for estimating our no-policy growth rate) fixed at January 22, 2020 (the day prior to city-wide lockdown), using the time series data between these two dates to estimate the parameters in columns (4) and (5). Column (6) provides notes on these data. Using a start date of 12/10/19, the same as Wu et al (2020), we obtain an estimated case doubling time of 5.7 days, similar to their estimated 5.2 days (they use an alternative structural modeling approach). As described in our Supplementary Notes on Epidemiological Data in China, we only use data in China beginning 1/16/20 because the first national guidelines for diagnosis were issued on 1/15/20. Prior to that date, there did not exist a consistent case definition to identify the earliest 41 confirmed cases in Wuhan.^249^ Additionally, the documented lack of testing capacity in the province of Hubei before 1/16/20 raises concern about data quality during that time period.^250^ These concerns about data quality appear consistent with irregularities in the official record of cumulative cases (column 2). For example, official cumulative cases decreased on 1/9/20, which should not be possible. Additionally, no new cases were reported between 1/9/20 and 1/15/20, when at least roughly five new cases per day should have been reported if case doubling time actually was 5.2 days. The reliability of these official reports during the 1/9/20-1/15/20 period has been called into question, with news sources suggesting that people who were likely to have been infected by COVID-19 in that period of time (and deaths attributed to the disease) were not counted in the official tally.^251, 252^ These data quality concerns motivate our use of the 1/16/20 start data for our sample in China, which provide an infection growth rate of 0.36 and case doubling time of 1.95 days (gray) using only the time series in Wuhan. These estimates are broadly consistent with our estimates from all other countries we examine, except Iran, and the global average growth rate of 0.37 we estimate (see Figure 2A in the main text).

| (1) | (2) | (3) | (4) | (5) | (6) |
| --- | --- | --- | --- | --- | --- |
| **Date**<br>(used as start of sample) | Raw case data on date in (1)<br>Source: Wu et al. (2020)<sup>247</sup> | Raw case data on date in (1) | Estimates using 1/22/20 as sample end date | Estimates using 1/22/20 as sample end date | **Notes** |
|  | **Cumulative confirmed cases** | **Active cases** | **Estimated infection growth rate** | **Case doubling time (days)** |  |
| 12/10/19 | 1 | 1 | 0.12 | 5.69 | ← Wu et al. (2020) and Li et al. (2020)<sup>248</sup> sample begins |
| 12/11/19 | 1 | 1 | 0.12 | 5.56 |  |
| 12/12/19 | 1 | 1 | 0.13 | 5.42 |  |
| 12/13/19 | 1 | 1 | 0.13 | 5.29 |  |
| 12/14/19 | 2 | 2 | 0.12 | 5.96 |  |
| 12/15/19 | 4 | 4 | 0.10 | 6.88 |  |
| 12/16/19 | 6 | 6 | 0.09 | 7.51 |  |
| 12/17/19 | 6 | 6 | 0.09 | 7.30 |  |
| 12/18/19 | 6 | 6 | 0.10 | 7.10 |  |
| 12/19/19 | 6 | 6 | 0.10 | 6.89 |  |
| 12/20/19 | 7 | 7 | 0.10 | 7.00 |  |
| 12/21/19 | 8 | 8 | 0.10 | 7.08 |  |
| 12/22/19 | 10 | 10 | 0.09 | 7.40 |  |
| 12/23/19 | 11 | 11 | 0.09 | 7.40 |  |
| 12/24/19 | 11 | 11 | 0.10 | 7.15 |  |
| 12/25/19 | 13 | 13 | 0.09 | 7.34 |  |
| 12/26/19 | 13 | 13 | 0.10 | 7.07 |  |
| 12/27/19 | 13 | 13 | 0.10 | 6.80 |  |
| 12/28/19 | 14 | 14 | 0.10 | 6.72 |  |
| 12/29/19 | 18 | 18 | 0.10 | 7.17 |  |
| 12/30/19 | 21 | 21 | 0.09 | 7.37 |  |
| 12/31/19 | 22 | 22 | 0.10 | 7.20 |  |
| 1/1/20 | 28 | 28 | 0.09 | 7.78 |  |
| 1/2/20 | 34 | 34 | 0.08 | 8.30 |  |
| 1/3/20 | 44 | 44 | 0.07 | 9.39 |  |
| 1/4/20 | 44 | 44 | 0.08 | 8.87 | ← Li et al. (2020) sample ends |
| 1/5/20 | 59 | 59 | 0.06 | 10.71 |  |
| 1/6/20 | 59 | 59 | 0.07 | 10.04 |  |
| 1/7/20 | 59 | 59 | 0.07 | 9.37 |  |
| 1/8/20 | 59 | 59 | 0.08 | 8.70 |  |
| 1/9/20 | 41 | 41 | 0.12 | 5.94 | ← Official cumulative cases decreases |
| 1/10/20 | 41 | 38 | 0.13 | 5.17 | ← No new official cases |
| 1/11/20 | 41 | 34 | 0.16 | 4.37 | ← No new official cases |
| 1/12/20 | 41 | 33 | 0.18 | 3.86 | ← No new official cases |
| 1/13/20 | 41 | 33 | 0.20 | 3.43 | ← No new official cases |
| 1/14/20 | 41 | 33 | 0.23 | 3.00 | ← No new official cases |
| 1/15/20 | 41 | 27 | 0.30 | 2.29 | ← National standards for diagnosis issued |
| 1/16/20 | 45 | 28 | 0.36 | 1.95 | ← Testing available in Hubei |
| 1/17/20 | 62 | 41 | 0.35 | 1.98 | Our sample begins (see the Supplementary Notes on Epi. Data in China) |
| 1/18/20 | 121 | 94 | 0.35 | 1.98 |  |
| 1/19/20 | 198 | 170 | 0.27 | 2.58 |  |
| 1/20/20 | 258 | 228 | 0.26 | 2.70 |  |
| 1/21/20 | 365 | 329 | 0.15 | 4.72 |  |
| 1/22/20 | 425 | 381 | - | - | ← Wu et al. (2020) and our sample for estimating no-policy growth rate end |

**Supplementary Table 3.** Country level regression results estimating the effect of policy interventions on COVID-19 infection growth rates. These regression tables **a-f** display the results from our main model estimating the effect of policy on daily COVID-19 infection growth rates in China, South Korea, Italy, Iran, France, and the United States. The regression model is estimated separately for each country, allowing for the policy type to have different average treatment effects for each country.

| Variables | Coefficient | Std Error | P-value |  | Coefficient | Std Error | P-value |  |
| --- | --- | --- | --- | --- | --- | --- | --- | --- |
| *Policy variables:* |  |  |  |  | Week 1 | -0.026 | 0.046 | 0.581 |
| Emergency declaration, Week 1 | -0.012 | 0.043 | 0.792 |  | Week 2 | -0.204*** | 0.049 | 0.000 |
| Emergency declaration, Week 2 | -0.173*** | 0.046 | 0.000 |  | Week 3 | -0.279*** | 0.047 | 0.000 |
| Emergency declaration, Week 3 | -0.234*** | 0.047 | 0.000 |  | Week 4 | -0.302*** | 0.046 | 0.000 |
| Emergency declaration, Week 4 | -0.250*** | 0.047 | 0.000 |  | Week 5 | -0.292*** | 0.047 | 0.000 |
| Emergency declaration, Week 5 | -0.248*** | 0.047 | 0.000 |  |  |  |  |  |
| Travel ban, Week 1 | 0.015 | 0.016 | 0.338 |  |  |  |  |  |
| Travel ban, Week 2 | -0.009 | 0.018 | 0.618 |  |  |  |  |  |
| Travel ban, Week 3 | -0.031 | 0.024 | 0.191 |  |  |  |  |  |
| Travel ban, Week 4 | -0.048 | 0.029 | 0.107 |  |  |  |  |  |
| Travel ban, Week 5 | -0.079** | 0.038 | 0.043 |  |  |  |  |  |
| Home isolation, Week 1 | -0.029** | 0.014 | 0.041 |  |  |  |  |  |
| Home isolation, Week 2 | -0.021 | 0.017 | 0.223 |  |  |  |  |  |
| Home isolation, Week 3 | -0.014 | 0.021 | 0.512 |  |  |  |  |  |
| Home isolation, Week 4 | -0.003 | 0.027 | 0.897 |  |  |  |  |  |
| Home isolation, Week 5 | 0.035 | 0.037 | 0.355 |  |  |  |  |  |
| *Testing regime dummy variables:* |  |  |  |  |  |  |  |  |
| Testing regime change on Jan 18, 2020 | 0.488*** | 0.046 | 0.000 |  |  |  |  |  |
| Testing regime change on Jan 28, 2020 | 0.065*** | 0.015 | 0.000 |  |  |  |  |  |
| Testing regime change on Feb 06, 2020 | -0.039*** | 0.012 | 0.002 |  |  |  |  |  |
| Testing regime change on Feb 13, 2020 | -0.002 | 0.002 | 0.356 |  |  |  |  |  |
| Testing regime change on Feb 20, 2020 | -0.005** | 0.002 | 0.015 |  |  |  |  |  |
| Testing regime change on Mar 05, 2020 | -0.006 | 0.005 | 0.237 |  |  |  |  |  |
| Observations | 3,669 |  |  |  |  |  |  |  |
| R-squared | 0.60 |  |  |  |  |  |  |  |

\*\*\* p<0.01, \*\* p<0.05, \* p<0.1

| Variables | Coefficient | Std Error | P-value |
| --- | --- | --- | --- |
| *Policy variables:* |  |  |  |
| Social distance (optional) | -0.083** | 0.038 | 0.032 |
| Social distance (mandatory) | -0.304** | 0.151 | 0.049 |
| Emergency declaration | -0.127*** | 0.046 | 0.009 |
| Quarantine positive cases | 0.020 | 0.019 | 0.295 |
| *Testing regime dummy variables:* |  |  |  |
| Testing regime change on Feb 20, 2020 | 0.074 | 0.087 | 0.398 |
| Testing regime change on Feb 29, 2020 | 0.040** | 0.019 | 0.044 |
| Testing regime change on Mar 22, 2020 | -0.010 | 0.023 | 0.655 |
| Testing regime change on Mar 27, 2020 | -0.039 | 0.038 | 0.309 |
| Observations | 595 |  |  |
| R-squared | 0.28 |  |  |

\*\*\* p<0.01, \*\* p<0.05, \* p<0.1

| Coefficient | Std Error | P-value |
| --- | --- | --- |
| -0.494*** | 0.150 | 0.002 |

**c. Italy**Dependent variable: growth rate of cumulative confirmed cases ( $\Delta$ log cases per day)
| Variables | Coefficient | Std Error | P-value |
| --- | --- | --- | --- |
| *Policy variables:* |  |  |  |
| Other social distance | 0.144** | 0.068 | 0.040 |
| School closure | -0.109 | 0.070 | 0.127 |
| Travel ban | -0.331** | 0.137 | 0.021 |
| Quarantine positive cases | -0.084 | 0.055 | 0.134 |
| Business closure | -0.121 | 0.098 | 0.225 |
| Home isolation | 0.048 | 0.051 | 0.360 |
| Observations | 2,898 |  |  |
| R-squared | 0.29 |  |  |

\*\*\* p<0.01, \*\* p<0.05, \* p<0.1

| Coefficient | Std Error | P-value |
| --- | --- | --- |
| -0.453*** | 0.105 | 0.000 |

d. Iran Dependent variable: growth rate of cumulative confirmed cases ( $\Delta \log$ cases per day)
| Variables | Coefficient | Std Error | P-value |
| --- | --- | --- | --- |
| *Policy variables:* |  |  |  |
| Travel ban (opt), work from home, school closure | -0.334*** | 0.089 | 0.001 |
| Home isolation | -0.154*** | 0.033 | 0.000 |
| *Testing regime dummy variable:* |  |  |  |
| Testing regime change on Mar 13, 2020 | -0.075 | 0.096 | 0.442 |
| Observations | 548 |  |  |
| R-squared | 0.29 |  |  |

\*\*\* p<0.01, \*\* p<0.05, \* p<0.1

| Coefficient | Std Error | P-value |
| --- | --- | --- |
| -0.487*** | 0.076 | 0.000 |

e. France Dependent variable: growth rate of cumulative confirmed cases ( $\Delta \log$ cases per day)
| Variables | Coefficient | Std Error | P-value |
| --- | --- | --- | --- |
| *Policy variables:* |  |  |  |
| Other social distance | -0.243*** | 0.072 | 0.003 |
| School closure | -0.009 | 0.042 | 0.826 |
| National lockdown | 0.083 | 0.052 | 0.127 |
| *Testing regime dummy variable:* |  |  |  |
| Testing regime change on Mar 15, 2020 | -0.112*** | 0.021 | 0.000 |
| Observations | 270 |  |  |
| R-squared | 0.30 |  |  |

\*\*\* p<0.01, \*\* p<0.05, \* p<0.1

| Coefficient | Std Error | P-value |
| --- | --- | --- |
| -0.169*** | 0.020 | 0.000 |

f. United States Dependent variable: growth rate of cumulative confirmed cases ( $\Delta \log$ cases per day)
| Variables | Coefficient | Std Error | P-value | Coefficient | Std Error | P-value |
| --- | --- | --- | --- | --- | --- | --- |
| *Policy variables:* |  |  |  | -0.380*** | 0.095 | 0.000 |
| Slow the Spread Guidelines (Mar 16 federal) | 0.049 | 0.037 | 0.193 |  |  |  |
| No gathering | 0.011 | 0.035 | 0.750 |  |  |  |
| Social distance | -0.246*** | 0.044 | 0.000 |  |  |  |
| Quarantine positive cases | -0.061* | 0.031 | 0.058 |  |  |  |
| Paid sick leave | -0.027 | 0.087 | 0.761 |  |  |  |
| Work from home | -0.047*** | 0.017 | 0.009 |  |  |  |
| School closure | 0.026 | 0.031 | 0.407 |  |  |  |
| Travel ban | -0.008 | 0.033 | 0.802 |  |  |  |
| Business closure | -0.055** | 0.021 | 0.014 |  |  |  |
| Religious closure | 0.008 | 0.020 | 0.713 |  |  |  |
| Home isolation | -0.030* | 0.017 | 0.096 |  |  |  |
| *Testing regime dummy variables:* |  |  |  |  |  |  |
| Testing regime change on Mar 13, 2020 in NY | -0.137*** | 0.043 | 0.003 |  |  |  |
| Testing regime change on Mar 16, 2020 in CA | -0.021 | 0.024 | 0.388 |  |  |  |
| Testing regime change on Mar 18, 2020 in NC | 0.169*** | 0.042 | 0.000 |  |  |  |
| Testing regime change on Mar 19, 2020 in CT | 0.106** | 0.040 | 0.012 |  |  |  |
| Testing regime change on Mar 19, 2020 in NV | 0.241*** | 0.039 | 0.000 |  |  |  |
| Testing regime change on Mar 19, 2020 in UT | -0.162*** | 0.038 | 0.000 |  |  |  |
| Testing regime change on Mar 20, 2020 in IA | -0.211*** | 0.024 | 0.000 |  |  |  |
| Testing regime change on Mar 21, 2020 in TN | 0.203*** | 0.037 | 0.000 |  |  |  |
| Testing regime change on Mar 22, 2020 in AL | -0.001 | 0.027 | 0.966 |  |  |  |
| Testing regime change on Mar 23, 2020 in HI | -0.034 | 0.024 | 0.162 |  |  |  |
| Testing regime change on Mar 24, 2020 in KS | -0.041** | 0.020 | 0.046 |  |  |  |
| Testing regime change on Mar 24, 2020 in NJ | 0.022 | 0.026 | 0.406 |  |  |  |
| Testing regime change on Mar 25, 2020 in OH | 0.010 | 0.027 | 0.717 |  |  |  |
| Testing regime change on Mar 27, 2020 in AZ | 0.047 | 0.033 | 0.162 |  |  |  |
| Testing regime change on Mar 28, 2020 in MD | 0.006 | 0.029 | 0.833 |  |  |  |
| Testing regime change on Mar 28, 2020 in MO | -0.034 | 0.051 | 0.502 |  |  |  |
| Testing regime change on Mar 30, 2020 in DE | -0.008 | 0.033 | 0.808 |  |  |  |
| Observations | 1,238 |  |  |  |  |  |
| R-squared | 0.29 |  |  |  |  |  |

\*\*\* p<0.01, \*\* p<0.05, \* p<0.1
This regression includes state fixed effects, day-of-week fixed effects, and clustered standard errors at the day level. P-values are from the two-sided test with the null hypothesis that the coefficient is equal to zero. The sample includes 1,238 observations at the state-day level.
"Other social distance" includes policies such as closing libraries, maintaining 6 feet distance from others in public, and limiting visits to long term care facilities.

**Supplementary Table 4.** Estimated Effect of Actual Policies on Infection Growth Rates With and Without Grouping Policies.

| Country | Main Specification (policies grouped) |  | Disaggregated Model (policies separate) |  |
| --- | --- | --- | --- | --- |
|  | Effect size | 95% CI | Effect size | 95% CI |
| China | -0.252 | (-.339, -.164) | -0.252 | (-.339, -.164) |
| South Korea | -0.248 | (-.423, -.073) | -0.21 | (-.377, -.044) |
| Italy | -0.24 | (-.372, -.107) | -0.237 | (-.369, -.104) |
| Iran | -0.355 | (-.48, -.231) | -0.291 | (-.437, -.146) |
| France | -0.123 | (-.16, -.086) | -0.123 | (-.176, -.07) |
| United States | -0.084 | (-.144, -.025) | -0.084 | (-.144, -.024) |

**Supplementary Table 5.**
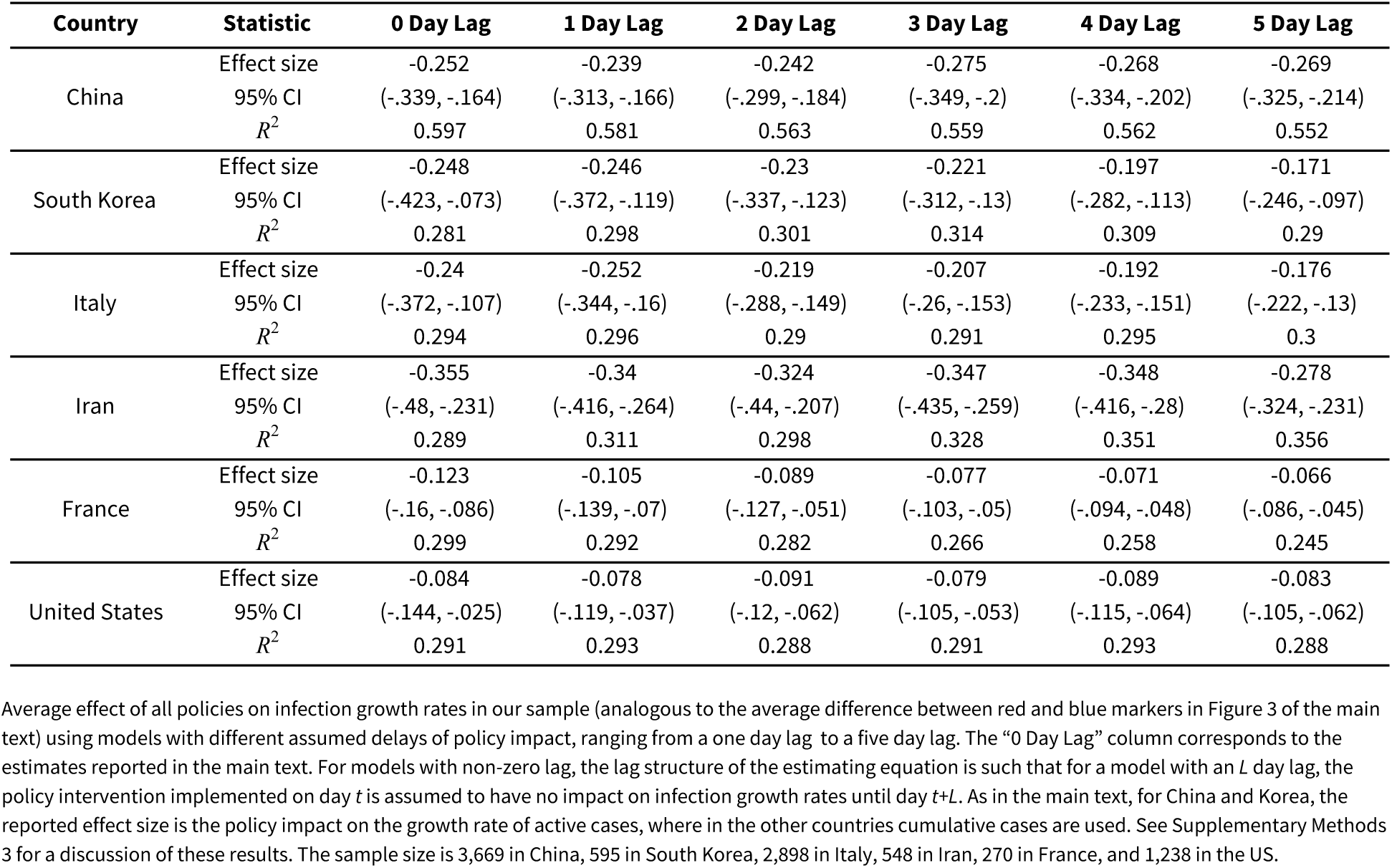
Estimated Effect of Actual Policies Combined on Infection Growth Rates Using Alternative Models that Assume Lagged Effects Of Policies.

| Country | Statistic | 0 Day Lag | 1 Day Lag | 2 Day Lag | 3 Day Lag | 4 Day Lag | 5 Day Lag |
| --- | --- | --- | --- | --- | --- | --- | --- |
| China | Effect size | -0.252 | -0.239 | -0.242 | -0.275 | -0.268 | -0.269 |
|  | 95% CI | (-.339, -.164) | (-.313, -.166) | (-.299, -.184) | (-.349, -.2) | (-.334, -.202) | (-.325, -.214) |
|  | R<sup>2</sup> | 0.597 | 0.581 | 0.563 | 0.559 | 0.562 | 0.552 |
| South Korea | Effect size | -0.248 | -0.246 | -0.23 | -0.221 | -0.197 | -0.171 |
|  | 95% CI | (-.423, -.073) | (-.372, -.119) | (-.337, -.123) | (-.312, -.13) | (-.282, -.113) | (-.246, -.097) |
|  | R<sup>2</sup> | 0.281 | 0.298 | 0.301 | 0.314 | 0.309 | 0.29 |
| Italy | Effect size | -0.24 | -0.252 | -0.219 | -0.207 | -0.192 | -0.176 |
|  | 95% CI | (-.372, -.107) | (-.344, -.16) | (-.288, -.149) | (-.26, -.153) | (-.233, -.151) | (-.222, -.13) |
|  | R<sup>2</sup> | 0.294 | 0.296 | 0.29 | 0.291 | 0.295 | 0.3 |
| Iran | Effect size | -0.355 | -0.34 | -0.324 | -0.347 | -0.348 | -0.278 |
|  | 95% CI | (-.48, -.231) | (-.416, -.264) | (-.44, -.207) | (-.435, -.259) | (-.416, -.28) | (-.324, -.231) |
|  | R<sup>2</sup> | 0.289 | 0.311 | 0.298 | 0.328 | 0.351 | 0.356 |
| France | Effect size | -0.123 | -0.105 | -0.089 | -0.077 | -0.071 | -0.066 |
|  | 95% CI | (-.16, -.086) | (-.139, -.07) | (-.127, -.051) | (-.103, -.05) | (-.094, -.048) | (-.086, -.045) |
|  | R<sup>2</sup> | 0.299 | 0.292 | 0.282 | 0.266 | 0.258 | 0.245 |
| United States | Effect size | -0.084 | -0.078 | -0.091 | -0.079 | -0.089 | -0.083 |
|  | 95% CI | (-.144, -.025) | (-.119, -.037) | (-.12, -.062) | (-.105, -.053) | (-.115, -.064) | (-.105, -.062) |
|  | R<sup>2</sup> | 0.291 | 0.293 | 0.288 | 0.291 | 0.293 | 0.288 |

**Supplementary Table 6.**
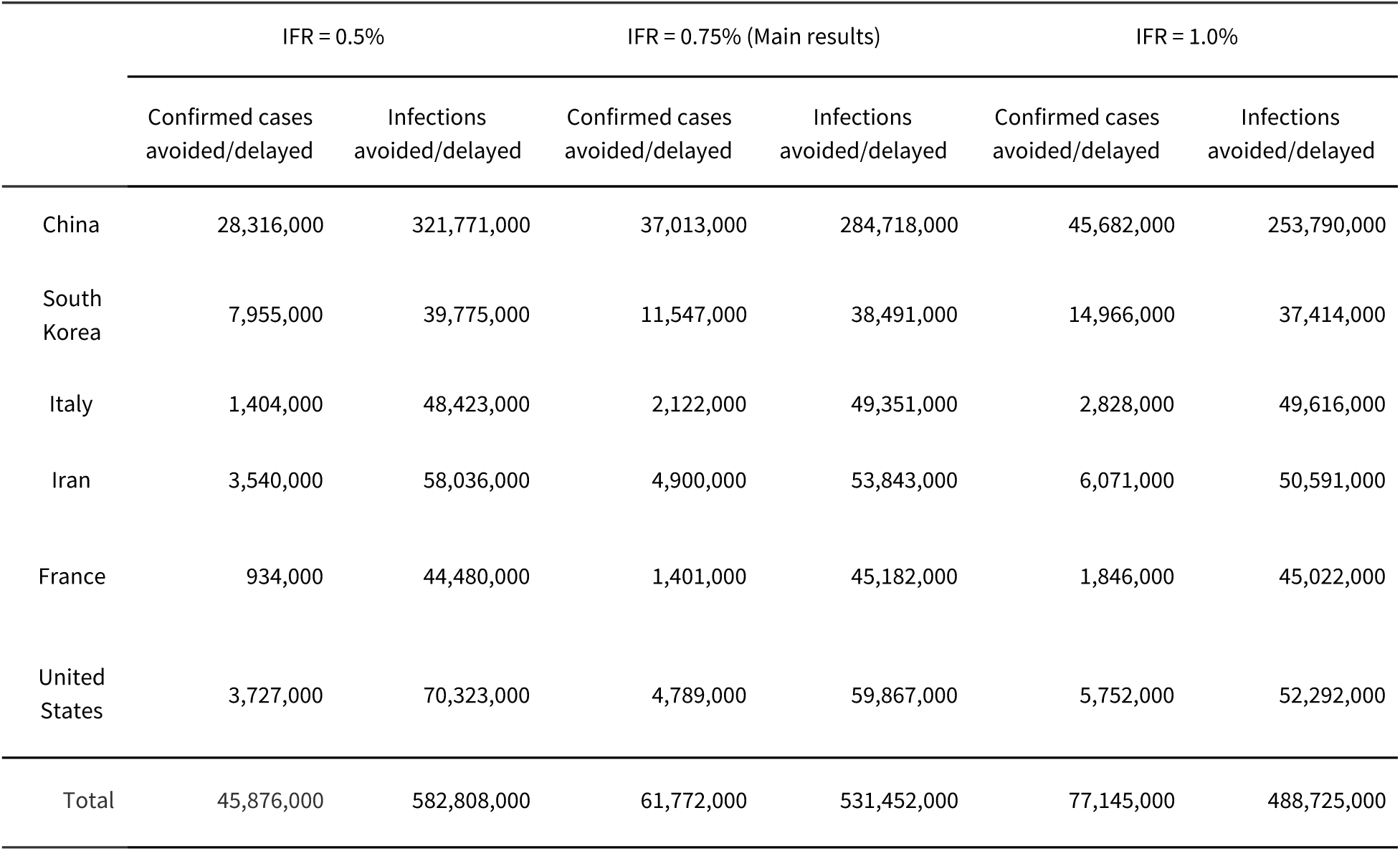
Sensitivity of cases and infections delayed/avoided to assumed infection fatality rate. Total estimated confirmed cases and infections avoided/delayed for assumed infection fatality ratios (IFRs) of 0.5%, 0.75% (main results) and 1.0%. Table shows values on the respective final days of our samples for each of the six countries presented in our analysis. We use the assumed IFRs to compute underreporting estimates following Russell et al.^253^

## Footnotes

1 https://github.com/CSSEGISandData/COVID-19 (access date: April 7, 2020)

2 Russell, T., Joel Hellewell, and S. Abbot. “Using a delay-adjusted case fatality ratio to estimate under-reporting.” *Centre for Mathematical Modelling of Infectious Diseases Repository* (2020). URL: https://cmmid.github.io/topics/covid19/severity/global_cfr_estimates.html (access date: April 18, 2020)

1 Angrist & Pischke, 2008

2 Holland, 1986

3 Mena-Lorcat, 1992

4 Brauer & Chowell, 2012

5 WHO Ebola Response Team

6 Mills et al, 2004

7 Andreason et al, 2008

8 Nishiura et al, 2010

9 Towers et al, 2014

10 WHO Ebola Response Team

11 Ma et al, 2020

12 Muniz-Rodriguez et al, 2020

13 Here we list some examples from Xiaogan, Hubei: 孝感市新型冠状病毒感染的肺炎防控指挥部(3号令); 孝感市新型冠 状病毒感染的肺炎防控指挥部(4号令); 孝感市新型冠状病毒感染的肺炎防控指挥部(8号令); 孝感市新型冠状病毒感 染的肺炎防控指挥部(9号令); (Policy document No. 3, 4, 8, and 9 from the COVID-19 Prevention and Control Center in Xiaogan, Hubei)

14 Tian, Huaiyu, Yonghong Liu, Yidan Li, Chieh-Hsi Wu, Bin Chen, Moritz UG Kraemer, Bingying Li et al. “An investigation of transmission control measures during the first 50 days of the COVID-19 epidemic in China.” *Science* (2020).

15 http://www.gov.cn/xinwen/2020-01/29/content_5472881.htm

16 Tian, Huaiyu, Yonghong Liu, Yidan Li, Chieh-Hsi Wu, Bin Chen, Moritz UG Kraemer, Bingying Li et al. “An investigation of transmission control measures during the first 50 days of the COVID-19 epidemic in China.” *Science* (2020).

17 List | Press Release | News Room: KCDC

18 The case definition of 2019 novel coronavirus will be expanded | Press Release | News Room: KCDC

19 S. Korea’s 31st virus patient still undergoing treatment: KCDC

20 List | Press Release | News Room: KCDC

21 제주도, 신천지 시설 7곳 폐쇄조치…신도 현황은 ‘깜깜’ (Jeju shut down 7 Shincheonji places… information on believers still unknown)

22 Coronavirus latest: South Korea declares red alert after rapid spike in cases

23 *“*대규모로 일어나고 있는 신천지 집단 감염 사태 이전과 이후는 전혀 다른 상황입니다*…*정부와 지방자치단체, 방역당국과 의료진, 나아가 지역주민과 전국민이 혼연일체가 되어 총력 대응해야 하는 중차대한 시점입니다*.”*

24 전문]文대통령 “신천지 집단감염 사태 전·후는 전혀 다른 상황” (President Moon “The situation is entirely different after the Shincheonji outbreak”)

25 South Korea Raises Threat Alert Level

26 [전문]文대통령 “신천지 집단감염 사태 전·후는 전혀 다른 상황” (President Moon “The situation is entirely different after the Shincheonji outbreak”)

27 South Korea Raises Threat Alert Level

28 List | Press Release | News Room: KCDC

29 Moon declares virus-hit Daegu, part of North Gyeongsang Province as special disaster zones

30 *“* 정부는 *3*월 *19*일(목*) 0*시부터 특별입국절차 적용대상을 국내의 모든 내*·*외국인 입국자로 확대하기로 하였다*…* 이는 최근 입국자 검역 과정에서 발생한 다수의 확진 사례 와 전 세계적인 코로나*19* 전파 속도 등을 고려하여 해외 위험요인이 국내로 재유입되는 것을 강력하게 차단하기 위한 조치이다*.” (Translation: “ The government decided to expand Special Entry Procedure to all Koreans and foreign nationals entering Korea from March 19… In consideration of the number of cases confirmed from quarantining inbound travelers, and the speed of global transmission, this measure is designed to strongly block the inflow of overseas risk factors into Korea.”)*

31 코로나바이러스감염증-19 중앙재난안전대책본부 정례브리핑 (3월 17일) (Coronavirus Infectious Diseases-19 Regular Disaster Prevention Headquarters Regular Briefing (March 17)

32 Korea Implements Special Entry Procedure | Official Korea Tourism Organization

33 The updates on COVID-19 in Korea as of 23 March | Press Release | News Room: KCDC

34 The updates on COVID-19 in Korea as of 27 March | Press Release | News Room: KCDC

35 The updates on COVID-19 in Korea as of 31 March | Press Release | News Room: KCDC

36 Coronavirus, primi due casi in Italia: sono due turisti cinesi (Coronavirus, first two cases in Italy: from two Chineses tourists)

37 Factbox: Latest on coronavirus spreading in China and beyond

38 https://metro.co.uk/2020/02/25/towns-ITA-lockdown-coronavirus-12298246/

39 Italian towns on lockdown after 2 virus deaths, clusters

40 Italy orders closure of all schools and universities due to coronavirus

41 Italy Locks Down Much of the Country’s North Over the Coronavirus

42 Coronavirus: Italy extends emergency measures nationwide

43 Coronavirus, negozi e locali chiusi in tutta Italia fino al 25 marzo. Garantiti servizi essenziali, alimentari e farmacie. Possibili riduzioni trasporti. Conte: “Torneremo ad abbracciarci” (Coronavirus, Conte: “In Italy until March 25, shops except food and pharmacies are closed. Possible reductions on transport. Effects for 14 days)

44 Virus News: Italian Industry Shuts Down

45 Coronavirus: Lombardy region announces stricter measures

46 Italy, Pandemic’s New Epicenter, Has Lessons for the World

47 How Iran Became a New Epicenter of the Coronavirus Outbreak

48 For example, early in the pandemic, officials appeared confident in the country’s ability to manage the crisis. But, this quickly gave way to a reportedly uncoordinated policy response: *“Iranian health officials initially boasted of their public health prowess. They ridiculed quarantines as ‘archaic’ and portrayed Iran as a global role model. President Hassan Rouhani suggested a week ago that by this past Saturday life would have returned to normal…* *Embarrassed anew by the spread of the disease, the Iranian authorities have responded with a hodgepodge of contradictory measures mixing elements of a crackdown with attempts to save face.”* Iran’s Coronavirus Response: Pride, Paranoia, Secrecy, Chaos (Accessed 5/5/20)

49 For example, the New York Times reported on infighting between the civilian government (led by President Rouhani) and the military after the Supreme Leader Khamenei authorized the military to take over control of the country’s coronavirus response: *“While [Khamenei] told the military to work with the civilian government, Mr. Khamenei effectively authorized it to sideline Mr. Rouhani’s government if needed. Almost instantly the infighting and conflicting public messaging began…* *Mr. Rouhani demanded that the armed forces adhere to his command. The generals refused and said Mr. Khamenei had authorized them to act independently*. *The military men proposed closing off Tehran and vast swaths of the country… Mr. Rouhani refused…* *Apparently ignoring Mr. Rouhani, General Bagheri said on Friday [March 13, 2020] that the military would severely restrict traffic, close businesses and clear people off the streets in Tehran and at least 11 provinces… Barely two days later, Mr. Rouhani said there would be no shutting of cities, no lockdowns, no forced closure of businesses…* *By Monday, Mr. Rouhani appeared to have dissuaded the military from imposing strict rules. But local authorities defied him, independently closing provinces and several cities.”* Power Struggle Hampers Iran’s Coronavirus Response (Accessed 5/5/20)

50 Revolutionary Guards to enforce coronavirus controls in Iran

51 Orsan plan

52 PRÉPARATION AU RISQUE ÉPIDÉMIQUE Covid-19

53 Ferguson, N., Cummings, D., Cauchemez, S. et al. Strategies for containing an emerging influenza pandemic in Southeast Asia. Nature 437, 209–214 (2005). Ferguson, N., Cummings, D., Fraser, C. et al. Strategies for mitigating an influenza pandemic. Nature 442, 448–452 (2006). Luca, G.D., Kerckhove, K.V., Coletti, P. et al. The impact of regular school closure on seasonal influenza epidemics: a data-driven spatial transmission model for Belgium. BMC Infect Dis 18, 29 (2018).

54 “ *Il est dès lors apparu indispensable au Conseil scientifique de prendre en compte comme objectif collectif principal et immédiat la réduction maximale de l’afflux prévisible de cas graves en réanimation*.” (Translation: “It therefore appeared essential to the Scientific Council to set the main goal as the maximum reduction of the foreseeable influx of serious cases in intensive care.”) Avis du Conseil scientifique COVID-19 16 mars 2020 Avis du Conseil scientifique COVID-19 12 mars 2020

55 First Case of 2019 Novel Coronavirus in the United States

56 Three months in: A timeline of how COVID-19 has unfolded in the US

57 Remarks at Coronavirus Press Briefing | Alex M. Azar II | Press | January 28, 2020 Washington, D.C.

58 Secretary Azar Declares Public Health Emergency for United States for 2019 Novel Coronavirus

59 In the press briefing detailing these actions, Robert Redfield, director of the CDC, focused on the number of cases internationally, saying: *“As of today, there are nearly 9,700 cases in China, with more than 200 deaths. Additionally, currently there are another 23 countries that have confirmed, totally, 132 cases. This also includes 12 individuals who have been confirmed in six countries who did not travel to China.”* Press Briefing by Members of the President’s Coronavirus Task Force | January 31, 2020

60 Suspension of Entry as Immigrants and Nonimmigrants of Certain Additional Persons Who Pose a Risk of Transmitting 2019 Novel Coronavirus

61 Suspension of Entry as Immigrants and Nonimmigrants of Certain Additional Persons Who Pose a Risk of Transmitting 2019 Novel Coronavirus

62 15 Days to Slow the Spread

63 White House Takes New Line After Dire Report on Death Toll

64 Impact of non-pharmaceutical interventions (NPIs) to reduce COVID-19 mortality and healthcare demand

65 Inslee signs bill package to support state effort combating the COVID-19 outbreak

66 Inslee announces statewide shutdown of restaurants, bars and expanded social gathering limits

67 “Stay Home, Stay Healthy” address transcript

68 CDE School Guidance for COVID-19

69 Gov. Gavin Newsom orders all of California to shelter in place

70 A timeline of Cuomo’s and Trump’s responses to coronavirus outbreak | April 3, 2020 | ABC News

71 Amid Lack of Federal Direction, Governor Cuomo, Governor Murphy and Governor Lamont Announce Regional Approach to Combatting COVID-19

72 Get Your Mass Gatherings or Large Community Events Ready

73 Audio & Rush Transcript: Amid Lack of Federal Direction, Governor Cuomo, Governor Murphy and Governor Lamont Announce Regional Approach to Combatting Covid-19

74 Amid Lack of Federal Direction, Governor Cuomo, Governor Murphy and Governor Lamont Announce Regional Approach to Combatting COVID-19

75 All Ohio Bars and Restaurants Ordered Closed at 9 p.m. and Until Further Notice

76 Coronavirus - Governor Whitmer Signs Executive Order Temporarily Closing Bars, Theaters, Casinos, and Other Public Spaces; Limiting Restaurants to Delivery and Carry-Out Orders

77 Pritzker closes bars and restaurants as federal officials to double staff to handle crush at O’Hare

78 Gov. Baker Closes Schools, Bans Mid-Size Gatherings, Eating at Restaurants

79 2020 Executive Orders

80 Kentucky’s Response to COVID-19

81 全球新冠病毒最新实时疫情地图 (The latest real-time global COVID-19 map)

82 BlankerL/DXY-COVID-19-Data: 2019新型冠状病毒疫情时间序列数据仓库 (COVID-19/2019-nCoV Infection Time Series Data Warehouse)

83 疫情通报 (National Health Commision of PRC; COVID-19 Report)

84 信息发布--湖北省卫生健康委员会 (Information release - Hubei Provincial Health Commission)

85 广东省卫生健康委员会网站 (Information Release - Guangdong Provincial Health Commission)

86 浙江省人民政府门户网站疫情通告 (Information Release - Zhejiang Provincial Health Commission)

87 CSSEGISandData/COVID-19: Novel Coronavirus (COVID-19) Cases, provided by JHU CSSE

88 ‘Confusion breeds distrust:’ China keeps changing how it counts coronavirus cases

89 Impact of changing case definitions for COVID-19 on the epidemic curve and transmission parameters in mainland China

90 Impact of changing case definitions for COVID-19 on the epidemic curve and transmission parameters in mainland China

91 Impact of changing case definitions for COVID-19 on the epidemic curve and transmission parameters in mainland China

92 Why China’s Huge Increase in New COVID-19 Cases Is Actually a Step in the Right Direction. We have found another data source indicating that this case definition change happened on February 5 (Impact of changing case definitions for COVID-19 on the epidemic curve and transmission parameters in mainland China); we control for both dates.

93 Impact of changing case definitions for COVID-19 on the epidemic curve and transmission parameters in mainland China Affiliation; China records 2 straight days of fewer than 1,000 new COVID-19 cases

94 Impact of changing case definitions for COVID-19 on the epidemic curve and transmission parameters in mainland China

95 서울특별시 코로나19 발생현황 (Seoul COVID-19 Status)

96 대구광역시 코로나19 확진자 추이 (Daegu COVID-19 The Confirmed Cases)

97 경상북도 코로나19 발생동향 (Gyeongsangbuk-do COVID-19 Status)

98 전라북도 코로나19 일일상황보고 (Jeollabuk-do COVID-19 Daily Reports)

99 세종특별자치시 코로나19 현황판 (Sejong COVID-19 Status)

100 강원도청 (Gangwon-do Provincial Government)

101 춘천시 코로나19 현황 (Chuncheon COVID-19 Status)

102 원주시 코로나19 현황 (Wonju COVID-19 Status)

103 강릉시 코로나바이러스감염증-19 비상대책 (Gangneung COVID-19 Emergency Plan)

104 태백시청 (Taebaek City Government)

105 속초시청 (Sokcho City Government)

106 삼척시청 (Samcheok City Government)

107 경기도 코로나19 발생동향 (Gyeonggi-do COVID-19 Status)

108 인천광역시 코로나19 상황판 (Incheon COVID-19 Status)

109 부산광역시 코로나19 발생현황 (Busan COVID-19 Status)

110 울산광역시 코로나19 발생현황 (Ulsan COVID-19 Status)

111 광주광역시청 (Gwangju COVID-19 Status)

112 충청남도 코로나19 발생현황 (Chungcheongnam-do COVID-19 Status)

113 충청북도 코로나19 발생현황 (Chungcheongbuk-do COVID-19 Status)

114 코로나19 경상남도 현황 (COVID-19 Gyeongsangnam-do Status)

115 제주특별자치도 코로나19 현황 (Jeju COVID-19 Status)

116 전라남도 코로나19 현황 (Jeollanam-do COVID-19 Status)

117 List | Press Release | News Room: KCDC

118 중국 후베이성 우한시 폐렴환자 집단발생 | 보도자료 | 알림·자료 (Pneumonia Outbreak in Wuhan City, Hubei, China)

119 신종코로나바이러스감염증 국내 발생 현황(1월 26일, 사례정의 확대) | 보도자료 | 알림·자료 (COVID-19 Domestic Status (Jan 26, Case Definition Broadened))

120 NB: The KCDC English website explains the testing regime change in a more condensed format: “Any citizens identified with a fever or respiratory symptoms and have visited Wuhan will be isolated and tested at a nationally designated isolation hospital, and any foreigners staying in Korea will be conducted in cooperation with police.” Urges cooperation in preventing the spread of 2019-nCoV in community | Press Release | News Room: KCDC

121 림 > 보도자료 내용보기 “ 신종 코로나바이러스 감염증 대응지침 일부 변경 “ (Revision in the Guidance Documents for COVID-19)

122 The updates on novel Coronavirus in Korea (since 3 January) | Press Release | News Room: KCDC NB: The date of this press release is February 8, 2020, but the definition of “suspected cases” was effective starting from February 7, 2020.

123 NB: The testing fee was already somewhat affordable; a person needed to pay 160,000 KRW (about $130 USD). A related article can be found here: 5 신종코로나 진단검사 비용은 얼마? (How much is the COVID-19 testing fee?)

124 신종 코로나바이러스감염증 중앙사고수습본부 정례 브리핑 (2월 7일) (Daily briefing on COVID-19, February 7)

125 The updates of COVID-19(as of Feb.19) in Korea | Press Release | News Room: KCDC

126 The updates of COVID-19 in Korea (As of 29 Feb. 2020)

127 List | Press Release | News Room: KCDC

128 The updates on COVID-19 in Korea as of 27 March | Press Release | News Room: KCDC

129 2020 coronavirus pandemic in Iran

130 Example of Ministry of Health data: شناسایی 1209 بیمار جدید مبتلا به کووید 19 در کشور (Identification of 1209 new patients with COVID-19 in the country)

131 Google Translate sometimes translates various Persian numbers as “1”. Persian numbers compared here: Persian numbers

132 ANOTHER senior Iranian official dies from coronavirus

133 Revolutionary Guards to enforce coronavirus controls in Iran

134 Fr-SARS-CoV-2

135 Infection à coronavirus

136 Points de situation coronavirus COVID-19

137 France 3 Régions: Actualités

138 Agence régionale de santé | Agir pour la santé de tous

139 Infection au nouveau Coronavirus (SARS-CoV-2), COVID-19, France et Monde

140 https://www.data.gouv.fr/fr/datasets/donnees-hospitalieres-relatives-a-lepidemie-de-covid-19/

141 Coronavirus: en quoi consiste le « stade 3 » de l’épidémie ?

142 CSSEGISandData/COVID-19: Novel Coronavirus (COVID-19) Cases, provided by JHU CSSE

143 2020 Hubei lockdowns

144 Tian, Huaiyu, Yonghong Liu, Yidan Li, Chieh-Hsi Wu, Bin Chen, Moritz UG Kraemer, Bingying Li et al. “An investigation of transmission control measures during the first 50 days of the COVID-19 epidemic in China.” *Science* (2020).

145 http://www.gov.cn/xinwen/2020-01/29/content_5472881.htm

146 China Extends Lunar New Year Holiday to Feb 2, Shanghai to Feb 9

147 코로나19 여파 “사회복지 이용시설 휴관 권고” (Social welfare facilities recommended to shut down)

148 부산 지역아동센터 모두 휴관…더 외로운 저소득층 아이들 (Busan child-care facilities shut down - worse for the lower-income children)

149 서울시, 노인복지관 등 사회복지시설 3601곳 휴관 (Seoul, 3601 social welfare facilities shut down)

150 코로나19 확산을 막기 위한 서울시 일일보고 (Seoul daily report on limiting the spread of COVID-19)

151 (브리핑) 이재명, “PC방·노래연습장·클럽형태업소에 밀접이용제한 행정명령” (Cyber cafes, karaokes, and clubs under the administrative order limiting close-distance usage)

152 19 > 뉴스 & 이슈 > 보도자료 내용보기 “ [카드뉴스] 중앙재난안전대책본부 정례브리핑(3.14.), 특별재난지역선포(대구, 경북 경산·청도·봉화) “ (Daily briefing on announcing the emergency declaration for the regions: Daegu; Gyeongsangbuk-do Gyeongsan, Cheongdo, Bonghwa)

153 보도자료 조회 “인천시, 인천애뜰 잠정 사용중단(금지) 조치” (Incheon prohibits usage of Incheon City Government Square)

154 코로나19 확산 방지를 위해 도심 집회 제한 강화 (Stronger limits on demonstrations in downtown)

155 신천지 관련시설 폐쇄조치, 확산 방지에 행정력 집중…대구시 경찰청과 긴밀히 협조 (Shincheonji-related facilities shut down, Daegu struggling to limit the spread of the virus with the police power)

156 경북, 신천지 1612명 중 221명 확진···31번이 156명 옮겼다 (Gyeongsangbuk-do, 221 out of 1612 tested positive, the 31st patient responsible for infecting 156 people)

157 서울시, 신천지 집회 시설 폐쇄 결정 (Seoul shuts down Shincheonji-related facilities)

158 제주 신천지 신도 전원 능동감시 종료…집회 금지는 유지 (Shincheonji believers now free from monitoring, still religious gatherings prohibited)

159 경기도, 신천지 353개 시설 14일간 강제폐쇄·집회금지 조치 내려 (Gyeonggi-do shuts down 353 Shincheonji facilities for 14 days)

160 광주일보 “전남도, 신천지 교회·시설 58곳 강제폐쇄 행정명령 발동” (Jeollanam-do shuts down 58 Shincheonji-related facilities)

161 여성조선 “경남, 신천지 시설폐쇄 및 집회 금지 행정명령 발동” (Gyeongsangnam-do shuts down Shincheonji facilities and forbids religious gatherings)

162 인천시, 신천지교회 종교시설 추가 폐쇄조치 시행 | 기관 소식 | 정책·정보 (Incheon shuts down more Shincheonji facilities)

163 울산시, 신천지교회 및 부속기관 폐쇄 조치 (Ulsan shuts down Shincheonji facilities)

164 [코로나19] 부산, 신천지 시설 폐쇄·집회 금지 2주 추가 연장 (Busan shuts down Shincheonji facilities for two weeks more)

165 전북 신천지 시설 폐쇄·집회 금지 연장… (Jeollabuk-do extends the period of shutting down Shincheonji facilities)

166 충북 신천지 시설 38개소 폐쇄‧방역 완료 (Chungcheongbuk-do shuts down 38 Shincheonji facilities)

167 광주광역시. 신천지 시설 폐쇄 행정명령 (Gwangju shuts down Shincheonji facilities)

168 충남도, 신천지 관련 시설 58개소 폐쇄 (Chungcheongnam-do shuts down 58 Shincheonji facilities)

169 대전광역시 신천지 시설 방역 및 폐쇄조치 현황입니다. 신천지 신도 및 교육생 현황입니다. (Status report on Shincheonji facilities shutdown, and Shincheonji believers and trainees)

170 ‘신종 코로나 확산’ 2월 취소 행사 확인하세요! (Event cancellation in February due to COVID-19)

171 코로나19(Covid-19) 확산 2~3월 취소 행사 확인하세요! (Event cancellation in February and March due to COVID-19)

172 연합뉴스 “강원 5명 코로나19 확진..공공시설 출입제한·행사 연기·취소” (Five confirmed cases in Gangwon-do, public facilities shutdown, events delayed or canceled)

173 충북도, 코로나19 확산될라…행사 줄줄이 취소 (Chungcheongbuk-do cancels events due to COVID-19)

174 ‘신종코로나 유입 막자’…충남 대규모 체육·문화 행사 줄취소(종합) (Chungcheongnam-do cancels events due to COVID-19)

175 세종시 신종 코로나여파 각종 행사 취소 및 자제요청 (Sejong urges cancellation of events amid COVID-19 outbreak)

176 ‘심각단계’ 격상 코로나19 대응 시정브리핑 (The alert level raised, COVID-19 daily briefing)

177 신종 코로나바이러스 여파로 경북도내 각종 축제·행사 취소 또는 연기 (Gyeongsangbuk-do cancels or delays events due to COVID-19)

178 신종 코로나 확산에 경남 지역행사 등 줄줄이 취소 (Gyeongsangnam-do cancels events due to COVID-19)

179 제주도내 행사 등 전면 취소, “코로나19 확산 방지 우선” (Jeju cancels events due to COVID-19)

180 신종 코로나바이러스 감염증 대응을 위한 도내 각종 행사 취소․축소 방침 (Gyeonggi-do cancels events due to COVID-19)

181 울산지역 주요행사 잇따라 취소·연기 (Ulsan cancels or delays events due to COVID-19)

182 코로나바이러스감염증-19 대응 관련 취소 행사 현황 (2.28. 현재) (The list of events canceled due to COVID-19)

183 2020년 정월대보름 관측행사 취소 안내 (Daeboreum events canceled)

184 인천시, 코로나19 확산방지 강력조치 (Incheon strict policies for limiting the spread of the virus)

185 신종코로나 확산…전남 지자체, 행사 줄줄이 취소 (Jeollanam-do cancels events)

186 송하진 도지사, 코로나바이러스 대응 ‘올인’ (Governor of Jeollabuk-do makes every effort to fight against the virus)

187 전국 모든 유·초·중·고·특 개학 2주간 추가연기 결정 (코로나19) (All kindergarten, elementary schools, middle schools, and high schools are closed for two more weeks)

188 The updates of COVID-19 in Korea, February 29

189 The updates of COVID-19 in Korea as of 22 February

190 The case definition of 2019 novel coronavirus will be expanded | Press Release | News Room: KCDC

191 Expand strict quarantine screening of 2019-nCoV to Hong Kong, Macao | Press Release | News Room: KCDC

192 The updates on COVID-19 in Korea as of 11 March | Press Release | News Room: KCDC

193 Updates on COVID-19 in Korea (as of 12 March)

194 The updates on COVID-19 in Korea as of 16 March | Press Release | News Room: KCDC

195 목록 | 보도자료 | 알림·자료 (The list of press release)

196 최신 여행경보단계 조정 (The latest adjustment on the travel alert levels)

197 최신 여행경보단계 조정 (The latest adjustment on the travel alert levels)

198 일본 전 지역(후쿠시마 원전 주변지역 제외)에 여행경보 2단계(황색경보, 여행자제)로 상향 조정 (All Japanese region, other than the Fukushima nuclear reactor area, now under the level 2 travel alert)

199 최신 여행경보단계 조정 (The latest adjustment on the travel alert levels)

200 The updates on COVID-19 in Korea as of 18 March

201 여행경보제도 소개 (The description on the travel alert policy)

202 홍콩 여행경보 2단계(여행자제)로 상향 조정 (Now travels to Hong Kong under the level two alert)

203 기니의 여행경보단계 상향 조정 (The alert level is raised against travels to Guinea)

204 The updates on COVID-19 in Korea as of 11 March | Press Release | News Room: KCDC

205 The updates on COVID-19 in Korea as of 22 March | Press Release | News Room: KCDC

206 The updates on COVID-19 in Korea as of 23 March | Press Release | News Room: KCDC

207 The updates on COVID-19 in Korea as of 27 March | Press Release | News Room: KCDC

208 The updates on COVID-19 in Korea as of 31 March | Press Release | News Room: KCDC

209 The updates on COVID-19 in Korea as of 5 April | Press Release | News Room: KCDC

210 Chronology of main steps and legal acts taken by the Italian Government for the containment of the COVID-19 epidemiological emergency

211 Virus News: Italian Industry Shuts Down

212 Italy Coronavirus News: Travel Ban Inside Country

213 Coronavirus: Lombardy region announces stricter measures

214 UPDATED: Timeline of the Coronavirus

215 2020 coronavirus pandemic in Iran

216 How Iran Became a New Epicenter of the Coronavirus Outbreak

217 How Iran Became a New Epicenter of the Coronavirus Outbreak

218 Revolutionary Guards to enforce coronavirus controls in Iran

219 Info Coronavirus COVID-19

220 Agence régionale de santé | Agir pour la santé de tous

221 Pandémie de Covid-19 en France

222 Info Coronavirus COVID-19

223 Confinement de 2020 en France

224 Décret n° 2020-293 du 23 mars 2020 prescrivant les mesures générales nécessaires pour faire face à l’épidémie de covid-19 dans le cadre de l’état d’urgence sanitaire

225 What’s New | COVID | CDC

226 Community Mitigation Strategies | CDC

227 15 Days to Slow the Spread

228 Schools, Workplaces & Community Locations | CDC

229 National Governors Association

230 CDC Travel Guidance & Warnings

231 US Travel Warnings

232 CDC COVID-19 Guidance Documents

233 NYT Article | “Wondering About Social Distancing?”

234 COVID-19 | Get Your Mass Gatherings or Large Community Events Ready for Coronavirus Disease 2019

235 US School Closures due to COVID-19

236 NYT Article | “See Which States and Cities Have Told Residents to Stay at Home”

237 행정구역(시군구)별, 성별 인구수 (Population by county and gender)

238 Iran: Administrative Division (Provinces and Counties) - Population Statistics, Charts and Map

239 Population de 1999 à 2020

240 datamade/census: A Python wrapper for the US Census API.

241 Epidemiology and Transmission of COVID-19 in Shenzhen China: Analysis of 391 cases and 1,286 of their close contacts

242 Using a delay-adjusted case fatality ratio to estimate under-reporting

243 A systematic review and meta-analysis of published research data on COVID-19 infection-fatality rates

244 Estimating epidemic exponential growth rate and basic reproduction number

245 MIDAS Network Online COVID-19 Portal: Parameter Estimates

246 Estimating epidemic exponential growth rate and basic reproduction number

247 Wu, J.T., Leung, K., Bushman, M. et al. Estimating clinical severity of COVID-19 from the transmission dynamics in Wuhan, China. Nature Medicine 26, 506–510 (2020). https://doi.org/10.1038/s41591-020-0822-7

248 Li, Qun, Xuhua Guan, Peng Wu, Xiaoye Wang, Lei Zhou, Yeqing Tong, Ruiqi Ren et al. “Early transmission dynamics in Wuhan, China, of novel coronavirus–infected pneumonia.” New England Journal of Medicine (2020).

249 Impact of changing case definitions for COVID-19 on the epidemic curve and transmission parameters in mainland China Affiliation

250 武汉肺炎:疫情从可控到失控的三十天 (Wuhan pneumonia: 30 days from outbreak to out of control)

251 Warning against cover-up as China virus cases jump

252 As families tell of pneumonia-like deaths in Wuhan, some wonder if China virus count is too low

253 Using a delay-adjusted case fatality ratio to estimate under-reporting

## Notes

### Competing Interest Statement

The authors have declared no competing interest.

